# Using LIBRA-seq to map the BK-polyomavirus specific B-cell response in kidney transplant recipients

**DOI:** 10.64898/2026.02.03.26345220

**Authors:** S. Marchand, A. Trochel, M. Loirat, J. Mignon, T. Letellier, M. Braud, L. Delbos, C. Fourgeux, S. Taouli, C. Peltier, L. Gautreau-Rolland, J. Poschmann, G. Blancho, X. Saulquin, C. Bressollette-Bodin, D. McIlroy

## Abstract

BK polyomavirus (BKPyV) is a major complication in kidney transplant recipients (KTR), for whom no specific antiviral therapy is available. Modulation of immunosuppressive therapy results in virus clearance in most KTR with BKPyV DNAemia (controllers), but a significant minority fail to clear the virus (non-controllers). Here, we adapt LIBRA-seq, which links antibody sequence data to antigen specificity, to intact viral capsids of the four BKPyV genotypes to study and compare BKPyV-specific B-cell repertoires in controllers (n=8) versus non-controllers (n=3). Sequences were obtained for 5197 BKPyV-specific antibodies, and predicted antigen specificities were validated by ELISA and neutralizing assays (n=24 antibodies). Patterns of cross-reactivity were reproducible between individuals, and B-cells binding multiple BKPyV genotypes were present in all patients. However, the specific subset of B-cells enriched for expression of broadly neutralizing antibodies made up only 1.2% of the BKPyV-specific repertoire, and was not detected in all patient samples. The proportions of broadly-specific and isotype switched antibodies, and rates of somatic hypermutation increased with time following peak DNAemia, and were comparable in both patient groups, indicating that there is no identifiable deficit in the humoral response mounted by BKPyV non-controllers, and supporting the notion that humoral immunity alone is insufficient to control established BKPyV replication. This work shows that LIBRA-seq can be successfully applied to intact virus capsids and provides a framework for analyzing antiviral B-cell repertoires and antibody specificity in clinically relevant settings.

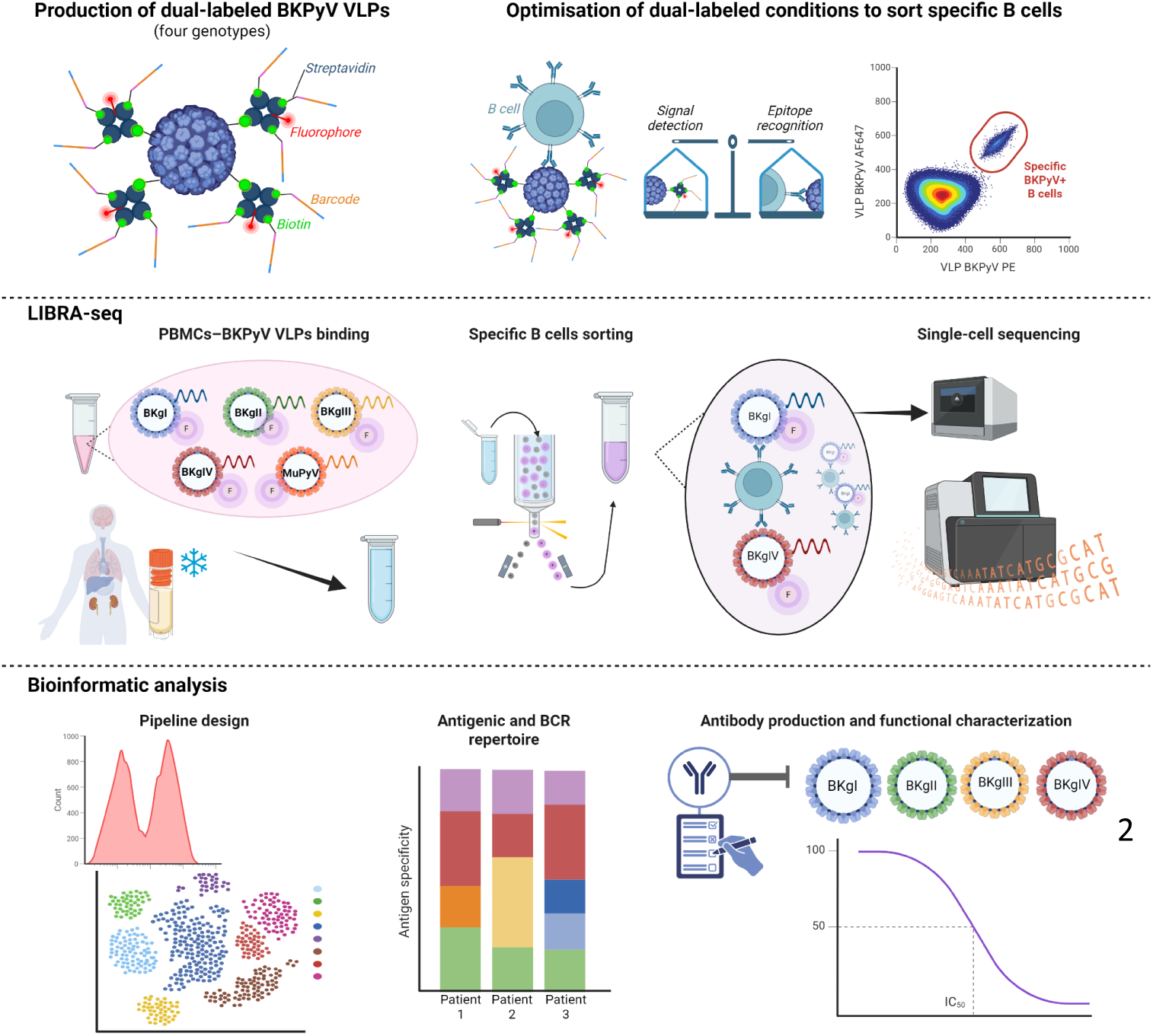

## INTRODUCTION

The identification and characterization of neutralizing antibodies is a major focus in antiviral immunity, particularly in view of their therapeutic and prophylactic potential against viral infections (1,2). With the onset of the COVID-19 pandemic, many research teams focused on understanding the host’s response to SARS-CoV-2 by isolating and producing monoclonal antibodies, providing a boost to the research on mAbs for other viral infections (3–5). Other previous studies had extensively investigated the humoral response and B-cell receptor (BCR) repertoires in patients with HIV, HCMV, influenza virus or RSV infections (6–9). These studies, focused on enveloped viruses, have significantly advanced our understanding of antibody diversity, antigen recognition, affinity maturation, and the development of immune memory, thereby shaping our current knowledge of antiviral humoral responses in both infection and vaccination contexts.

Despite these advances, the characterization of BCR repertoires targeting non-enveloped viruses remains limited. Many non-enveloped viruses such as adenoviruses, enteroviruses, and polyomaviruses can cause severe disease, especially in immunosuppressed patients, for whom virus-specific therapeutic options are lacking. One such virus is BK polyomavirus (BKPyV), a small, non-enveloped double-stranded DNA virus that reactivates under immunosuppression, particularly in kidney transplant recipients (KTR). High levels of BKPyV replication in the renal epithelium of transplanted kidneys can lead to polyomavirus-associated nephropathy (PyVAN), resulting in deteriorating graft function and, potentially, graft loss (10,11). While therapeutic options remain limited, mAbs may represent a viable adjuvant to current clinical practice, which relies on modulation of immunosuppressive treatment (12). Significant progress has been made in this area, with the characterization of several BKPyV-specific broadly neutralizing antibodies (bNAbs) (13–17), two of which have advanced to clinical trials with the aim of treating persistent BKPyV-DNAemia in KTR (18,19). However, the role of antibodies in the clearance of BKPyV in this clinical context is not firmly established. BKPyV DNAemia in KTR is managed by modulation of immunosuppressive therapy, following which, patients - including those who do not clear the virus - develop high anti-BKPyV antibody titers (20,21), suggesting that the humoral response has a limited role in the suppression of BKPyV replication once high levels of DNAemia have been reached. On the other hand, persistent BKPyV DNAemia is characterized by the accumulation of mutations in the major capsid protein, VP1 (22,23) which are associated with neutralization escape (24,25), implying that the humoral response does indeed exert significant selection pressure on the virus. This raises important questions about the quality of the humoral response in patients with persistent BKPyV DNAemia, suggesting that it may lack sufficient breadth or diversity, and thereby be more susceptible to viral escape variants. In this context, understanding the clonal composition and functionality of the BKPyV-specific B-cell repertoire in patients who control virus replication, as well as in those who fail to do so, could help to identify the features of a protective response and contribute to the rational use of bNAbs in the control of BKPyV DNAemia.

The diversity and complexity of the BCR repertoire generated during viral infection present a challenge for researchers, due to the rarity of antigen-specific cells in the total B-cell population. However, recent technological advances have revolutionized study methodologies, with a notable advance being the development of LIBRA-seq, a high-throughput sequencing technology that links BCR sequences to antigen specificity, by labeling antigens with both fluorophores and oligonucleotide barcode probes (6). After sorting B-cells based on fluorescence, both heavy-and light-chain BCR sequences and antigen-associated barcodes are identified via scRNA-seq using the 10X Genomics platform. This approach allows for the direct detection of cross-reactive antibodies by multiplexing diverse antigens, or variants of the same antigen, each tagged with unique barcodes to ensure their identification. Through this dual analysis, LIBRA-seq offers a highly detailed picture of the B-cell response to specific antigens.

To date, this methodology has been used to study of enveloped viruses such as HIV, SARS-CoV-2, influenza virus, and hepatitis C virus, focusing on surface glycoproteins as antigens (6,26–28), and has so far only been applied to one non-enveloped virus (29). Here we develop and validate a dual fluorescence and oligonucleotide labeling technique for polyomavirus virus-like particles (VLPs), which is generalisable to any non-enveloped virion, then use it to interrogate the BKPyV-specific B-cell repertoires in a cohort of KTR, stratified according to their ability to control BKPyV-DNAemia. The goals of this study were to: compare the clonal and transcriptional profiles of BKPyV-specific B-cells between viremic controllers and non-controllers, in order to identify immunological correlates of viral control and features of an effective humoral response; and to develop an analytical approach capable of identifying broadly neutralizing antibody candidates. To our knowledge, this work represents the first adaptation of LIBRA-seq to a complete virus capsid and provides a high-resolution map of the BKPyV-specific B-cell landscape in transplant patients.

## MATERIALS AND METHODS

### 1) Patient samples

Patients included in this study were transplanted between 2010-2013, and between 2021-2023. All patients gave informed consent authorizing the use of blood samples for research purposes. All clinical and biological data were extracted from the hospital databases and anonymized for this study, according to the recommendation of the CNIL (Commission Nationale de l’Informatique et des Libertés). The 23 patients in this study were diagnosed with BKPyV reactivation based on the detection of viruria >10⁷ copies/ml. A total of 27 cryopreserved peripheral blood mononuclear cells (PBMC) samples were selected following peak viral load, including two longitudinal samples from four patients. Due to insufficient numbers of BKPyV-specific B-cells in some samples, scRNA-seq analysis was ultimately performed on 20 samples corresponding to 17 patients.

### 2) Cell culture

HEK 293TT cells, obtained from the National Cancer Institute’s Developmental Therapeutics Program (Frederick, Maryland, USA), were cultured in DMEM High Glucose (ThermoFisher) containing 10% FBS (ThermoFisher), 100 U/mL penicillin, 100 µg/mL streptomycin (Dutscher), 1x Glutamax (ThermoFisher), and 250 µg/mL Hygromycin B (Sigma-Aldrich). Cells were incubated at 37°C with 5% CO2 in a humidified incubator. At confluence, cells were passaged by trypsinization for 10 minutes with 1x TrypLE Express (ThermoFisher).

### 3) Plasmids

The BKPyV VP2 and VP3 expression plasmids ph2b (#32109) and ph3b (#32110), as well as the murine polyomavirus (MuPyV) VP1 plasmid (#22519) were purchased from Addgene (Cambridge, MA). VP1 expression plasmids encoding BKPyV genotypes Ib2, II, III and IVc2, and the phGLuc reporter plasmid were kindly provided by Dr Christopher Buck, National Cancer Institute (NCI), Bethesda, MD (30). The BK BKgIb2 VP1 plasmid was modified by site-directed mutagenesis to introduce the K73E and D82E substitutions, in order to generate a VP1 sequence more representative of the majority of circulating variants. The modified BKgIb2-EE plasmid was used to prepare BKgI VLPs and pseudovirus (PSV).

### 4) Barcoded oligonucleotides

The barcoded oligonucleotides used have the following structure: 5’-CCTTGGCACCCGAGAATTCCAXXXXXXXXXXXXXXXCCCATATAAGA*A*A-3’. The 5’ region represents a small RNA Read 2 (Read 2S), Xs represent a 15 nucleotide feature barcode BEAM (10X Genomics), and the 3’ region represents the capture sequence. 5’ biotinylated barcoded oligonucleotides were supplied by Eurofins Genomics.

### 5) VLP production

The workflow for dual-labeled BKPyV production is shown in S1 Fig.

MuPyV and BKPyV VLPs were prepared according to the protocols of the Buck laboratory with minor modifications (31). Approximately 8-10×10^6^ HEK 293TT cells were seeded into a 75 cm2 flask containing DMEM supplemented with 10% FBS and no antibiotics. Transfection was performed using the Lipofectamine 2000 reagent (Invitrogen) according to the manufacturer’s instruction with 40 µg VP1 plasmid and 80 µL Lipofectamine 2000 per T75 flask.

48 hours post-transfection, cells were harvested by trypsinization, washed once with PBS and resuspended in a volume of PBS equal to the pellet volume then mixed with 0,4× volume of 25 U/ml type V neuraminidase (Sigma-Aldrich) and incubated at 37°C for 15 minutes. To lyse the cells, 0,125× volume of 10% Triton X-100 (Sigma-Aldrich) was added and incubated at 37°C for 45 minutes. The pH of the lysate was then adjusted by the addition of 0,075× volume of 1 M sodium bicarbonate and 1 μl of 250 U/μl Pierce Nuclease (Pierce) was added to degrade free DNA. After incubation for 2 hours at 37°C, the lysates were adjusted to 0,8 M NaCl, incubated for 10 minutes on ice and then centrifuged at 5000*g* for 5 minutes at 4°C. The supernatant was carefully transferred to a new tube and the pellet was resuspended in two pellet volumes of PBS containing 0,8M NaCl, followed by another round of centrifugation.

The lysate was carefully layered on an OptiPrep gradient consisting of 27%/33%/39% (Sigma-Aldrich) prepared in DPBS/0,8 M NaCl. The sample was then centrifuged overnight at 175000*g* at 4°C using a Sw55TI rotor (Beckman). After centrifugation, the tubes were punctured with a 25G syringe needle and 12 fractions of approximately 250 µL from each gradient were collected in 1,5 ml microcentrifuge tubes. A portion of each fraction (10 μL) was retained for SDS-PAGE analysis to identify peak fractions, which were then pooled, exchanged 3x in PBS and concentrated using 100kDa Amicon Ultra centrifugal filter units (Merck). The VP1 concentration of each purified and concentrated MuPyV and BKPyV VLP was determined by loading 10 μl onto a 10% polyacrylamide gel. After gel electrophoresis, the gel was revealed with PageBlue protein staining solution (ThermoFisher) for 1 hour at room temperature. Quantification was performed using BSA standards migrated on the same gel and Image Lab software (Biorad).

### 6) VLP biotinylation

Purified MuPyV and BKPyV VLPs were biotinylated for 30 minutes at room temperature using the EZ-Link™ Sulfo-NHS-LC-Biotinylation kit (ThermoFisher). Different concentrations of Sulfo-NHS-LC-Biotin were tested with the aim of obtaining between 1-3 biotin groups per VP1, and the final protocol used a stoichiometric Sulfo-NHS-LC-Biotin:VP1 ratio of 6,67.

In order to quantify the Sulfo-NHS-LC-Biotin added on MuPyV and BKPyV VLPs, 10 μl was loaded onto a 10% polyacrylamide gel, electrophoresed and transferred to nitrocellulose membrane by using Trans-Blot Turbo RTA Nitrocellulose Transfer Kit (Biorad). The blot was revealed with streptavidin-peroxidase (Jackson ImmunoResearch). Quantification was performed using biotinylated HLA class I protein (P2R platform, SFR Bonamy, Nantes, France) standards and Image Lab software (Biorad).

Biotinylated MuPyV and BKPyV VLP morphology was confirmed at the electron microscopy platform of SC3M (Nantes Université, France).

### 7) Dual-labeling (fluorescent and oligonucleotide barcoded) of antigens

The 5’-biotinylated barcode oligonucleotides were incubated with streptavidin-conjugated fluorophore (PE, AF647 or AF488, Biolegend) at a 3:1 molar ratio for 30 minutes at room temperature. Then, biotinylated MuPyV and BKPyV VLPs were incubated with streptavidin-fluorophore-oligonucleotide complexes. Different streptavidin:VP1 ratios were tested, either to bind from 36 streptavidin-oligonucleotide complexes per VLP (ratio=0.1, R0.1) up to ∼350 per VLP (ratio=0.9, R0.9), depending on the molecular weight and the concentration of the VLP. The binding reaction was incubated for 30 minutes at room temperature and protected from light, leading to the generation of dual labeled (fluorescent and oligonucleotide barcoded) VLPs.

### 8) Immunological and biological validation of dual-labeled (fluorescent and oligonucleotide barcoded) VLPs

#### *a)* ELISA

An ELISA was performed using two antibodies, 41F17 (13) and P8D11 (16), that bind to distinct epitopes on the capsid, and recognize all four BKPyV genotypes, and a negative control (IgG1). Nunc MaxiSorp 96-well plate (Sigma-Aldrich) was initially coated with 50 ng of MuPyV and BKPyV VLPs in 50 μl of PBS at 37°C overnight, followed by blocking with 5% powdered milk in PBS for 1 hour. Antibodies were then added to the plate at five-fold serial dilutions starting at 4 μg/ml in the blocking buffer and incubated 1 hour at 37°C. Antibodies bound to VLPs were detected using a goat anti-human IgG peroxidase-conjugated secondary antibody (Jackson ImmunoResearch) diluted 1:5000 in blocking buffer for 1 hour at 37°C. Washes were performed between each step with PBS-0,05% Tween-20 (Sigma-Aldrich). Then, 50 μL of TMB substrate (BD Biosciences) was added and 5 minutes later, the reaction was stopped by the addition of 50 μL of 0,5 M H_2_SO_4_. Absorbance was measured at 450 nm using a TECAN Spark reader.

#### ***b)*** Binding to HEK 293TT

Cells were trypsinized for 10-12 minutes at 37°C, and resuspended in OptiPRO (Thermofisher) + 25 mM HEPES pH 7,5 (Sigma-Aldrich), 0,1% BSA (Sigma-Aldrich) at 2 million cells/mL. Cells were then seeded in a 96-well V-bottom plate at 50 μL per well and incubated for 30 minutes at 4°C protected from light with 2µg/mL dual-labeled MuPyV and BKPyV VLPs prepared with different VP1:streptavidin-oligonucleotide complex ratios (R0,9, and 3-fold dilutions). After staining, the cells were washed three times with PBS containing 0,5% FBS by centrifugation at 1000*g* for 1 minute each time. Fluorescence intensity was measured using a Canto II flow cytometer (Becton Dickinson), and flow cytometry data were analyzed using FlowJo VX software (BD Life Sciences).

### 9) Antigen-specific B cell labeling and FACS sorting

Cryopreserved PBMC aliquots were thawed by gentle agitation in a 37°C water bath, then transferred to a 15 mL conical tube containing 10 mL of warm RPMI 1640 with 20% FBS. After centrifugation at 400*g* for 10 minutes at room temperature, the supernatant was removed and the cell pellet was resuspended in 1 mL of warm OptiPRO. The cells were then mixed with 40 µL of 25 U/ml type V neuraminidase and incubated at 37°C for 30 minutes. The cells were centrifuged at 400*g* for 10 minutes at 4°C. B-cells were isolated using B cell isolation kit II (Miltenyi Biotec), according to the manufacturer’s protocol. Briefly, after counting the cells, they were resuspended in PBS 1x, pH 7.2, 0,5% BSA, 2 mM EDTA (Fluka), 100 U/mL penicillin and 100 µg/mL streptomycin. Biotin-Antibody cocktail and Anti-Biotin MicroBeads were added in order to target non B-cells, which were separated magnetically using the AutoMACS Pro Separator with “DepleteS” program following the manufacturer’s instructions (Miltenyi Biotec). Enriched B-cells were collected and counted, then centrifuged at 400*g* for 8 minutes at 4°C and resuspended in 100 µL of cold buffer containing 1X DPBS (Sigma-Aldrich) with 1% FBS, 1 mM EDTA and 25 mM HEPES. Each sample was labeled with FACS anti-human antibodies diluted 1/80 (anti-CD3-BV510, anti-CD19-PE-Cy7, anti-CD16-PerCP-Cy5.5, anti-CD14-PerCP-Cy5.5, BD Pharmingen)and the cocktail of dual-lableled VLPs at 1 µg/ml each (BKPyV-gI-VLP-AF647, BKPyV-gI-VLP-PE, BKPyV-gII-VLP-AF647, BKPyV-gII-VLP-PE, BKPyV-gIII-VLP-AF647, BKPyV-gIII-VLP-PE, BKPyV-gIV-VLP-AF647, BKPyV-gIV-VLP-PE, MuPyV-VLP-AF488). Specific hashtag anti-human TotalSeq-C oligonucleotide-labeled antibodies (BioLegend) were added into the mix at 2,5 µg/ml. The mix was incubated for 30 minutes at 4°C protected from the light. Cells were washed three times in 10 mL of cold buffer consisting of 1X DPBS supplemented with 1% FBS, 1 mM EDTA, and 25 mM HEPES, followed by centrifugation at 400*g* for 8 minutes at 4°C. After the final wash, cells were resuspended in 300 µL of the same buffer and pooled. A DAPI (1:2000) viability staining solution (BD Pharmingen) was added, and the pool was incubated for 5 minutes in the dark. The pooled samples were sorted using an Aurora CS (Cytek Biosciences) for experiments 1 and 4, and an Aria FACS sorter (Becton-Dickinson) for experiments 2 and 3. Purity sort-mode was used for experiments 1 and 2, whereas yield sort-mode was selected for experiments 3 and 4. Sorted cells were collected differently depending on the experiment. For experiment 1, cells were collected into a 15-mL Falcon tube pre-coated with FBS, washed once by adding cold 1X PBS, briefly centrifuged, and subsequently resuspended in 500 µL of cold 1X PBS. For experiments 2, 3, and 4, sorted cells were collected into 1X PBS containing BSA (5%) and RNase inhibitor (Sigma Aldrich) in a well of a 96-well plate. The plate was centrifuged at 2500 rpm for 2 min, after which the cells were resuspended in 47 µL of cold 1X PBS. Regardless of the collection method, all sorted cells were immediately processed for single-cell RNA-seq.

### 10) Single cell library preparation and sequencing

Cells were loaded onto a Next GEM Chip K (PN-1000286) and run-on Chromium Single Cell Controller using the Chromium Single Cell 5′ V2 Next GEM single cell kit (PN-1000263) with 5′ feature barcode technology (PN-1000541) according to the manufacturer’s instructions (CG000330, 10× Genomics). Gex, Cell Surface, BEAM and VDJ libraries were prepared and sequenced on an S1 flow cell on a Nova-Seq 6000 (Illumina) at the GenoBiRD platform (IRS-UN, CHU Nantes) and libraries are demultiplexed with bcl2fastq2 (v2.20) in FASTQ format.

### 11) Bioinformatic analysis

FASTQ files were analyzed with Cellranger (7.2.0) to identify cellular barcodes, BEAM, Cell Surface (ADT/HTO) and VDJ, as well as align the 5’GEX reads to the human transcriptome (hg38) and then count UMIs for each gene in order to obtain a count matrix (RNA, BEAM, Cell Surface) and BCR annotation files (VDJ). Count matrices were integrated and analyzed in R script using Seurat (v5.2.1) (32). Data were filtered to retain only high quality cells according to the following criteria: number of genes per cell < 200; number of reads per cell > 4000; mitochondrial genes < 5% of total UMI; no BCR doublet (IGH/IGK/IGL). Genes expressed in fewer than 3 cells were also removed from the analysis.

For antigen specificity analysis, VLP barcode UMI counts for each cell were imported into R, log transformed, and the background log10 MuPyV UMI count was subtracted from all the log10 BKPyV columns, which were then visualized as a UMAP projection. To define clusters of cells with similar antigen-binding properties, the Leiden algorithm was implemented using the igraph package (33), by first converting the background-subtracted log-UMI count distance matrix to a k=20 nearest neighbours graph, then performing Leiden clustering with varying resolution, using the clustree package (34) to visualize the stability of clusters as a function of resolution. Data from each experiment was analyzed separately, due to the differences in UMI count signal between runs.

To annotate clusters, the first step was to assume that background-subtracted log UMI counts for each VLP barcode had a bimodal distribution, then to determine the cutoff value between positive and negative cells using the mixtools package (35), so as to label each cell as “positive” or “negative” for each BKPyV VLP probe. Secondly, the proportion of positive cells for each VLP probe was determined for all Leiden clusters, with each cluster being designated as positive for a particular VLP probe when the majority of cells within the cluster were labelled as “positive”. Binding of different BKPyV genotypes was then summarized by the *Beam_code* variable, for example, *Beam_code = 1* represents a cluster binding only genotype I BKPyV, *Beam_code = 12* represents a cluster binding genotypes I and II, and so on. Finally, all cells within the cluster were labelled according to the *Beam_code* of the cluster. In some experiments, certain probes did not display a clear bimodal distribution due to substantial overlap between negative and positive populations. In experiments 1 and 2, all probes were included, except for the BKgIII-AF647 probe in experiment 1, for which the barcode oligonucleotide was subsequently found to lack biotinylation. In experiments 3 and 4, only AF647-labeled probes were used, as PE-labeled VLPs showed discordant, non-bimodal UMI count distributions, with substantial overlap between negative and positive populations, and were therefore excluded from the analysis. Sub-clustering of the *Beam_code = 1234* cluster was done using data from AF647-labeled probes.

The B-cell subtypes of BKPyV-specific B-cells were determined from 5’Gex data, after pooling and normalizing data from the four experiments using the SCTransform method and integration with Seurat. Cell clusters were identified using Seurat’s FindAllMarkers function. Differentially expressed genes were classified as upregulated (avg_log2FC > 0, adjusted p-value < 0.05) or downregulated (avg_log2FC < 0, adjusted p-value < 0.05). Functional annotation of each cluster was performed based on the top differentially expressed genes.

The Immcantation suite of tools was used for BCR repertoire analysis, firstly by importing data from the filtered_contig.fasta and filtered_contig_annotations.csv files from each experiment, then assigning V, D, and J genes using IgBLAST to construct an AIRR compliant.tsv file compatible with downstream Immcantation packages. An experiment ID column was added, then concatenated with the cell barcode to give a unique_cell_id identifier. The.tsv files from each experiment were then merged before the identification of clonotypes using the hierarchicalClones function in SCOPer (36) using the Density method to determine the threshold distance, and only_heavy = TRUE, split_light = TRUE for paired heavy and light chains. SHM was analyzed using the SHazaM package (37), and BCR trees were built with Dowser (38). Figures illustrating various aspects of the data were prepared with ggplot2 (39).

### 12) Production of monoclonal antibodies

Monoclonal antibodies to BKPyV were produced as previously described (40). The selected paired variable heavy (VH) and light chain (VL) sequences were sent to Eurofins for gene synthesis. VH and VL sequences were cut from the plasmid backbone by restriction enzymes prior to cloning into expression vectors containing constant regions of heavy chain (Cγ1 of IgG1) and light kappa chain (Cκ) or light lambda chain (Cλ). Cloned expression vectors were confirmed by Sanger sequencing, then plasmid Maxipreps were prepared.

Antibodies were first produced on a small scale to check specificity. The day before transfection, 1.5×10^4^ HEK 293A cells were seeded in 96-well plates in 200 μL DMEM supplemented with 1% Glutamax, 10% FBS. 125 ng of vH and 125 ng of vL expression vectors were diluted in 25 µL of 150 mM NaCl, then mixed with 0.5 µL transfection reagent jetPEI (Polyplus) diluted in 25 µL of 150 mM NaCl. After 15 minutes of incubation at room temperature, the complex was gently added onto pre-plated 293A cells. After 16h post transfection, the medium was replaced with serum-free Pro293a medium (Lonza) to avoid serum-Igs contamination. Cells were cultured for 5 days at 37°C in a humidified 5% CO2 incubator, then supernatants were harvested and centrifuged at 460*g* for 5 minutes to eliminate cells and debris. The presence of antibody in supernatant was confirmed by ELISA using Affinity Purified goat anti-human IgG Fc Fragment (BD Biosciences) to coat plates and goat anti-human IgG horseradish peroxidase-conjugated (BD Biosciences) as secondary antibody.

Antibodies with confirmed VLP binding were scaled up for production and purification. Briefly, the day before transfection, 6×10^6^ HEK 293A cells seeded into 175 cm2 flasks were transfected with 10 µg of vH and 10 µg of vL expression vectors following jetPEI DNA transfection protocol. At 5 days post-transfection, supernatants were harvested and centrifuged at 460*g* for 5 minutes to remove cells, then filtered using a 1,22 µm filter, then a 0,45 µm filter prior to purification.

Antibodies were purified using a 1 mL HiTrap rProtein A Fast Flow column (Sigma-Aldrich) on a fast protein liquid chromatography (FPLC) system (Bio-Rad). First, the protein A sepharose column was equilibrated with 20 mM pH 7,2 phosphate buffer. Filtered supernatant containing antibodies was loaded onto the column, then washed with 20 mM pH 7,2 phosphate buffer, and finally eluted with 0,1 M pH 3 citrate buffer. 500 µL of each fraction was collected into tubes containing 1 M pH 9 Tris buffer. Optical density was read at 280 nm on a spectrophotometer (Eppendorf) to determine peak fractions to pool. Pooled fractions were dialyzed in a Slide-A-Lyser cassette (ThermoFisher) with a 3,5K molecular weight cutoff against PBS overnight at 4°C with agitation, then sterilized by filtration at 0,2 µm. Antibody purity was verified by size-exclusion chromatography using Superdex200 Increase 10/300 GL column (GE Healthcare) following the manufacturer’s instructions.

### 13) Pseudotype production and neutralization assays

BKPyV PSV particles were prepared following the protocols developed by the Buck lab with slight modifications (31). Briefly, cell preparation and transfection were performed similarly to BKPyV VLP production using a total of 36 µg plasmid DNA consisting of 16 µg VP1 plasmid, 4 µg ph2b, 8 µg ph3b and 8 µg phGLuc for transfection of HEK 293TT cells. After 48h of transfection, producer cells were collected by trypsinization, then cells were resuspended in PBS 9,5 mM MgCl_2_ then treated with 1 U/mL Type V neuraminidase (Sigma) for 30 minutes at 37°C before lysis with 0.5% Triton X100, and digestion of nucleic acids with Pierce nuclease (Pierce). After overnight incubation at 37°C, the lysate was clarified by centrifuging twice at 5000*g* for 5 minutes at 4°C before purifying PSV on an Optiprep gradient as described for VLP production. After ultracentrifugation and fraction collection, 5 µL of each fraction was diluted to 50 µL in dH_2_O containing 250 µg/ml proteinase K (Qiagen). This solution was incubated at 50°C for 60 minutes followed by 95°C for 10 min, then 1 µL of this extract was used for qPCR using Applied Biosystems 2x SYBR Green Mix (Applied Biosystems). Primers were phGLuc-F 5’-GGTCGAGACCGGGCCTTT-3’ and phGLuc-R 5’-GGGTGGCAGGTATTAGGG-3’. Thermal cycling was initiated with a first denaturation step at 95°C for 10 min, followed by 35 cycles of 95°C for 15 sec and 55°C for 40 seconds. Standard curves were constructed using serial dilutions from 10^7^ to 10^2^ copies of the phGluc plasmid per tube. Peak fractions were pooled, BSA was added to a final concentration of 0,1%, then PSV were aliquoted and stored at-80°C until use in neutralization assays.

Human serum samples were heat inactivated for 30 minutes at 56°C. Each experiment contained four wells of cells receiving diluted PSV without test serum (100% control) and six wells with cells that received only culture medium (negative control). Also, the human anti-BKPyV monoclonal antibody BK120 (15) was used in each experiment as a neutralization control.

HEK 293TT cells were plated at a density of 10^4^ cells per well in a volume of 100µL, then after overnight culture PSV of BKgIb2, BKgII, BKgIII and BKgIV were diluted in DMEM supplemented with 25 mM HEPES pH 7,4 and 0,1% BSA to a final concentration of 5×10^7^ pGLuc copies/mL. Inactivated sera were incubated with PSV in 5-fold dilutions, starting at 1:50 for patient serum samples, 25 µg/ml for expressed monoclonal antibodies, and 1 µg/ml for the BK120 positive control. The plate was incubated 1 hour at 4°C, before adding 100µL of the mixture to cells. After an incubation of five days at 37°C, cells were lysed by pipetting 50µL/well of 1% Triton X100 in PBS, and 10µL of the lysate were transferred into a black plate with the luciferase substrate (Nano-Glo Luciferase Assay System, Promega). Half-maximal inhibitory concentrations (IC50) of the human serum samples or expressed monoclonal antibodies were determined by using GraphPad Prism 10 (GraphPad Software).

## Results

### 1. Optimization of BKPyV VLP probes for LIBRA-seq

To extend the applicability of LIBRA-seq beyond enveloped viruses, we developed a dual-labeled VLP system for BK polyomavirus (BKPyV). The first step was to biotinylate BKPyV VLPs and a murine PyV VLP, to be used as a negative control in B-cell sorting experiments. SDS-PAGE analysis of biotinylated VLP samples revealed a single protein band corresponding to VP1, migrating between approximately 40 and 43 kDa depending on the BKPyV genotype, and at 42 kDa for murine VLPs. Purity exceeded 90%, with yields ranging from 25 to 84 µg (Fig 1A). Consistent biotinylation across VLP preparations, with estimated biotin-to-VP1 ratios ranging from 1 to 4 (Fig 1B) was confirmed by Western blot using biotinylated HLA class I protein as a quantitation standard. Negative-stain electron microscopy revealed a homogeneous polyomavirus-like morphology of the biotinylated VLPs (Fig 1C).

**Fig 1.**
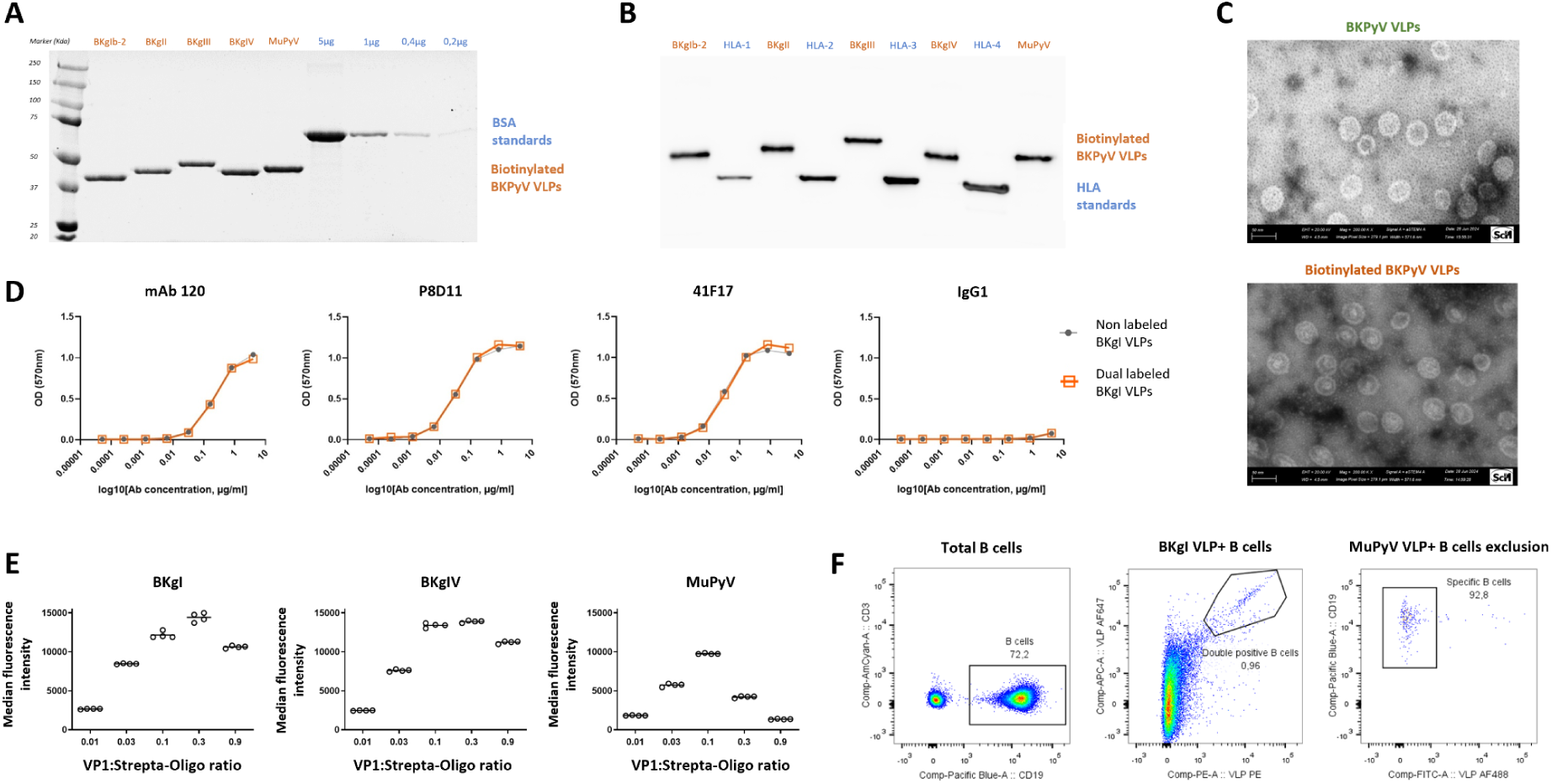
Characterization and validation of biotin-labeled BKPyV and MuPyV VLPs. (A) Coomassie Blue SDS–PAGE gel of biotin-labeled BKPyV and MuPyV VLPs. Increasing amounts of BSA were included as standard controls. (B) Western blot of biotin-labeled BKPyV and MuPyV VLPs. Increasing amounts of biotinylated HLA class I protein were used as quantitation standards. (C) Negative-stain electron microscopy of unlabeled BKPyV VLPs and biotin-labeled BKPyV VLPs. (D) ELISA performed on BKPyV VLPs and dual-labeled BKPyV VLPs using three monoclonal antibodies recognizing all four BKPyV genotypes (mAb120, 41F17, and P8D11), with IgG1 used as a negative control. (E) Increasing amounts of streptavidin–oligonucleotide complexes (R = 0,9 and serial 3-fold dilutions) were added to VLPs, followed by incubation of dual-labeled VLPs with HEK 293TT cells. Binding was analyzed by flow cytometry. Bars represent the median, and each condition was performed in quadruplicate. (F) Dual-labeled BKPyV and MuPyV VLPs were incubated with PBMC samples, and antigen-specific B-cells were analyzed by flow cytometry. After gating on lymphocytes, doublets and non-B cell populations (CD3+CD14+CD16+) were excluded, followed by gating on CD19⁺ B-cells. Within this population, cells negative for MuPyV-AF488 and double positive for BKPyV-PE and-AF647 VLPs were quantified.

In the second step, biotinylated VLPs were conjugated with a streptavidin–fluorophore–oligonucleotide complex to enable dual labeling. Because such modification could conceivably interfere with antibody binding, we validated the antigenicity of the particles. Dual-labeled VLPs displayed equivalent binding to three broadly neutralizing monoclonal antibodies compared with unmodified VLPs, confirming that epitopes remained accessible (Fig 1D).

We next optimized the labeling conditions using HEK 293TT cells to determine the optimal streptavidin-to-VP1 ratio for the detection of cells that bound VLPs. Increasing streptavidin:VP1 ratios of 0,01 to 0,1 gave proportional increases in the fluorescence associated with labeled cells, whereas further increases led to a plateau, or indeed reductions, in the observed fluorescence. A streptavidin:VP1 ratio of 0.1 was therefore used for subsequent experiments (Fig 1E). To assess functionality in a relevant biological context, dual-labeled VLPs were tested on PBMC samples from a BKPyV-seropositive KTR. Flow cytometry revealed a distinct population of CD19⁺ lymphocytes that bound both AF647 and PE-labeled BKPyV VLPs, but not AF488-labeled MuPyV VLPs (Fig 1F). Finally, we compared different degrees of biotinylation to maximize detection of antigen-specific B-cells (S2 Fig). PBMC samples from two donors were stained with VLPs carrying either 15–20 or 3–5 biotins per VP1. The latter condition proved superior, yielding approximately twice as many double-positive cells, and increasing the frequency of detected BKPyV-specific B-cells to 2–3% of the CD19⁺ population. These optimized labeling conditions: 3-5 biotins per VP1, and 0,1 streptavidin-oligonucleotide complexes per VP1, were used for single-cell RNA sequencing experiments.

To validate barcode detection, we performed two scRNA-seq experiments using HEK 293TT cells labeled with different antigen combinations. In the first, five different cell samples were each labeled with a single VLP-streptavidin-barcode complex, and tagged in parallel with a hashtag antibody (HTO), then mixed together. After demultiplexing, positive VLP barcode UMI counts were detected on the correct cell population in each case (S3 Fig A), showing that binding of BKPyV VLPs to cells could be detected by the barcode oligonucleotide UMI counts associated with those VLPs. The second experiment combined multiple antigens within a single sample in order to develop an analytical workflow to distinguish antigen-positive from antigen-negative cells under more complex conditions, again using HTO data to define the “actual positives” for each VLP binding combination. For each VLP probe, UMI count thresholds that discriminated between positive and negative populations were defined by mixture modeling (S3 Fig B), then the performance of purely cutoff-based classification of cells positive for single or multiple VLPs was compared to that of Leiden clustering using the whole dataset. With one exception (cluster 6, corresponding to cells labeled with only one barcoded VLP), Leiden clustering increased sensitivity while maintaining high specificity, demonstrating the accuracy and robustness of the integrated pipeline (S3 Fig C).

### 2. Single-cell profiling of BKPyV-specific B cell repertoires in KTR

#### 2.1. Characterization of the KTR cohort

KTR transplanted at the Nantes CHU hospital between 2010 and 2023 were retrospectively selected based on the availability of PBMC samples collected either during a phase of declining BKPyV DNAemia or under conditions of persistent high-level DNAemia. Patients were stratified into two groups: those who controlled viral replication with DNAemia either undetectable or remaining below 3 log UI/mL three months after PBMC sampling (controllers = C), and those with DNAemia remaining above 3 log UI/mL after PBMC sampling (non-controllers = NC). A total of 27 PBMC samples were collected from 23 kidney transplant recipients (KTR), including 18 controllers and 5 non-controllers, with four patients sampled longitudinally at two distinct time points (S1 Table). Prior to single-cell sequencing, the frequency of BKPyV-specific B-cells was assessed by flow cytometry to identify suitable samples for LIBRA-seq. All samples had a detectable population of double-positive BKPyV genotype I VLP–binding B-cells (PE and AF647), ranging from 0.3% to 8% of CD19+ B-cells. However, seven samples with <0.5% specific B-cells and samples with low PBMC recovery were excluded from sequencing, corresponding to six controller samples and one non-controller sample. Fig 2B shows the percentage of BKPyV VLP–specific B-cells among CD19⁺ B-cells, with open symbols indicating samples that were not selected for single-cell sequencing. PBMC samples from controller patients exhibited a significantly higher frequency of BKPyV VLP–specific B-cells compared with those from non-controller patients (p<0.05).

**Fig 2.**
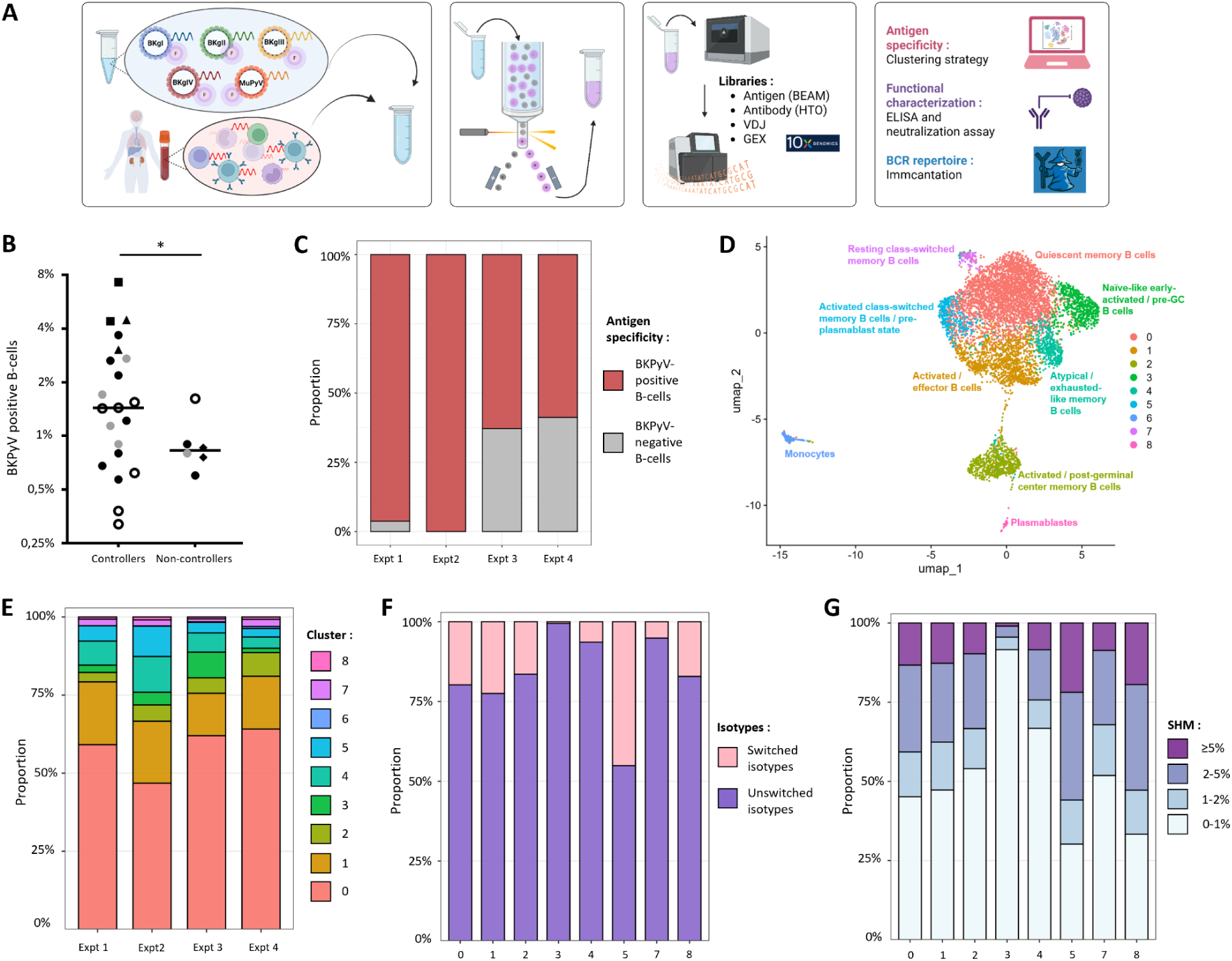
LIBRA-seq workflow and single-cell characterization of BKPyV-specific B-cells. (A) Experiment workflow of the LIBRA-seq assay. Dual-labeled BKPyV and MuPyV VLPs, carrying both fluorescent tags and DNA barcodes, were incubated with PBMC samples, each uniquely labeled with hashtag oligonucleotides (HTOs). Antigen-specific B-cells were subsequently sorted and processed for single-cell sequencing using the 10x Genomics platform. Antigen capture (BEAM), VDJ, gene expression (GEX), and cell surface (HTO) libraries were generated and sequenced. Bioinformatic analyses included antigen specificity assignment using clustering strategies and BCR repertoire analysis using the Immcantation framework. Functional characterization was performed through antibody identification and production, followed by ELISA and neutralization assays. (B) Percentage of BKPyV-specific cells in PBMC samples from controller and non-controller patients. Open symbols represent samples not sequenced by scRNA-seq, grey symbols represent samples with fewer than 50 cells after bioinformatic processing, and black symbols represent samples with more than 50 cells that were included in downstream analyses. Circles indicate a single PBMC sample per patient. Triangles, squares, and diamonds each represent two PBMC samples collected at different time points from the same patient, with each symbol shape corresponding to a distinct patient. Statistical significance was assessed using a Lognormal Welch’s t-test: * p < 0,05. (D) Distribution of transcriptionally defined B-cell clusters across the four single-cell experiments. Clusters were characterized based on their dominant differential gene expression signatures. (E) Distribution of switched (IgG and IgA) and unswitched (IgM and IgD) isotypes across B-cell clusters. (F) Distribution of SHM frequencies across B-cell clusters, stratified into four levels: 0–1% (unmutated), 1–2% (lowly mutated), 2–5% (moderately mutated), and ≥5% (highly mutated).

Clinical and biological characteristics reported hereafter correspond exclusively to the sequenced samples (20 PBMC samples from 17 KTR) (Table 1). The median age was 63 years and 55 for C and NC patients respectively. The median time post-transplant at sampling was 9,9 months for C patients and 13,0 months for NC patients. Graphical representations of BKPyV DNAemia over time, the timing of PBMC sampling, and longitudinal adjustments of immunosuppressive treatments are provided in S4 Fig.

**Table 1.**
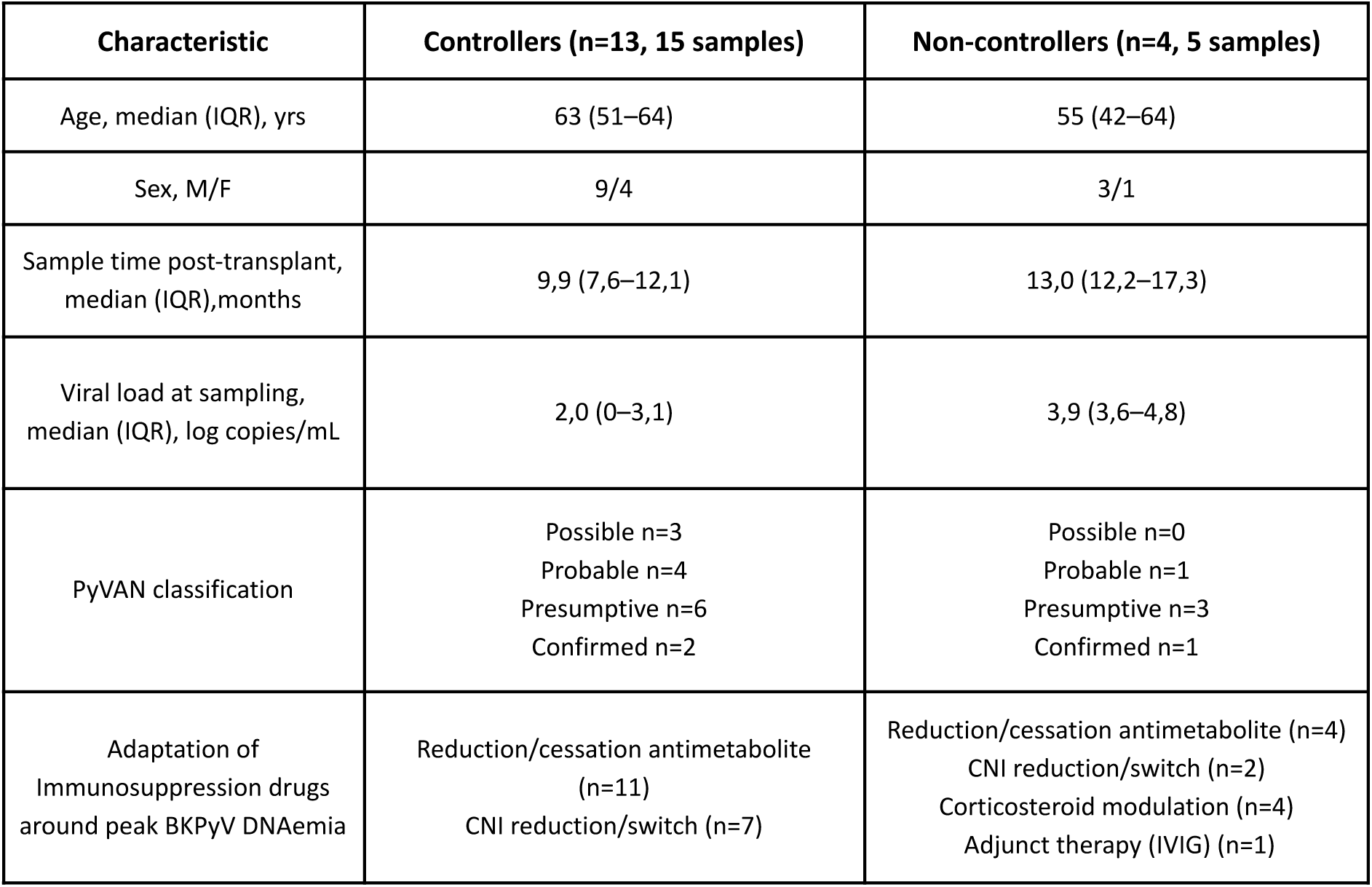
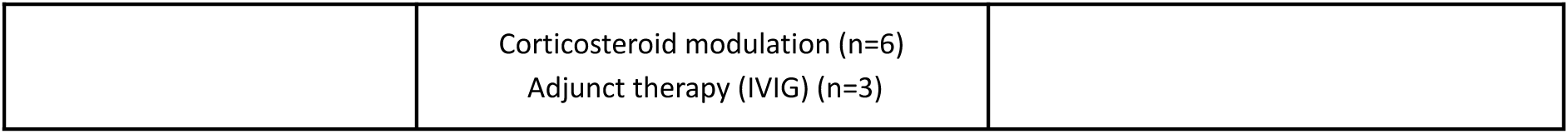
Clinical and biological characteristics of the study cohort.

| Characteristic | Controllers (n=13, 15 samples) | Non-controllers (n=4, 5 samples) |
| --- | --- | --- |
| Age, median (IQR), yrs | 63 (51–64) | 55 (42–64) |
| Sex, M/F | 9/4 | 3/1 |
| Sample time post-transplant, median (IQR), months | 9,9 (7,6–12,1) | 13,0 (12,2–17,3) |
| Viral load at sampling, median (IQR), log copies/mL | 2,0 (0–3,1) | 3,9 (3,6–4,8) |
| PyVAN classification | Possible n=3<br>Probable n=4<br>Presumptive n=6<br>Confirmed n=2 | Possible n=0<br>Probable n=1<br>Presumptive n=3<br>Confirmed n=1 |
| Adaptation of Immunosuppression drugs around peak BKPyV DNAemia | Reduction/cessation antimetabolite (n=11)<br>CNI reduction/switch (n=7) | Reduction/cessation antimetabolite (n=4)<br>CNI reduction/switch (n=2)<br>Corticosteroid modulation (n=4)<br>Adjunct therapy (IVIg) (n=1) |
|  | Corticosteroid modulation (n=6)<br>Adjunct therapy (IVIG) (n=3) |  |

Thirteen patients were infected with BKPyV genotype Ib2, two patients with genotype Ia (2012-5 and 2012-21), one patient with genotype II (2021–07), and one patient with genotype IV (2010–6). Patients were further classified according to the most recent Kotton et al., 2024 criteria for BKPyV-associated nephropathy (PyVAN possible/probable/presumptive/confirmed) (11). In group C, 62% (8/13) patients were classified as presumptive or confirmed PyVAN, against 75% (3/4) patients for the NC group.

#### 2.2. Single-cell sequencing

To investigate the BKPyV-specific B-cell repertoire, B-cells from KTR were incubated with dual-labeled VLPs representing each BKPyV genotype, along with MuPyV VLPs as a negative control (Fig 2A). BKPyV-specific B-cells (CD3^-^ CD16^-^ CD14^-^ CD19⁺ BKPyV-VLP⁺) were then sorted and processed using the 10x Chromium platform for scRNA-seq. Four scRNA-seq experiments in total were performed, including n=4, n=5, n=5 and n=6 PBMC samples respectively (S2 Table). Different sorting conditions were applied: for experiments 1 and 2, the purity mode was selected, whereas the yield mode was used for the last two experiments. Overall, approximately 7000 to 10000 sorted cells were loaded onto the 10X Genomics scRNA-seq platform, depending on the experiment. The raw sequencing data yielded 1350, 5750, 3950, and 3302 cells for experiments 1 to 4, respectively. After quality control and filtering, 40–48% of the cells per experiment were retained for downstream analyses. In the first two experiments, analysis of filtered data revealed that the vast majority of recovered cells bound to at least one BKPyV genotype (*Beam_code* ≠ 0), with only 2% of non-specific B-cells (*Beam_code* = 0). In contrast, the proportion of non-specific B-cells increased to 39% in the last two experiments, which was attributable to differences in the cell-sorting modes applied during the wet-lab procedures (Fig 2C).

To assess the consistency of cellular profiles across experiments and characterize global B-cell heterogeneity, we performed an integrated analysis of all four datasets. B-cell populations were defined based on transcriptional clustering and the expression of differentially expressed genes (Fig 2D). Clusters were characterized according to their dominant molecular signatures, allowing the identification of naïve-like early activated B cells (cluster 3), activated/effector B cells (cluster 1), quiescent memory B cells (cluster 0), activated memory B cells (cluster 2), activated class-switched memory B cells (cluster 5), resting class-switched memory B cells (cluster 7), atypical memory B cells (cluster 4), and plasmablasts (cluster 8). A minor contaminating population of CD14⁺ monocytes was also identified (cluster 6). Examining the data at the sample level revealed no major differences in cellular composition between experiments, although clusters 4 and 5 were slightly more represented in experiment 2 (Fig 2E). When integrating VDJ data with the transcriptionally defined B-cell clusters (Fig 2E–F), cluster 3, corresponding to naïve-like B cells, was almost exclusively composed of unswitched sequences (>99%) and showed very limited somatic hypermutation (SHM), with >90% of cells carrying less than 1% SHM. In contrast, cluster 5, corresponding to activated class-switched memory B cells, showed the highest proportion of class-switched isotypes among the clusters (45%) and the highest level of SHM, with 70% of cells carrying more than 1% SHM.

#### 2.3. Identification of cross-reactive BKPyV-specific B cells

To visualize the LIBRA-seq data, UMI counts for each antigen barcode were first log10 transformed, then background log10 UMI counts for the MuPyV probe were subtracted from the log10 UMI counts of each BKPyV VLP probe. Cells were then projected onto a UMAP with data points colored by log10 UMI counts for each VLP, with increasing red intensity indicating higher positivity for a given genotype. Figs 3A and 3B illustrate this analysis for experiment 2. In parallel, positivity thresholds were defined for each probe, assuming a bimodal distribution. Cells identified as positive for a given genotype showed strong concordance between the PE and AF647 probes and Leiden clustering grouped cells according to their antigen-specific signatures, enabling the assignment of a *Beam_code* to each cluster, including a cluster (*Beam_code = 0*) representing cells that did not bind any BKPyV VLP probe (Fig 3A–B). Cross-reactive clusters are defined as those positive for more than one genotype. In this experiment, we observed several combinations: B-cells binding VLP probes for genotypes 1+2 (*Beam_code = 12*), genotypes 1+3 (*Beam_code = 13*), genotypes 1+4 (*Beam_code =* 14), genotypes 1+2+4 (*Beam_code = 124*), and genotypes 1+2+3+4 (*Beam_code = 1234*). This last cluster indicates cells positive for all four genotypes and reflecting potential cross-reactivity. Notably, *Beam_code = 1234* cells were identified in all experiments, although their relative frequencies varied from 3,1% in experiment 1 to 19,6% in experiment 4 (Fig 3C).

**Fig 3.**
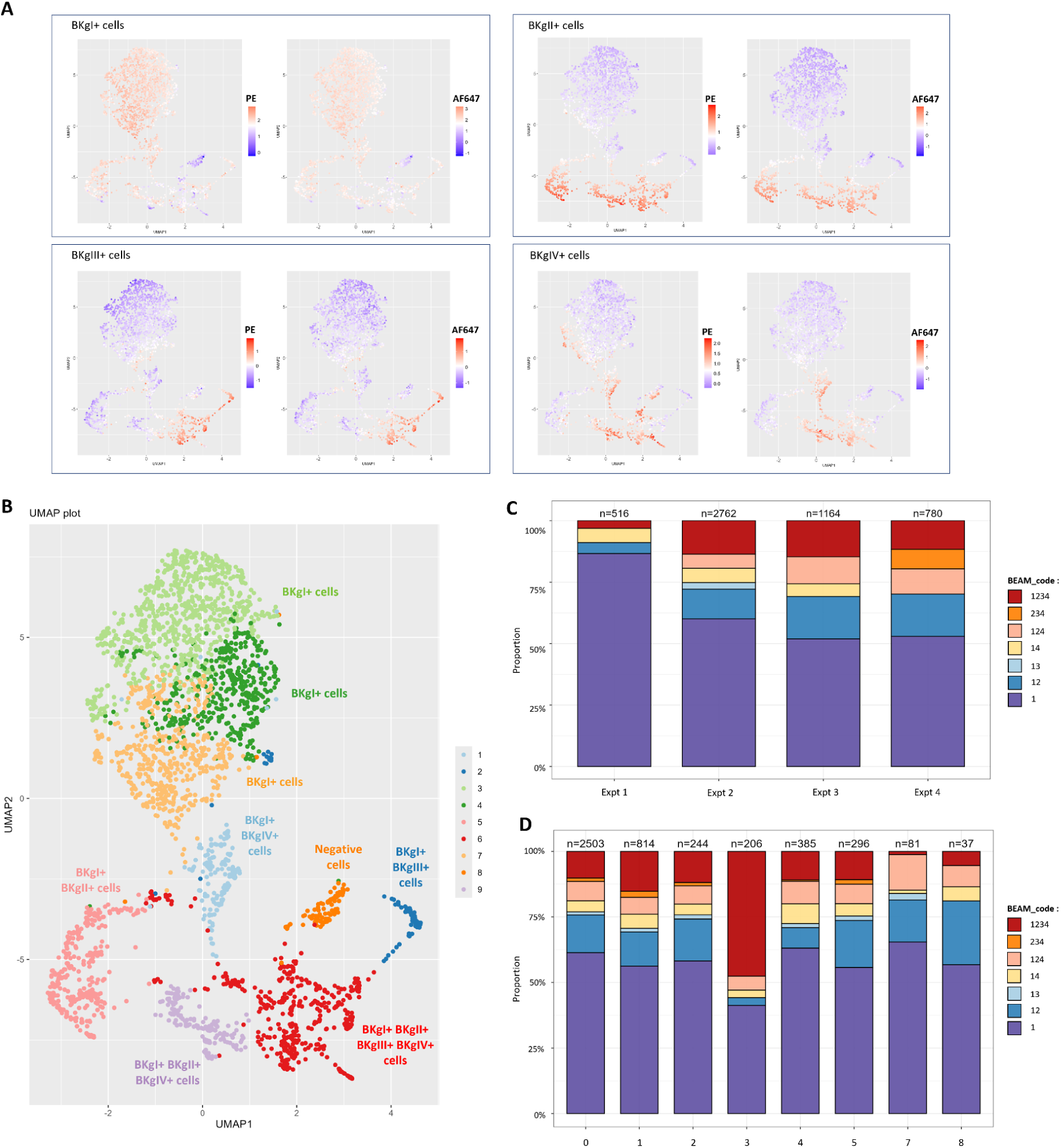
Identification of antigen specificity of BKPyV-specific B-cells using thresholding and clustering strategies. (A) UMAP projection of B-cells from experiment 2 based on UMI counts for each genotype and fluorescent probe, revealing distinct clusters. (B) Leiden clustering identified nine B-cell clusters in experiment 2, with each cluster assigned according to positivity thresholds for the respective probes. Distribution of antigen specificity (=*Beam_code*) across the four experiments (C) and across B-cell clusters (D). *Beam_code = 1* corresponds to B-cells positives for BKPyV genotype 1, *Beam_code = 12* corresponds to B-cells positives for both BKPyV genotypes 1 and 2, and so on.

The internal consistency of antigen specificity clusters was evaluated by inspecting the *Beam_code* of individual B-cells within expanded clonotypes. In general, all cells of a given clonotype were associated with the same *Beam_code* (S5 Fig), as would be expected if the *Beam_code* reflected the epitope bound by a particular antibody. The only exception was *Beam_code* 13, which was interspersed with *Beam_code* 1 in the rare clonotypes when this antigen specificity cluster was found. Some clonotypes showed *Beam_code* distributions suggestive of the acquisition of broader specificity together with SHM, with cells closer to the root of the tree associated with more restricted *Beam_codes* (clonotypes 746, 1697, 3233, 4509, 4956, S5 Fig), but this was not a consistent observation.

In terms of B-cell differentiation, cross-reactive and BKgI-restricted cells were observed across multiple transcriptionally defined clusters (Fig 3D). Interestingly, the naïve-like cluster 3 showed the highest proportion of *Beam_code = 1234* cells (47%), suggesting that the germline BCR repertoire encodes multiple clones with the potential to bind all four BKPyV genotypes, and indeed several expanded clonotypes, with all cells in the clonotype classed as *Beam_code = 1234*, had BCRs close to germline (e.g. 3569, 3498, 4623, S5 Fig).

Thirty-eight antibodies (7 IgM, 1 IgA, and 30 IgG) from B-cells with different *Beam_code* annotations were selected for expression in order to test their antigen-specificity directly. Cells with antigen UMI counts that were either fully consistent (antigen UMI counts of the cell above or below the threshold that defined the cluster annotation for all BK genotypes), or partially consistent with their Leiden cluster were chosen in order to determine whether the *Beam_code* annotation would still reflect the BK genotype specificity of the antibody, even in cases where the individual cell’s antigen UMI counts were ambiguous. Of 7 IgM antibodies, only mAb_4370 could be expressed and purified as an IgG1 monoclonal. This antibody did not show specific binding to BKPyV VLPs in ELISA, but did nevertheless display weak neutralization of BKPyV BKgI and BKgII (Fig 4). In contrast, 24 of the 31 IgG or IgA antibodies were successfully expressed, all of which showed binding to at least one BKPyV genotype. Among the antibodies from cells with antigen UMI counts fully consistent with their *Beam_code*, there was good qualitative concordance between the antibody’s specificity in binding and neutralization assays, and its *Beam_code*, with the sole exception of the *Beam_code = 13* antibody. On the other hand, antibodies from cells with ambiguous antigen UMI counts had less consistent patterns of antigen-specificity, with antibody binding less broad than predicted by the *Beam_code* cluster in some cases (mAb_1790, mAb_4221), and broader in others (mAb_4258, mAb_4509, mAb_318).

**Fig 4.**
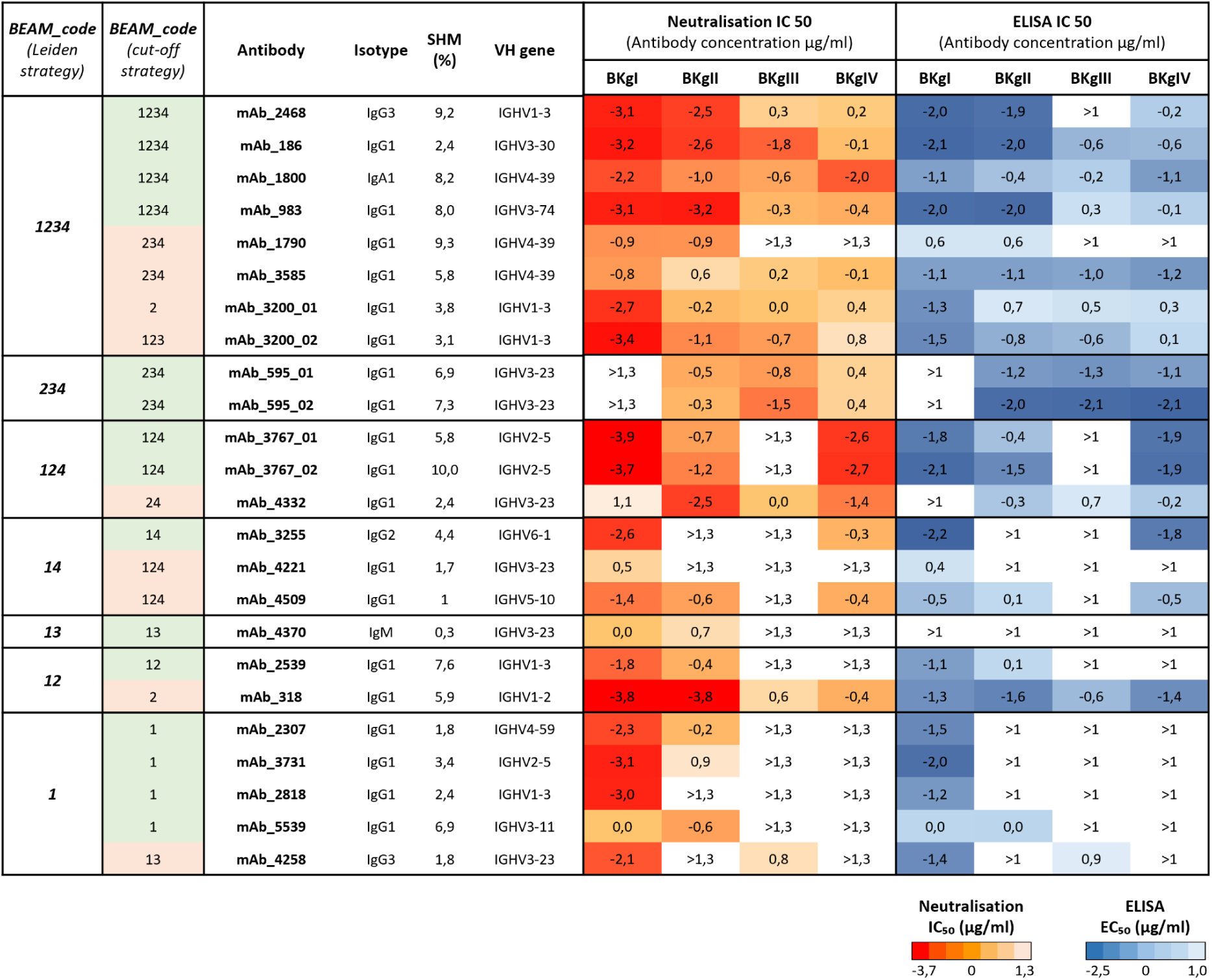
Functional characterization of selected BKPyV-specific monoclonal antibodies. (A) Summary of binding and neutralization properties of the expressed monoclonal antibodies. For each antibody, isotype, SHM frequency, and VH gene usage are indicated. Neutralization of BKPyV genotypes I–IV PSV was assessed first and expressed as half-maximal inhibitory concentration (IC₅₀), followed by binding to BKPyV genotypes I–IV VLP measured by ELISA and expressed as half-maximal effective concentration (EC₅₀). IC₅₀ and EC₅₀ values are displayed as heatmaps, ranging from high potency (dark red for IC₅₀ and dark blue for EC₅₀) to low potency (light red and light blue, respectively). Non-neutralizing and non-binding antibodies are shown in white.

When taking into account antibody binding EC50 and neutralizing IC50, it was evident that most antibodies in the *Beam_code = 1234* category did not display full cross-reactivity; that is, they showed 10 to >100-fold better binding and neutralization of BKgI BKPyV compared to BKgIII and BKgIV. Inspection of UMI counts for different BKPyV genotype VLPs in the *Beam_code = 1234* group showed that lower counts were observed for gIII and gIV probes compared to gI and gII probes (Fig 5A), suggesting that relative UMI counts for the different VLP probes binding to B-cells could reflect the relative affinities of the BCR for different BKPyV genotypes. Subclustering of the *Beam_code = 1234* population based on the delta of log10-transformed gI-gII, gI-gIII and gI-gIV UMI counts allowed us to identify a group of 60 B-cells with highly similar UMI counts for all four BKPyV genotype probes (Cluster 3, Fig 5B–C). Six IgG antibodies from this sub-cluster were selected, of which five were successfully expressed and purified. Two of these antibodies gave only weak binding when tested at 5 µg/ml, while the three others showed equivalent binding to all four BKPyV genotypes by ELISA (Fig 5D). Two antibodies also showed potent neutralisation of all four BKPyV genotypes, with IC50 values close to those observed for the previously described antibody mAb120. Subcluster 3 of the *Beam_code = 1234* population was therefore enriched in B-cells expressing antibodies with equivalent binding to all four BKPyV genotypes, and in subsequent analyses, the *Beam_code = 1234* population was split into *Beam_code = 12_34* (subclusters 1 and 2), and *Beam_code = 1234\** (subcluster 3).

**Fig 5.**
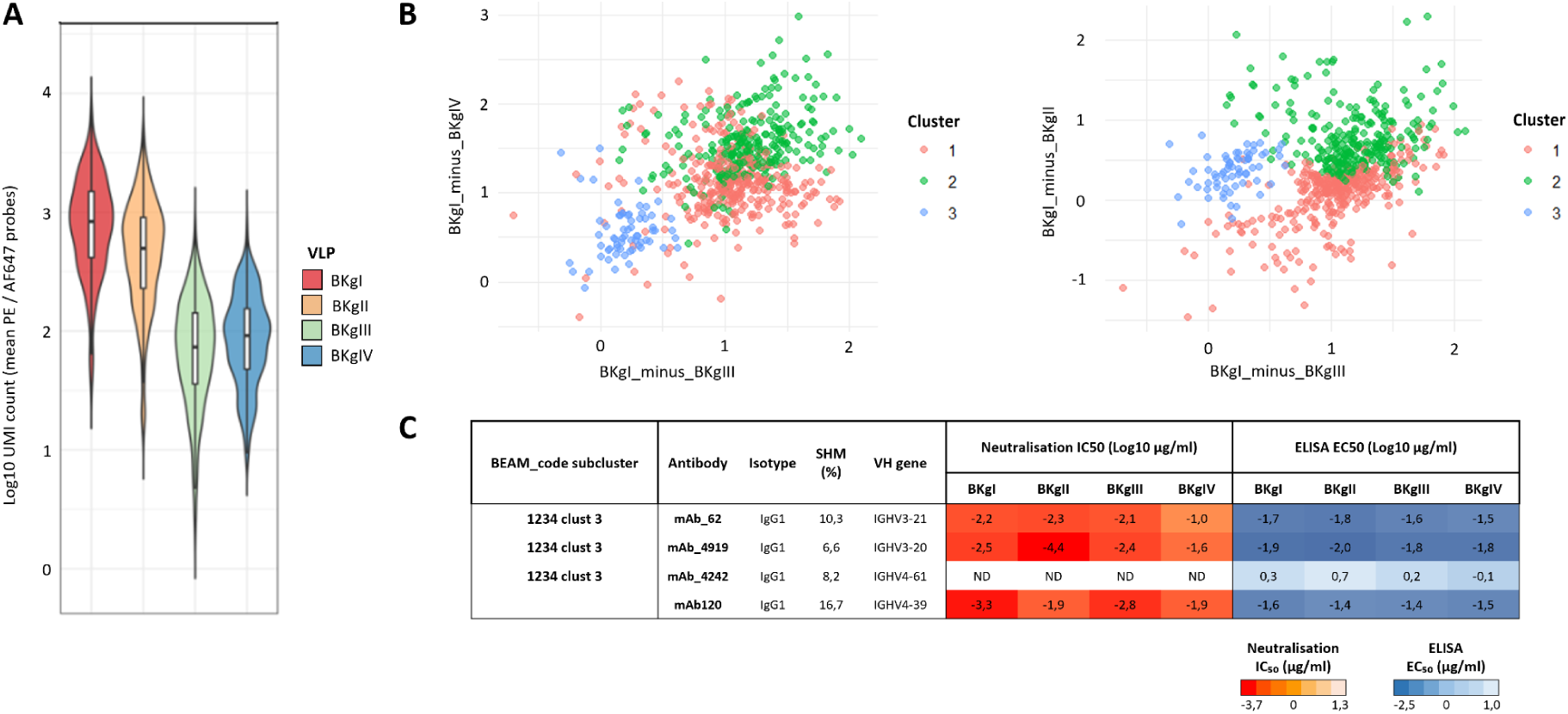
Identification of B-cell cluster enriched for broadly BKPyV-specific antibodies. (A) Log10 UMI counts for different BKPyV genotype VLP probes within the *BEAM_code = 1234* cluster. (B) Log10 UMI counts for gIII and gIV VLP probes to cells in the *BEAM_code = 1234* cluster, relative to gI VLP probe. (C) Log10 UMI counts for gIII and gII VLP probes to cells in the *BEAM_code = 1234* cluster, relative to gI VLP probe. (D) Summary of binding and neutralization properties of three monoclonal antibodies from subcluster 3. *ND - Not determined*

#### 2.4. Integrating repertoire analysis with antigen specificity

To characterize the diversity of paired BCR repertoires from BKPyV-infected kidney transplant patients at the single-cell level, we analyzed their VDJ sequences, including isotype usage, IGHV/IGKV/IGLV gene assignments, SHM levels, and clonotype composition using the Immcantation framework, then combined BCR repertoire data with the antigen specificity data. Cells with *Beam_code* = 0 and samples with fewer than 50 BKPyV-specific cells were excluded from these analyses. Most BKPyV-reactive cells belonged to non-switched isotypes, predominantly IgM (81%) with a minor proportion of IgD (1%), whereas switched isotypes accounted for 16% IgG and 2% IgA overall. Notably, *Beam_code = 1234\**, *12* and *234* contained a higher fraction of class-switched cells (37%, 37% and 42% respectively) compared with the other *Beam_codes*, which ranged between 12% and 22% (Fig 6A). When restricting the comparison to genotype-specific (*Beam_code = 1*) versus cross-reactive cells (cells with *Beam_code* > 1), cross-reactive cells displayed a significantly higher proportion of class-switched isotypes (26%) than BKgI-specific cells (12%) (Chi-square test, p-value < 0,001) (Fig 6B). SHM levels were significantly higher in cross-reactive cells (Wilcoxon rank-sum test, p-value < 0,001), with BCRs from cells in *Beam_code = 124*, *234* and *1234\** categories exhibiting the highest SHM levels when stratified by *Beam_code* and isotype (Fig 6C–D).

**Fig 6.**
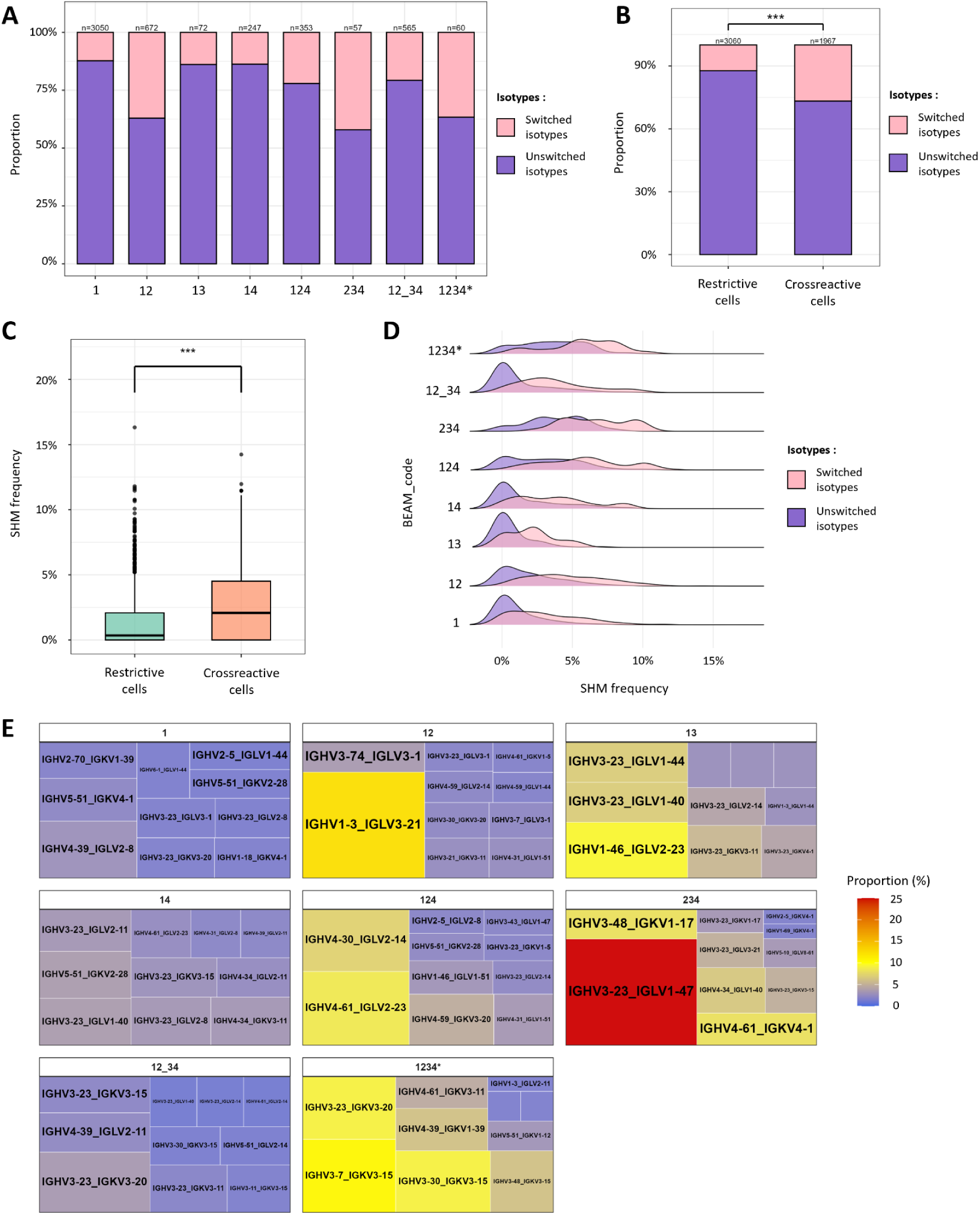
B repertoire according to antigen specificity (*Beam_code*). (A) Distribution of isotype subgroups (switched and unswitched) according to antigen specificity (*Beam_code*). (B) Distribution of isotype subgroups (switched and unswitched) among restrictive B-cells (*Beam_code = 1*) and cross-reactive B-cells (*Beam_code ≠ 1)*. Statistical significance was assessed using a Chi-square test: *** p < 0,001. (C) Distribution of SHM frequencies among restrictive B-cells (*Beam_code = 1*) and cross-reactive B-cells (*Beam_code ≠ 1*). Statistical significance was assessed using a Wilcoxon rank-sum test: *** p < 0,001. (D) Ridge plots showing SHM frequencies according to antigen specificity (*Beam_code*). (E) Treemap showing the usage frequency of VH–JH gene pairs according to antigen specificity (*Beam_code*). Each rectangle represents a unique VH–JH gene combination, with rectangle size and color indicating the relative frequency of each gene pair.

Regarding the most frequent paired heavy and light chain clonotypes, we observed distinct VH–VL combinations depending on the *Beam_code* (Fig. 6E). In the specificity groups *Beam_code* = *1*, *14* and *12_34*, the repertoire was diverse, with clearly over-represented VH–VL pairing observed In contrast, certain combinations showed marked enrichment, with more than 10% of cells expressing IGHV1-3/IGLV3-21 for *Beam_code = 12* (12,4%) and IGHV3-23/IGLV1-47 for *Beam_code = 234* (29%). A moderate enrichment was also observed for the categories *Beam_code = 124*, *13* and *1234\** where one or more VH-VL pairing made up 5-10% of the repertoire. Interestingly, of the top five VH-VL pairings in the *Beam_code = 1234\** category, only two (IGHV3-23/IGKV3-20 and IGHV3-30/IGKV3-15) were also strongly represented in the *Beam_code = 12_34* group, with the other three pairings being specifically over-represented in *Beam_code = 1234\**. These findings indicate that while most BKPyV-specific responses mobilize diverse VH–VL configurations, the genotype combinations 1+2 and 2+3+4 may impose stringent structural constraints, leading to preferential selection of particular VH–VL pairs.

#### 2.5. BKPyV-specific B-cell repertoire in relation to suppression of virus replication

We next sought to characterize the B-cell repertoire using the full set of data, with the aim of analyzing isotype usage, SHM, and clonotype diversity across patients, and to relate these findings to antigen specificity. Our primary objective was to compare the B-cell repertoire between two patient groups: controllers, whose BKPyV-DNAemia became undetectable or declined to low levels after the peak, and non-controllers, whose BKPyV-DNAemia remained persistently high. Our initial hypothesis was that controllers would display a broader BKPyV-specific B-cell repertoire, whereas non-controllers would exhibit a more restricted response. More than 50 BKPyV-specific B-cells were obtained for 11 patients, therefore, only these individuals were included in subsequent analyses.

When examining genotype-specific proportions (=*Beam_code*) at the patient level, both broad and restricted reactivity profiles were observed in controller and non-controller patients (Fig 7A). Some patients displayed broadly cross-reactive repertoires, such as patient 2013-7, in whom *Beam_code = 12_34* and *124* B-cells accounted for >28% of the repertoire, while only 31% of B-cells were specific for genotype 1. In contrast, other patients exhibited highly restricted cross-reactivity profiles, with 80% to 83% of B-cells reactive to genotype 1 alone (2012-21, 2023-01 and 2023-07). The only patient with BKPyV gII infection (2021–07) had a noticeably distinct *Beam_code* distribution. This individual showed the lowest proportion of cells positive exclusively for genotype I (12%), with cells predominantly positive for the combined genotypes I+II (29%), and genotypes II+III+IV (24%). The presence of genotype I monospecific B-cells in this patient was surprising, and we therefore examined the VP1 protein sequence in the patient’s virus, which was found to differ from the gII VLP antigen probe that was used in LIBRA-seq. Specifically, amino acids 61 and 62 were N61D62 in the gII VLP probe, but D61N62 in the patient’s virus and E61N62 in the gI VLP probe. This may explain the presence of a *Beam_code* = 1 population in this patient, and suggests a potential epitope involving these residues. B-cells with some ability to bind all four genotypes (*Beam_code = 12_34* or *1234\**) were detected in every patient, irrespective of clinical group, and the frequency of these B-cells was not reduced in non-controllers. The more restricted *Beam_code = 1234\** group was not observed in all patients, but was nevertheless present in one of three non-controllers.

**Fig 7.**
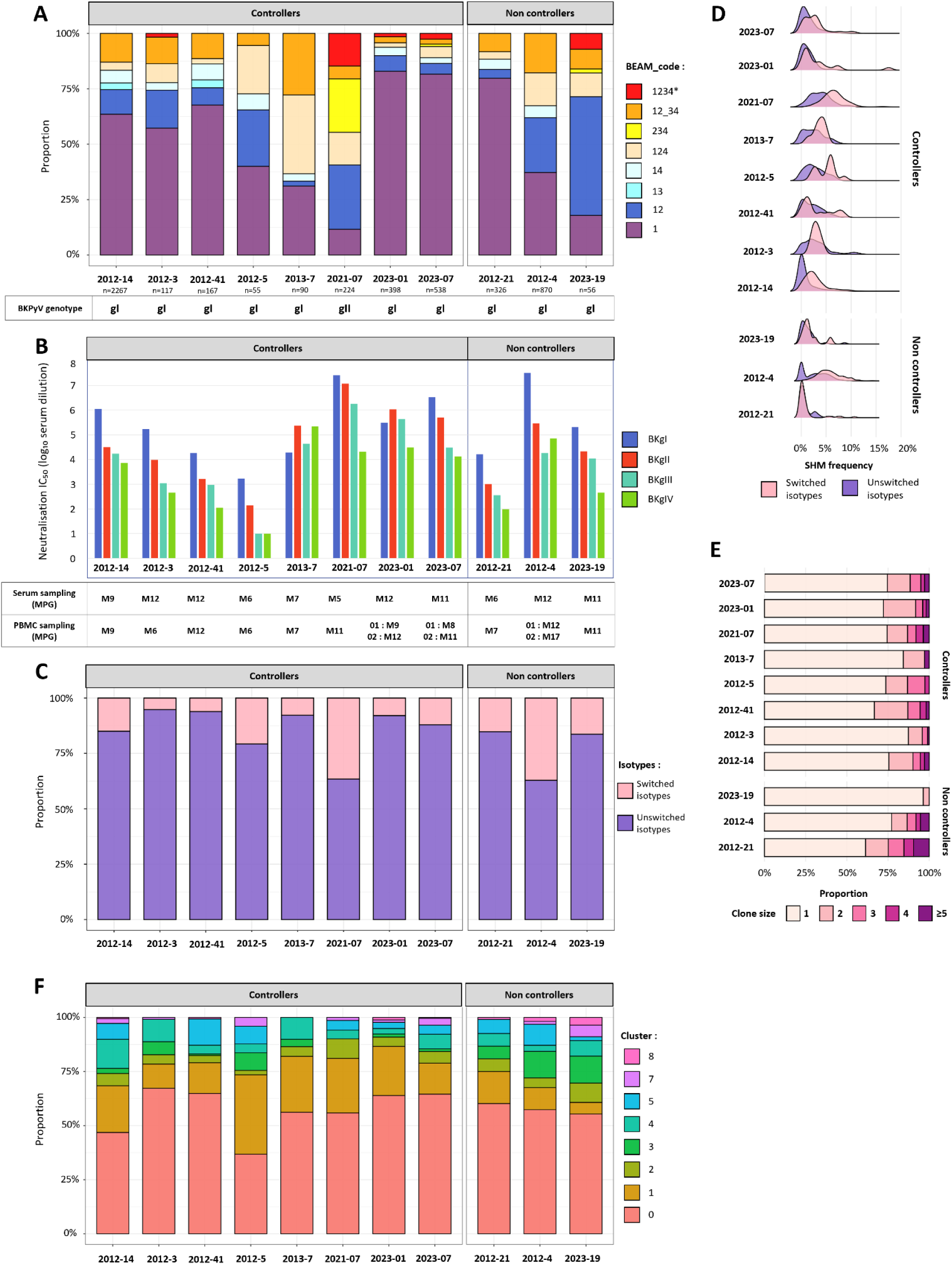
Antigen specificity and B-cell repertoire features in controllers and non-controllers. (A) Distribution of antigen specificity (=*Beam_code*) across patient samples, grouped as controllers or non-controllers. The infecting BKPyV genotype is indicated below. (B) Neutralisation IC50 values (log10 serum dilution) for each patient against the four BKPyV genotypes. Serum and PBMC sampling time points used for neutralisation and scRNA-seq experiments, respectively, are shown below as months post-graft. (C) Distribution of isotype groups (switched and unswitched) across patient samples, grouped as controllers or non-controllers. (D) Ridge plots showing SHM frequencies for switched and unswitched isotypes in each controller and non-controller patients. (E) Distribution of clone size proportions, ranging from single-cell clones to clones containing five or more cells, across controller and non-controller patients. (F) Distribution of B-cell sub-populations across controller and non-controller patients.

Neutralizing titers against the four BKPyV genotypes were assessed in parallel using serum samples from each patient (Fig 7B). For most patients, serum samples were collected at the same time point as the PBMC samples used for scRNA-seq analyses. Two exceptions were due to limited serum availability: for patient 2012-3, the serum sample was collected more than six months after the PBMC sampling, and for patient 2021-07, the serum sample was collected six months prior to PBMC sampling. Overall, strong neutralizing titers were observed against BKPyV genotypes I and II, whereas weaker neutralization was detected for genotypes III and IV in the majority of patients, with no obvious differences in neutralizing titer or breadth between controllers and non-controllers.

The proportion of IgA/IgG class-switched B-cells and SHM levels were similar between patients with persistent DNAemia and those who controlled BKPyV replication (Fig 7C–D), although two of the three non-controllers (2023-19 and 2012-21) had very low levels of SHM in class-switched isotypes. Overall, clonal expansions were limited, with a mean of 76,7% of clonotypes containing only a single cell across patients. However, some patients exhibited moderate expansion, such as patient 2012-21, in whom clones of size ≥5 accounted for up to 9,5% of the repertoire (Fig 7E). The distribution of B-cell clusters appeared to differ slightly between controllers and non-controllers (Fig 7F). Cluster 1, corresponding to immediate-early activated B cells, accounted for 11–36% of B cells in controllers, compared with 5–10% in non-controllers. In contrast, cluster 3, characterized by a naïve-like, early activated phenotype, represented 0,7–8% of B cells in controllers and 6–12% in non-controllers.

Taken together, these findings indicate that the B-cell repertoire does not differ substantially between controllers and non-controllers, thereby not supporting our initial hypothesis that non-controllers would exhibit a more restricted repertoire. However, despite the overall similarity between patient groups, we noted considerable heterogeneity in the PBMC samples, which we hypothesized might be related to the interval between PBMC sampling and the peak of DNAemia. In order to explore this hypothesis, we next analyzed the data at the sample level by stratifying specimens according to the time elapsed since the DNAemia peak. Three groups were defined: samples collected <3 months, 3–6 months, or >6 months after the peak.

To avoid over-representation of patients with higher cell numbers, all analyses were performed after downsampling to *n* = 55 cells per timepoint category, followed by 100 bootstrap resamplings. Using this normalized dataset, we observed that within the first three months following the peak of DNAemia, the antigen-specific B-cell repertoire was highly restricted, with a predominance of genotype 1–specific cells, representing 73% of the response. As the time from the viremic peak increased, regardless of whether BKPyV-DNAemia was controlled or not, a progressive broadening of the repertoire was observed. The proportion of cross-reactive cells was 59% at 3–6 months and 48% beyond 6 months post-peak (Fig 8A), with the proportion of *BEAM_code = 1234\** cells increasing progressively over time. Isotype switching and antibody maturation also increased with time post peak DNAemia class-switched cells making up 26% of BKPyV-specific B-cells after 6, compared with 10% and 13% at <3 months and 3–6 months, respectively (Fig 8B), and the proportion of BKPyV-specific B-cells exhibiting a mutation rate greater than 5% rising from 8% at <3 months to 13% at 3–6 months, and reaching 23% beyond 6 months post-peak (Fig 8C). The same tendencies were observed longitudinally in three patients where sequential samples had been analyzed (S5 Fig), however, this progressive maturation and broadening of the response was not accompanied by changes in B-cell transcriptional profiles, as the relative proportions of different B-cell subtypes remained stable over time (Fig 8D).

**Fig 8.**
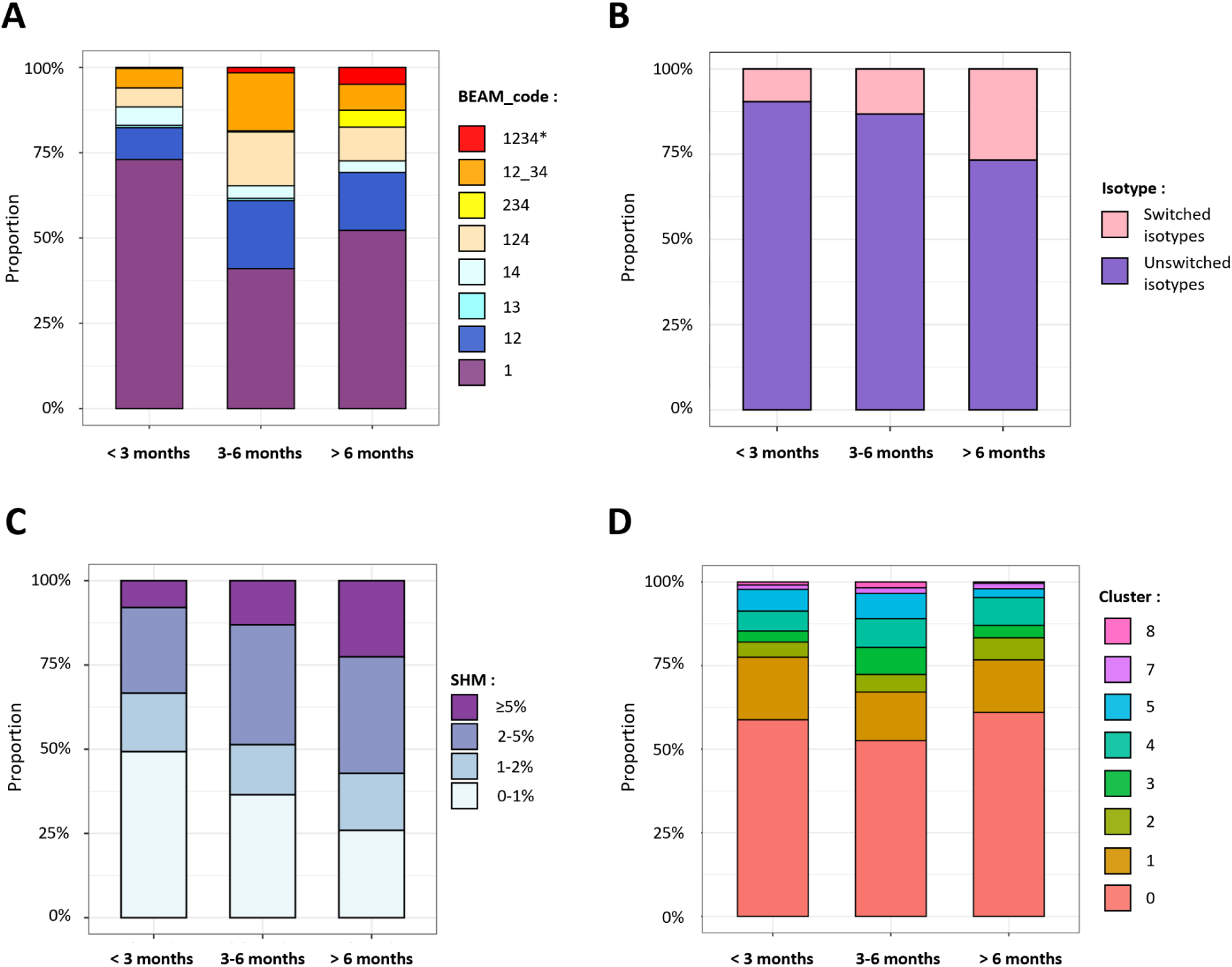
Longitudinal and time-dependent analysis of B-cell repertoire following BKPyV-DNAemia. Distribution of antigen specificity (*Beam_code*) (A), isotype subgroups (switched and unswitched) (B), SHM frequencies (C) and B-cell sub-clusters (D) across three PBMC sampling time points relative to the BKPyV-DNAemia peak (<3 months, 3–6 months, or >6 months after the peak).

## Discussion

The main objective of this study was to analyze the anti-BKPyV humoral response at the single-cell level in KTR who suppressed viral replication compared to those who did not, in order to test the hypothesis that the humoral response in KTR with persistent high-level DNAemia might have a more restricted BKPyV-specific B-cell repertoire, thus making these patients more susceptible to the emergence of neutralization escape mutations in the virus capsid (24,25). In order to address this question, we adapted the LIBRA-seq technique (6) to BKPyV, using virus-like particles as antigen probes. We also developed a new approach for the analysis of LIBRA-seq data by applying Leiden clustering to the UMI counts for the different BKPyV antigen probes, then annotating each cluster. This allowed us to predict the genotype specificity, summarized by the “*BEAM_code*” parameter, for all of the 5197 BKPyV-specific B-cells in the dataset. The validity of the predicted genotype specificity was tested in 24 antibodies chosen from different *BEAM_code* categories, and while some divergence was seen between predicted and experimentally verified antibody properties, observed binding specificities were generally consistent with predicted genotype specificity, with antibodies showing increasing binding and neutralizing breadth across BKPyV genotypes as the *Beam_code* parameter increased from 1 to 1234. This supports the overall validity of our experimental procedure and bioinformatic pipeline, and allows us to draw conclusions concerning the functional breadth of the BKPyV-specific B-cell repertoire between patients, based on our LIBRA-seq data.

In our cohort, all but one of the patients for whom we generated LIBRA-seq data had BKgI BKPyV DNAemia, and the most abundant *BEAM_code* category in the dataset corresponded to BKgI monospecific B-cells. The BKPyV-specific B-cell repertoire was therefore predominantly directed against the infecting genotype, but it also contained a substantial proportion of cross-reactive B cells, which had a higher level of SHM and class-switching than mono-specific B-cells (Fig 6B–C). Cross-reactive B-cells were detected in all patients in the cohort (Fig 7A), although there was considerable heterogeneity in both the proportion and the nature (ie. predominant *BEAM_code* categories) of cross-reactive B-cells between patients.

When comparing controllers and non-controllers, we found no major differences in the composition of the BKPyV-specific B-cell repertoire (Fig 7). The proportions of broadly reactive B cells were comparable between groups, as were the proportion of class-switched B-cells, and clonal analysis did not reveal major oligoclonal expansions. Although a few clones comprising more than five cells were detected, they consistently represented less than 10% of the repertoire, indicating that both controllers and non-controllers were able to generate a diverse and polyclonal BKPyV-specific response. The only significant difference we observed was the lower proportion of BKPyV-specific B-cells in circulating CD19^+^ B-cells in non-controllers (Fig 2B) which, taken together with the low level of SHM in class-switched BKPyV-specific B-cells in two of the three non-controllers, may reflect a lack of T-cell help in these patients. This is consistent with naïve-like B-cells making up a somewhat higher proportion of the BKPyV-specific B-cell population in non-controllers (Fig 7F).

In contrast, the maturation of the humoral response was strongly associated with the time elapsed since peak DNAemia. Class switching, SHM accumulation, and recognition of multiple BKPyV genotypes all increased with time after peak DNAemia in cross-sectional analyses and in the three patients with longitudinal samples. This pattern indicates progressive recruitment and maturation of BKPyV-specific B-cell clones following viral replication. Notably, all patients underwent reduction of immunosuppressive therapy to manage BKPyV replication, suggesting that this intervention was sufficient to allow a classical antiviral B-cell response to develop, even in patients who failed to suppress viral replication.

A major advantage of LIBRA-seq is that antigen-specific UMI counts can be directly linked to individual BCR sequences, providing a bridge between antigen-binding phenotype and antibody sequence, and we sought to leverage this property to identify broadly neutralizing antibody candidates in our dataset. Significantly, B-cells with *Beam_code = 1234* were found in all patients, and although most of these cells had weak cross-reactivity to BKgIII and BKgIV, further analysis of the LIBRAseq data allowed us to identify the *BEAM_code = 1234\** subcluster, characterized by B-cells that bound all four BKPyV genotype probes equally well. Functional characterisation of antibodies from B-cells in this cluster confirmed that it was enriched for true bNAbs, showing that quantitative, as well as qualitative, features of LIBRA-seq data can be exploited to narrow the search for bNAbs. The *BEAM_code = 1234\** subcluster contained a high proportion of class-switched B-cells and both IgG and IgM memory B-cells in this population had high levels of SHM, similar to those seen in the *BEAM_code = 124* and *BEAM_code = 234* clusters. The BCR repertoire of *BEAM_code = 1234\** cells did not appear to rely on previously described VH-VL pairings. Although three cells within the 4242 clonotype displayed 41F17-like CDR3 motifs in the context of an IGHV4-61/IGKV3-11 pairing, the canonical VH-VL combinations of previously described BKPyV bNAbs (13,15–17) were not enriched in the *1234\** population. Together, these findings suggest that broadly neutralizing activity against BKPyV can arise from multiple independent B-cell lineages that undergo affinity maturation, and that the repertoire available for bNAb isolation may be considerably larger than previously appreciated.

Despite the presence of cross-reactive and broadly neutralizing B cells, we found no evidence that the breadth or diversity of the humoral response was associated with control of established BKPyV DNAemia. This is consistent with previous studies indicating that neutralizing antibodies alone are insufficient to clear BKPyV once substantial viral replication has been established, with cellular immunity likely playing an essential role in viral clearance (21,41–43), and has implications for antibody-based interventions. Clinical studies of IVIG and the monoclonal antibody MAU868/P8D11 have so far provided limited evidence for efficacy in patients with established BKPyV DNAemia (18,44) Our results show that the humoral response generated by at least some KTR with persistent BKPyV DNAemia is not deficient, neither in quantitative (anti-BKPyV serum titres) nor in qualitative (repertoire diversity, maturation, and functional breadth) terms, and it is therefore difficult to see how antibody-based approaches could benefit these patients. Antibody-based strategies may therefore be more effective when used prophylactically, before substantial viral replication is established. This is consistent with observations that low pre-transplant BKPyV antibody titres are associated with an increased risk of post-transplant viral replication (20,21,44,45), and supports the investigation of broadly neutralizing monoclonal antibodies or IVIG in high-risk recipients with low pre-existing neutralizing titres (12).

Several limitations should be considered. First, the small number of non-controllers (n = 3) limits statistical power and may have prevented the detection of subtle differences between groups. Second, LIBRA-seq analyses were restricted to patients with sufficient numbers of BKPyV-specific B cells, introducing a potential selection bias toward patients with more readily detectable humoral responses. Finally, PBMC samples were collected at different time points following peak BKPyV DNAemia, which introduced temporal heterogeneity in the observed levels of class switching, SHM, and cross-reactivity that is difficult to control in the comparison of controller and non-controller groups. Longitudinal studies in larger cohorts will be required to determine how these features of the humoral response relate to viral control and to assess the potential of broadly neutralizing antibodies for BKPyV prevention.

## Data Availability

The authors are in the process of depositing FASTQ files of scRNA-seq data in the European Genome-Phenome Archive (https://ega-archive.org/), and R objects containing read-count matrices plus annotated AIRR files with BCR sequences in Zenodo (https://zenodo.org/). DOIs will be added with updates to the present MedRxiv submission.
Pending completion of this process, all data produced in the present study are available upon reasonable request to the authors

https://doi.org/10.5281/zenodo.18414907

https://doi.org/10.5281/zenodo.18432248

## Acknowledgments and Funding

This work was supported by the Agence Nationale de la Recherche (grant number ANR-22-CE15-0043) and by the Région Pays de la Loire (Trajectoire Nationale 2023).

## Ethics Statement

Patient samples from 2010 to 2012 used in this study were obtained from patients recruited into a prospective observational study approved by the local ethics committee and declared to the French Commission Nationale de l’Informatique et des Libertés (CNIL, n°1600141). Samples collected between 2021 and 2023 were derived from the French DIVAT multicentric prospective cohort of kidney transplant recipients (www.divat.fr, CNIL, n°914184). All patients gave informed consent authorizing the use of archived urine and blood samples for research purposes.

## Data availability statement

The authors are in the process of depositing FASTQ files of scRNA-seq data in the European Genome-Phenome Archive (https://ega-archive.org/), and R objects containing read-count matrices plus annotated AIRR files with BCR sequences in Zenodo (https://zenodo.org/).

HEK 293TT cell setup experiments:

S3 Fig A: https://doi.org/10.5281/zenodo.18414907

S3 Fig B: https://doi.org/10.5281/zenodo.18432248

Patient B-cell LIBRA-seq experiments (Figures 4, 7 and 8); annotated AIRR files https://doi.org/10.5281/zenodo.20067938 (data embargoed until publication)

---

**S1 Fig.**
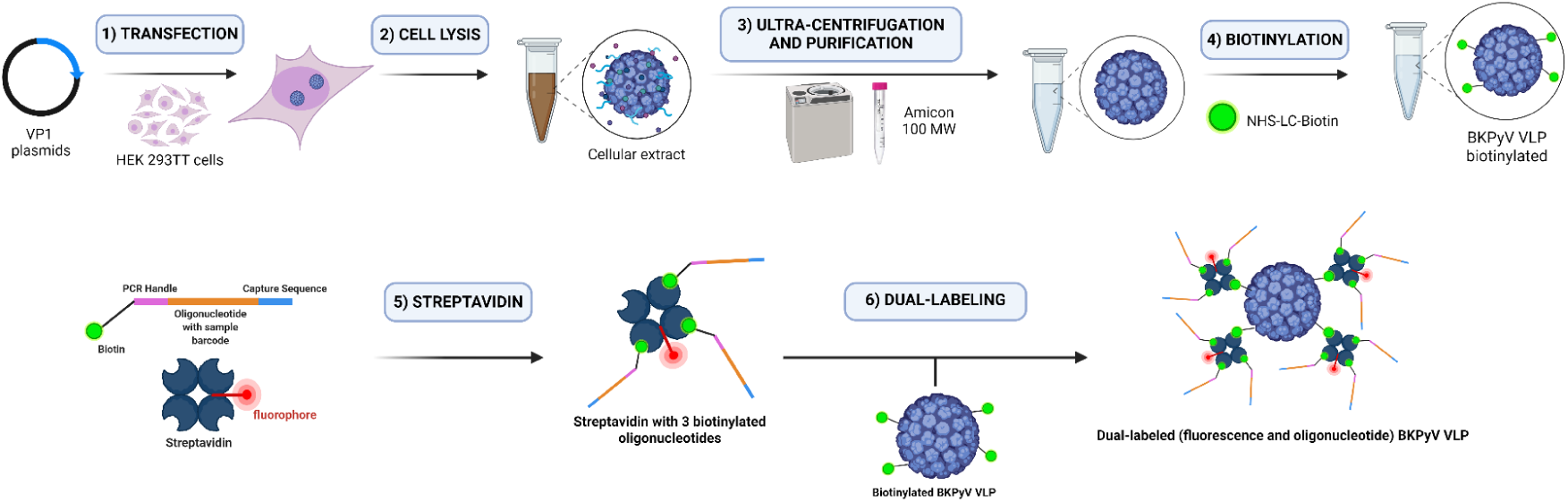
Production of dual fluorescence/oligonucleotide labeled VLPs

**S2 Fig.**
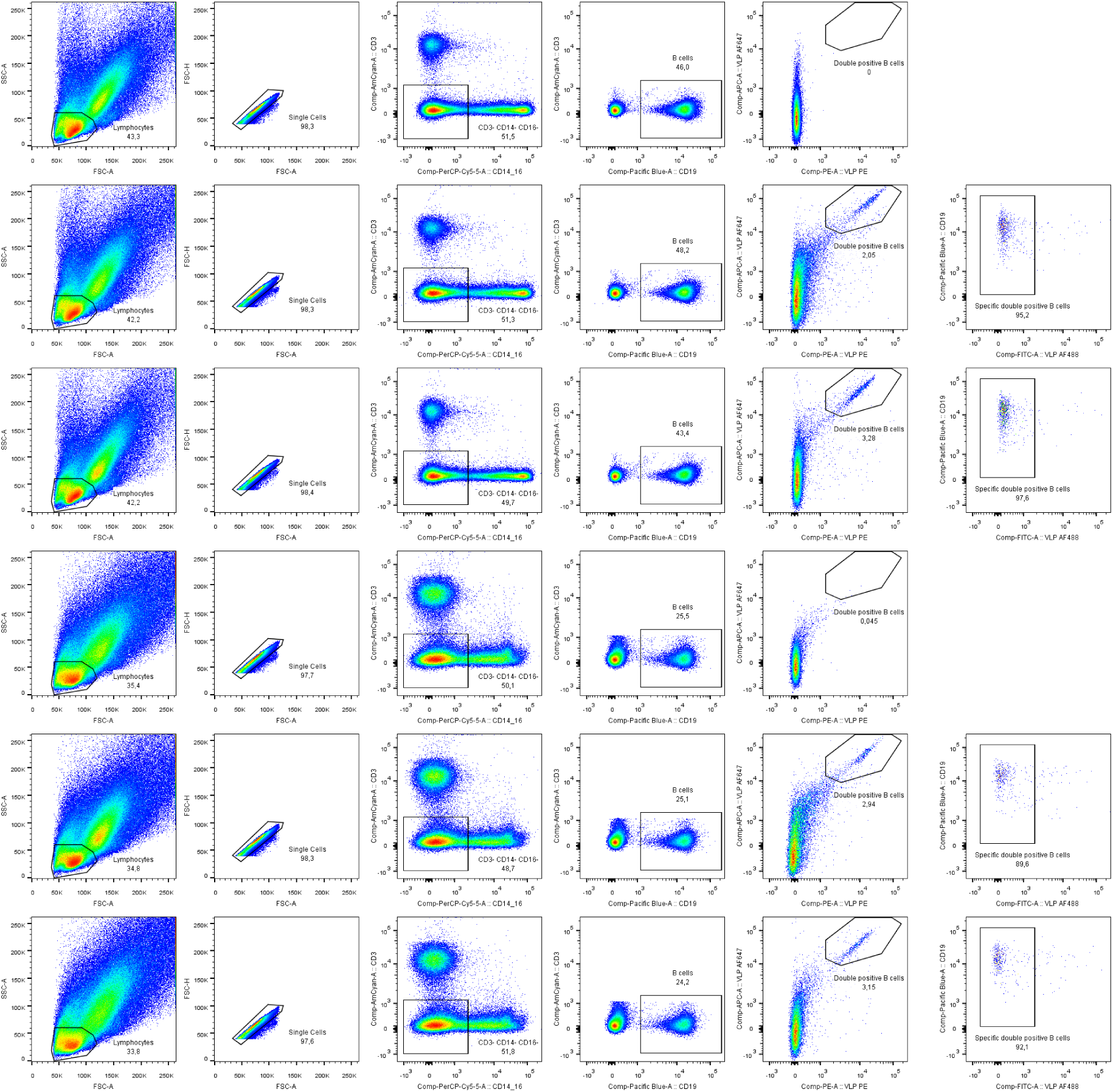
Identification of BKPyV positive B-cells in PBMC samples

**S3 Fig.**
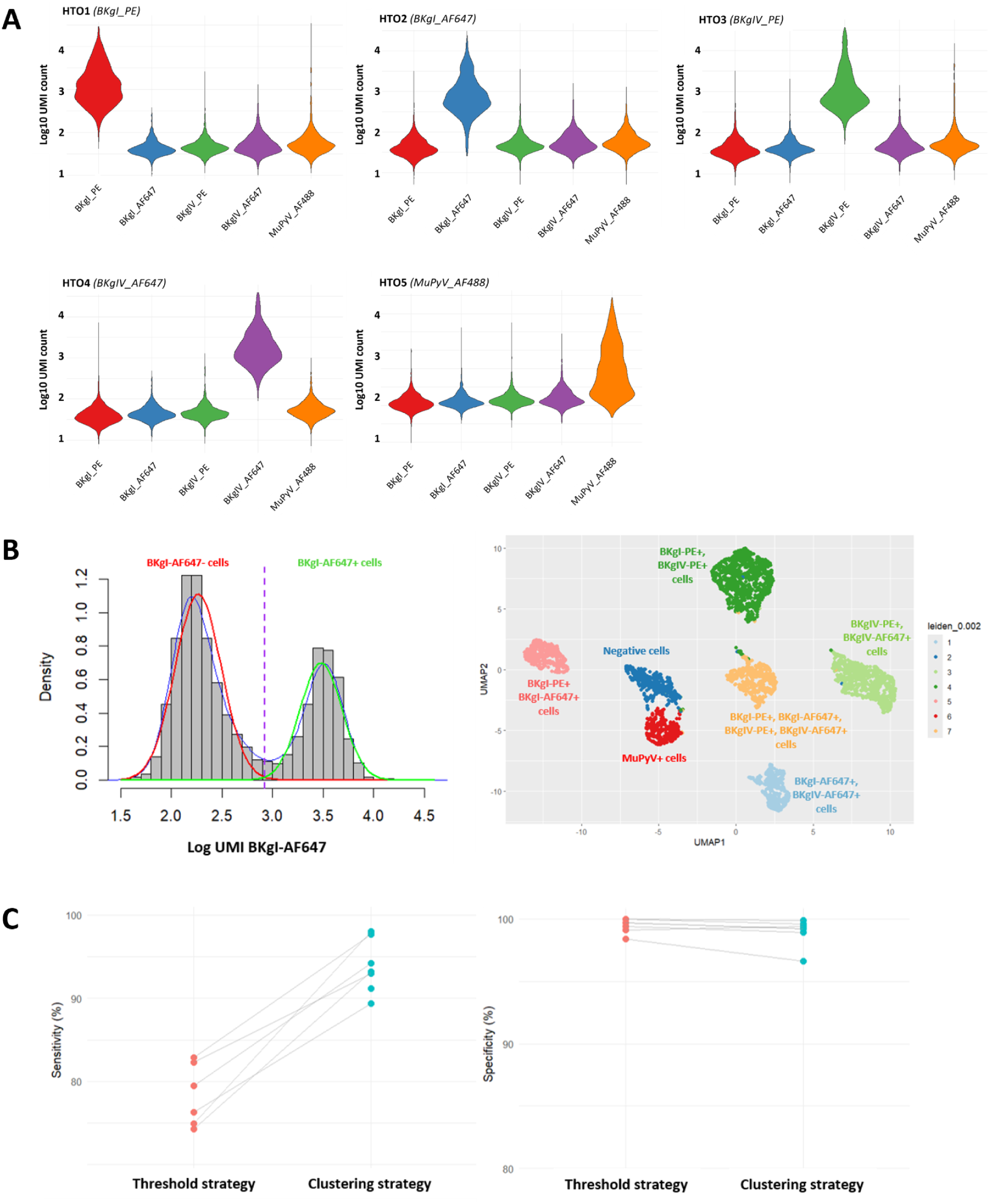
Validation of VLP barcode detection by scRNA-seq. (A) Five separate HEK 293TT cell samples were each labeled with a single polyomavirus VLP–barcode complex (BKPyV I, BKPyV IV, or MuPyV) and a unique hashtag oligonucleotide (HTO), pooled, and processed together using the 10x Genomics Chromium platform, enabling the quantification of probe-specific UMI counts across all five samples. (B) Multiple antigens were combined within a single sample, and two complementary bioinformatic analysis strategies were applied. First, an antigen-specific thresholding approach based on mixture modeling was used to distinguish positive and negative cell populations, illustrated here by BKgI-AF647, for which negative and positive cells were separated using a cutoff of approximately 2,9 log₁₀ UMI counts. In parallel, patterns of VLP binding were visualized by UMAP embeddings, then Leiden clustering was applied to identify seven antigen-defined clusters which were annotated based on the thresholding strategy. (C) Comparison of sensitivity and specificity between UMAP visualization followed by Leiden clustering with threshold-based cluster annotation, and threshold-based classification alone.

**S4 Fig.**
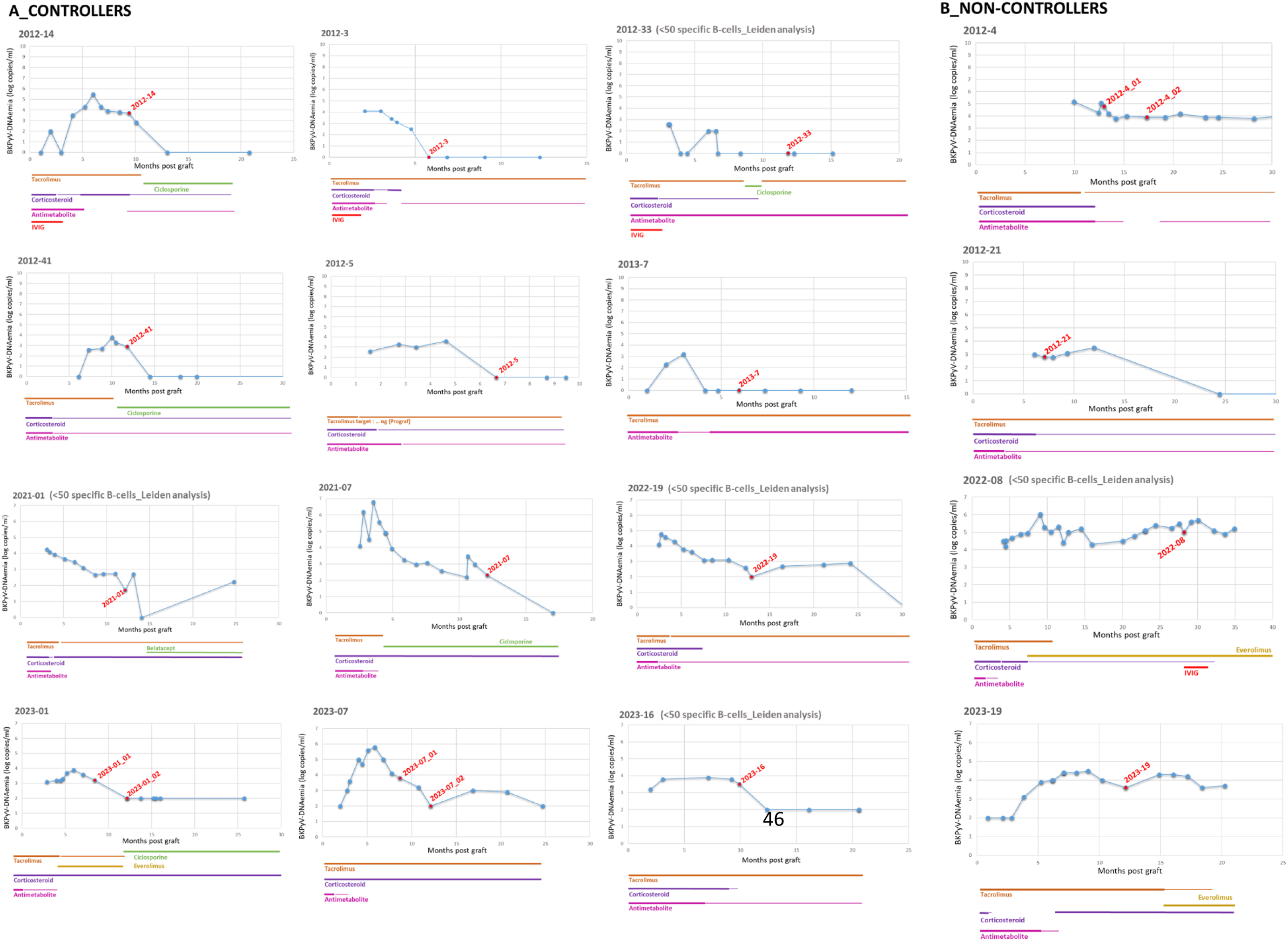
Longitudinal monitoring of BKPyV DNAemia in controllers (A) and non-controllers (B) KTR. Each panel shows BKPyV viral load expressed as log10 copies/mL plotted against time post-transplantation (months) for an individual patient. Colored bars below each graph indicate the intensity of immunosuppressive treatments over time; decreases in bar thickness correspond to dose reductions.

**S6 Fig.**
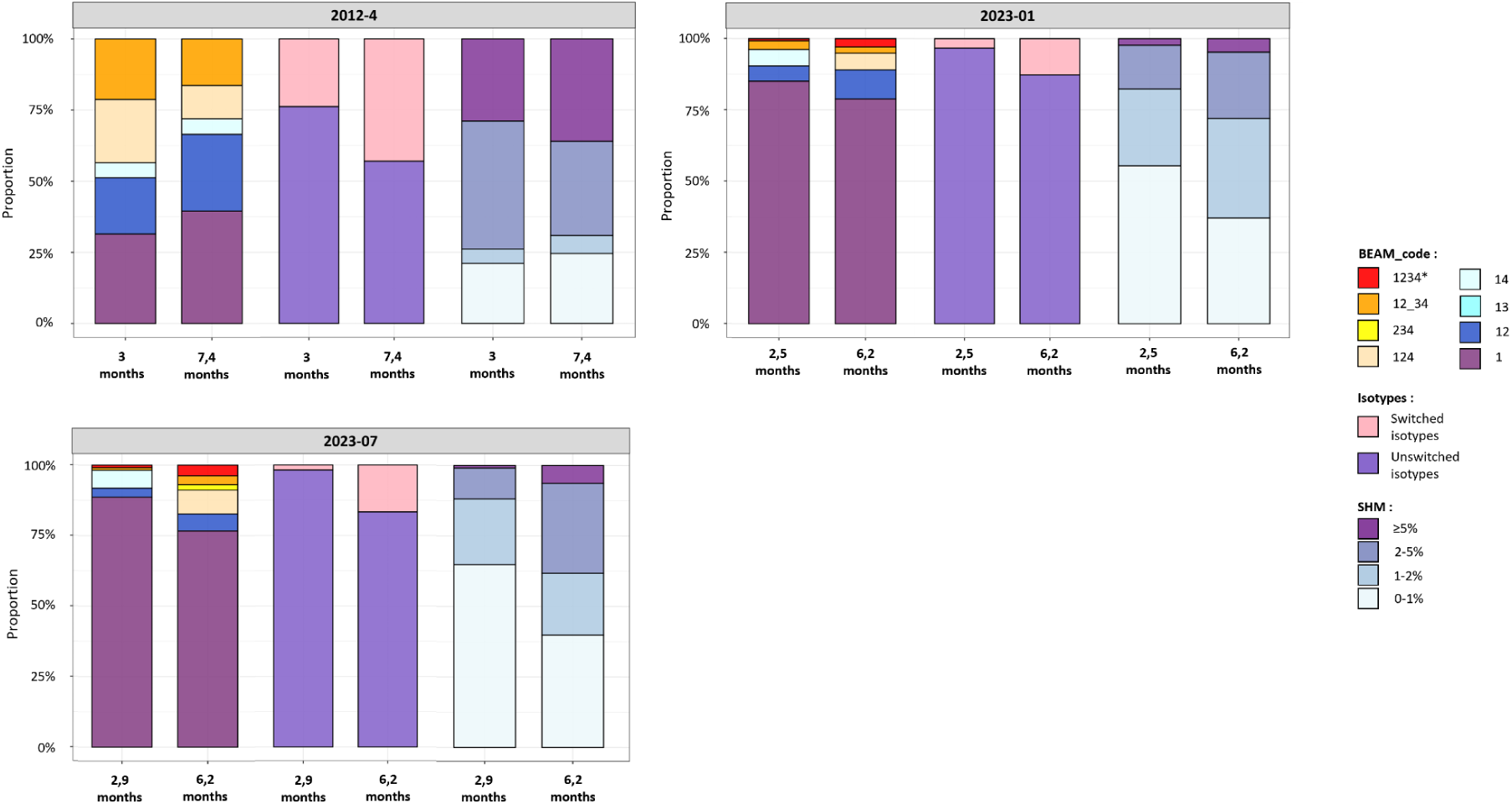
Longitudinal analysis of three patients with two PBMC samples each. Antigen specificity (Beam_code), isotype subgroups, and SHM frequencies are shown for each sample.

**S1 Table.** Summary clinical data. Grey - sample not processed for scRNAseq Salmon - sample processed for scRNAseq, but data for < 50 BKPyV-specific B-cells obtained Green - sample processed for scRNAseq, and data for > 50 BKPyV-specific B-cells obtained

| PBMC sample | Age range (years) | Sex | Transplant number | C/NC | PBMC sampling (months post graft) | PBMC sampling (months post DNAemia peak) | WBC (G/L) | Lymphocytes (G/L) | CD3<sup>+</sup> T cells (G/L) | CD4<sup>+</sup> T cells (G/L) | CD8<sup>+</sup> T cells (G/L) |
| --- | --- | --- | --- | --- | --- | --- | --- | --- | --- | --- | --- |
| 2010-6 | 60-65 | F | 1 | C | 2,8 | no DNAemia | 4,35 | * | 0,692 | 0,518 | 0,15 |
| 2011-07 | 50-55 | F | 1 | C | 5,9 | no DNAemia | 8,46 | 1,1 | 0,633 | 0,265 | 0,351 |
| 2012-06 | 30-35 | M | 1 | C | 6,8 | 4,7 | * | * | 1,203 | 0,563 | 0,615 |
| 2012-14 | 25-30 | F | 2 | C | 9,4 | 3,4 | 10,21 | 2,24 | 1,398 | 0,355 | 0,963 |
| 2012-21 | 45-50 | M | 1 | NC | 7,1 | 1,0 | 9,08 | 0,61 | 1,186 | 0,351 | 0,771 |
| 2012-3 | 40-45 | M | 2 | C | 6,9 | 4,8 | 4,33 | 0,47 | 0,175 | 0,093 | 0,07 |
| 2012-33_01 | 50-55 | M | 1 | C | 4,5 | 1,4 | 9,8 | 0,59 | 0,19 | 0,137 | 0,046 |
| 2012-33_02 |  |  |  |  | 11,9 | 8,8 | 9,02 | 0,26 | 0,061 | 0,043 | 0,019 |
| 2012-4_01 | 25-30 | M | 1 | NC | 13 | 3,0 | 7,55 | 1,32 | 0,699 | 0,188 | 0,478 |
| 2012-4_02 |  |  |  |  | 17,3 | 7,4 | 6,57 | * | * | * | * |
| 2012-41 | 35-40 | M | 1 | C | 11,8 | 1,8 | 8,92 | 1,65 | 1,001 | 0,465 | 0,533 |
| 2012-5 | 60-65 | F | 2 | C | 6,7 | 2,0 | 5,41 | 0,6 | 0,426 | 0,099 | 0,261 |
| 2013-7 | 70-75 | M | 1 | C | 6,0 | 3,0 | 3,8 | 0,83 | 0,654 | 0,536 | 0,118 |
| 2021-01 | 65-70 | M | 1 | C | 12,2 | 9,1 | 8,06 | 1,94 | 1,463 | 0,445 | 0,908 |
| 2021-02 | 25-30 | M | 1 | C | 12,1 | 9,2 | 3,03 | 0,45 | 0,28 | 0,148 | 0,114 |
| 2021-05 | 65-70 | M | 1 | C | 12,6 | 8,6 | 5,97 | 1,47 | 0,968 | 0,522 | 0,434 |
| 2021-07 | 60-65 | M | 1 | C | 12,1 | 8,6 | 12,46 | 1,28 | 0,346 | 0,158 | 0,172 |
| 2022-08 | 60-65 | F | 1 | NC | 28,2 | 19,2 | 5,78 | 1,26 | 1,089 | 0,42 | 0,652 |
| 2022-19 | 50-55 | M | 1 | C | 13,0 | 10,2 | 6,11 | 0,57 | 0,189 | 0,115 | 0,062 |
| 2023-01_01 | 60-65 | M | 1 | C | 8,4 | 2,5 | 8,16 | 0,62 | * | * | * |
| 2023-01_2 |  |  |  |  | 12,2 | 6,2 | 8,58 | 0,37 | 0,166 | 0,111 | 0,049 |
| 2023-07_01 | 65-70 | F | 1 | C | 8,7 | 2,9 | 14,5 | 5 | * | * | * |
| 2023-07_02 |  |  |  |  | 12,1 | 6,2 | 12,28 | 1,42 | 0,568 | 0,24 | 314 |
| 2023-08 | 65-70 | F | 1 | NC | 10,2 | 0,5 | 8 | 0,72 | 0,442 | 0,181 | 0,241 |
| 2023-14 | 75-80 | F | 1 | C | 12,2 | 7,3 | 8,74 | 2 | 1,511 | 0,285 | 1,202 |
| 2023-16 | 60-65 | M | 1 | C | 9,9 | 2,8 | 8,07 | 1,04 | 0,801 | 0,236 | 0,531 |
| 2023-19 | 65-70 | M | 1 | NC | 12,2 | 5,1 | 4,81 | 0,82 | 0,612 | 0,305 | 0,294 |

\*Data not available

**S2 Table.**
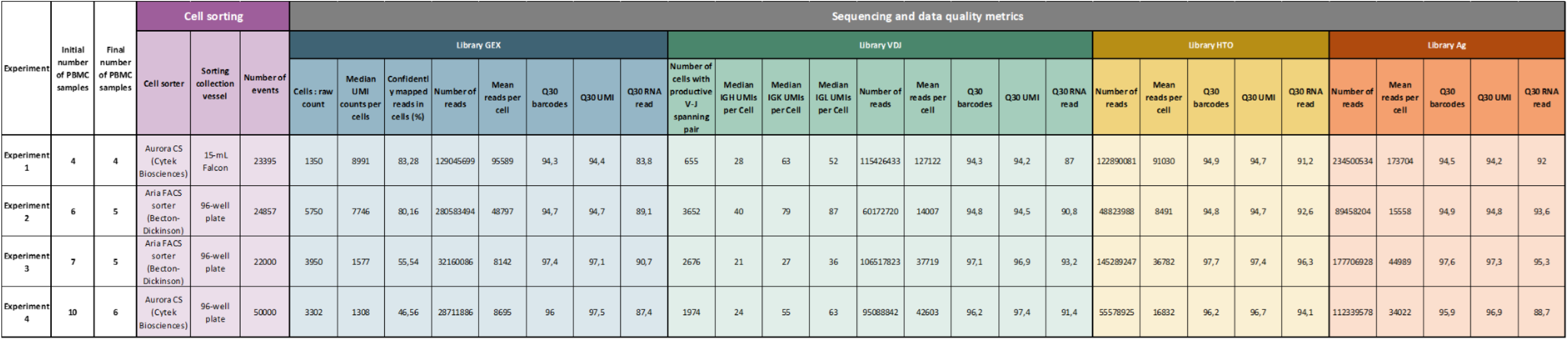
Summary data for yield and QC of cell sorting and scRNAseq runs.

