## Supplemental Figure 5 for "Using LIBRA-seq to map the BK-polyomavirus specific B-cell response in kidney transplant recipients"

5042

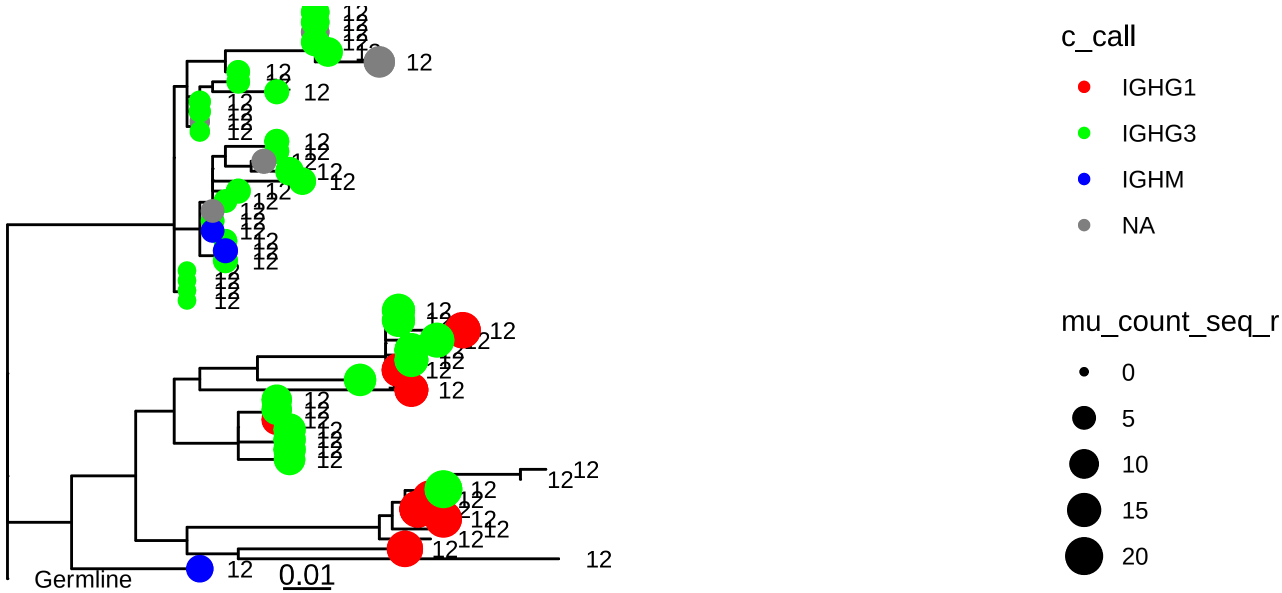


234

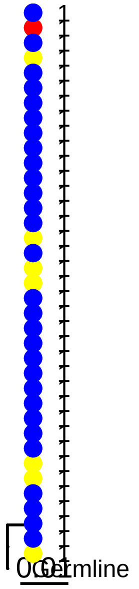


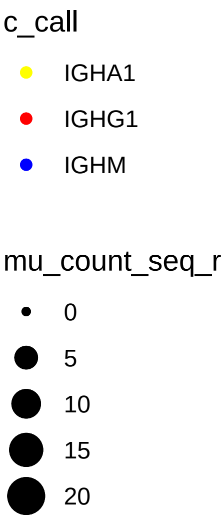


Legend Figure S5:

Clonotype ID is indicated above each phylogram
Tip colour represents antibody isotype
TIp size represents percentage SHM
Tip label represents BEAM_code

2427

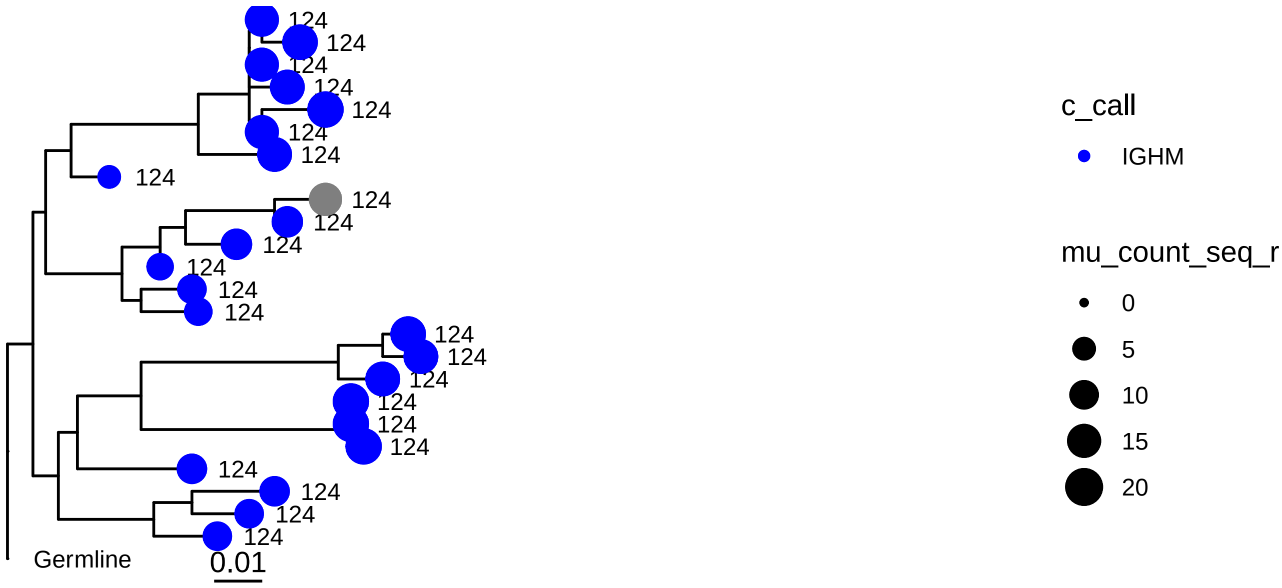
Germline
0.01
c_call
IGHM
mu_count_seq_r
0
5
10
15
20

3597

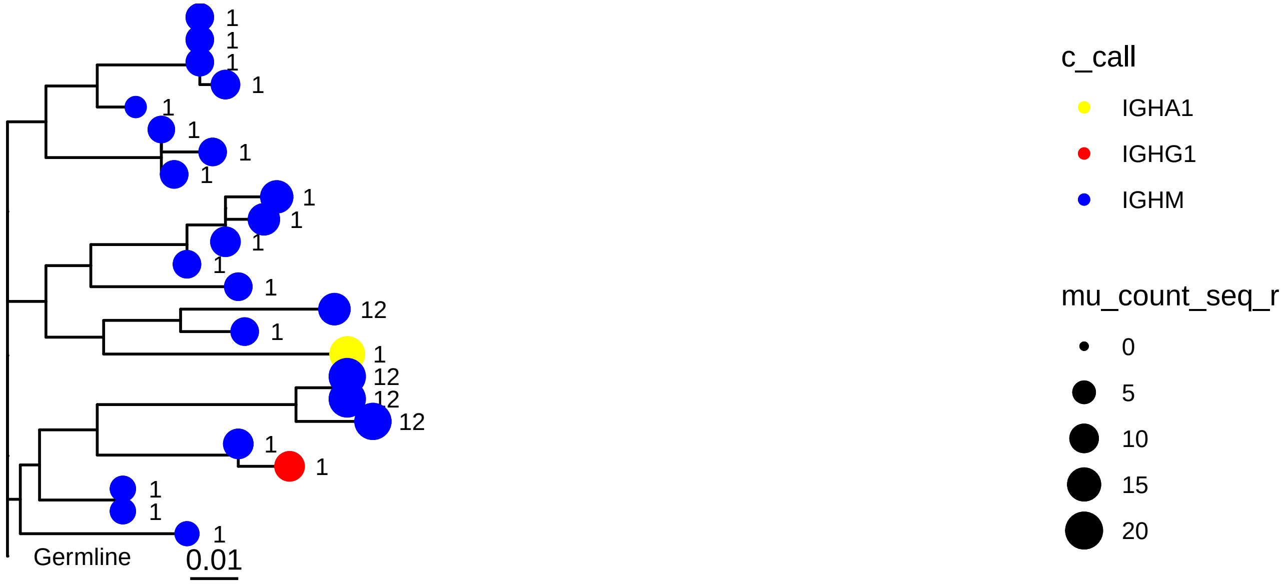
Germline
0.01
c_call
IGHA1
IGHG1
IGHM
mu_count_seq_r
0
5
10
15
20

595

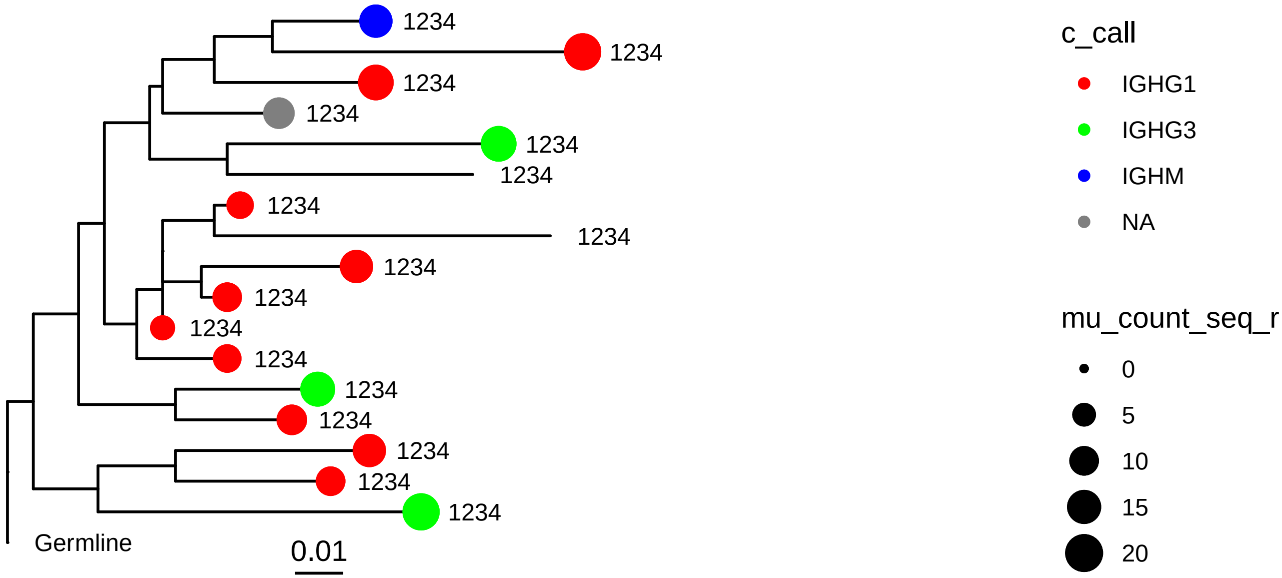
1234
1234
1234
1234
1234
1234
1234
1234
1234
1234
1234
1234
1234
1234
1234
1234
1234
Germline
0.01
c_call
IGHG1
IGHG3
IGHM
NA
mu_count_seq_r
0
5
10
15
20

3195

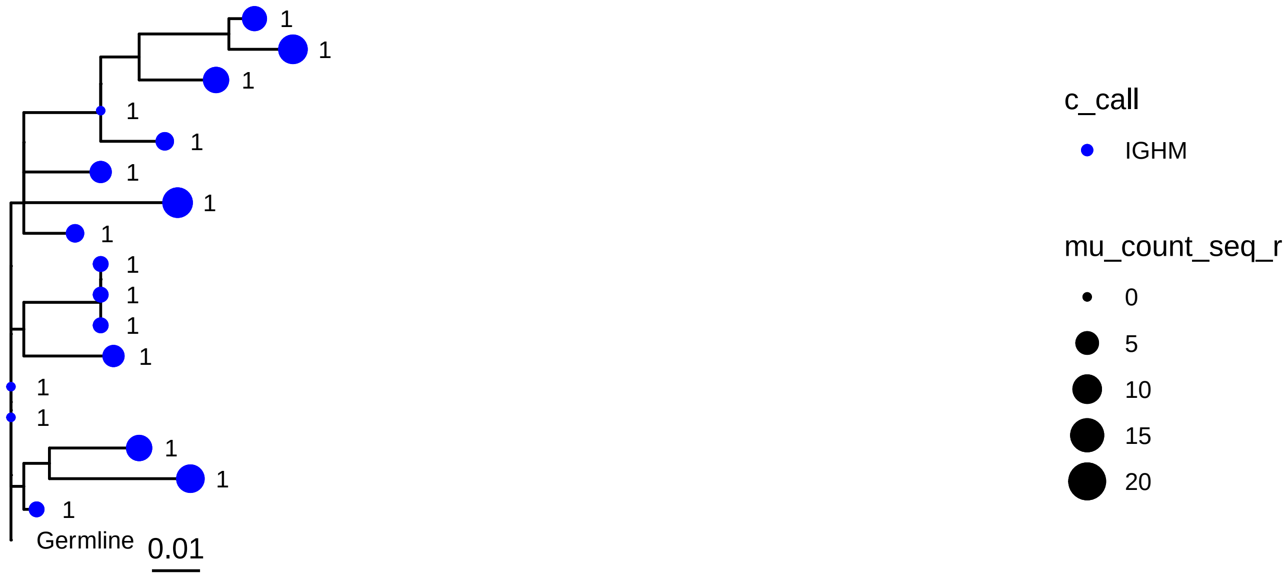
1
1
1
1
1
1
1
1
1
1
1
1
1
1
1
1
1
Germline
0.01
c_call
IGHM
mu_count_seq_r
0
5
10
15
20

4167

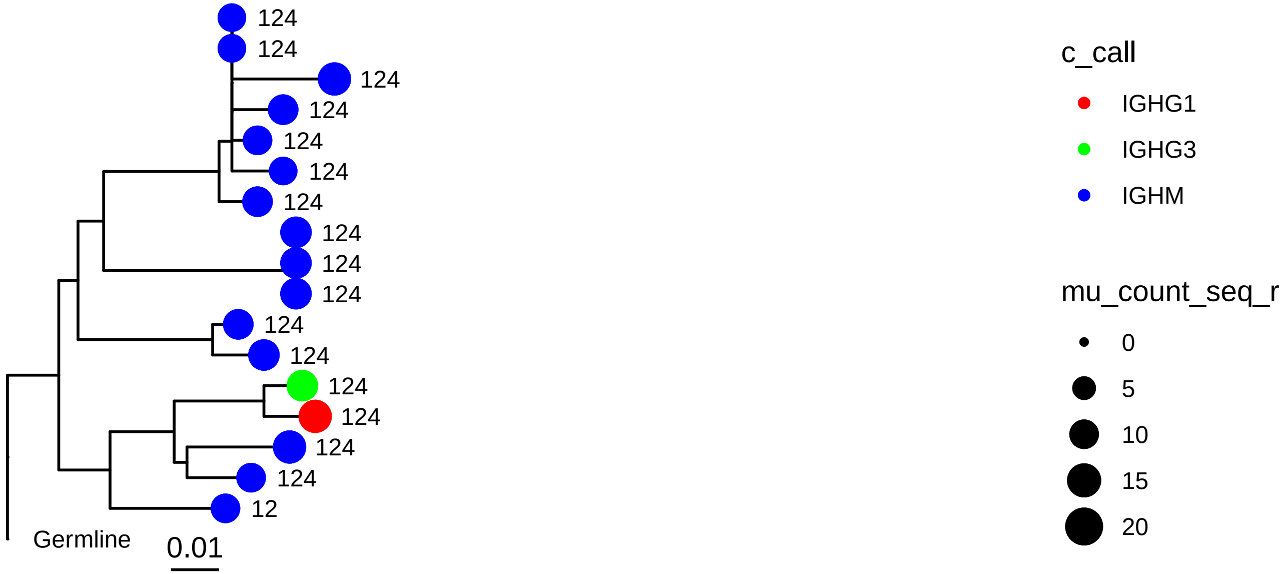
124
124
124
124
124
124
124
124
124
124
124
124
124
124
124
124
12
Germline
0.01
c_call
IGHG1
IGHG3
IGHM
mu_count_seq_r
0
5
10
15
20

4842

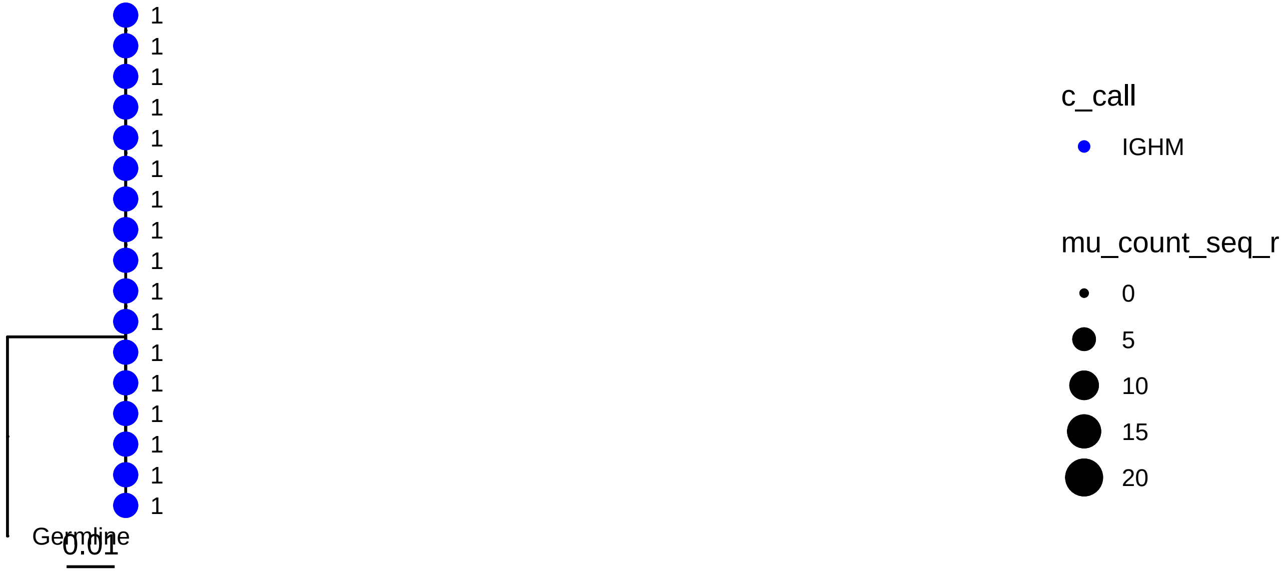
1
1
1
1
1
1
1
1
1
1
1
1
1
1
1
1
1
Germline
0.01
c_call
IGHM
mu_count_seq_r
0
5
10
15
20

973

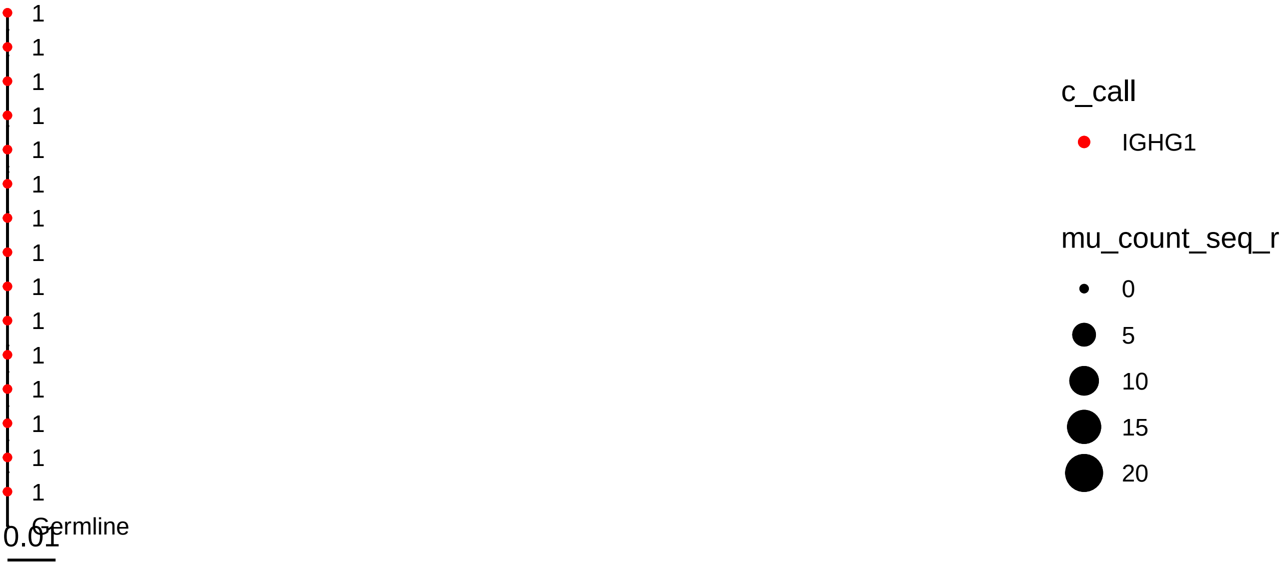
1
1
1
1
1
1
1
1
1
1
1
1
1
1
1
Germline
0.01
c_call
IGHG1
mu_count_seq_r
0
5
10
15
20

4041

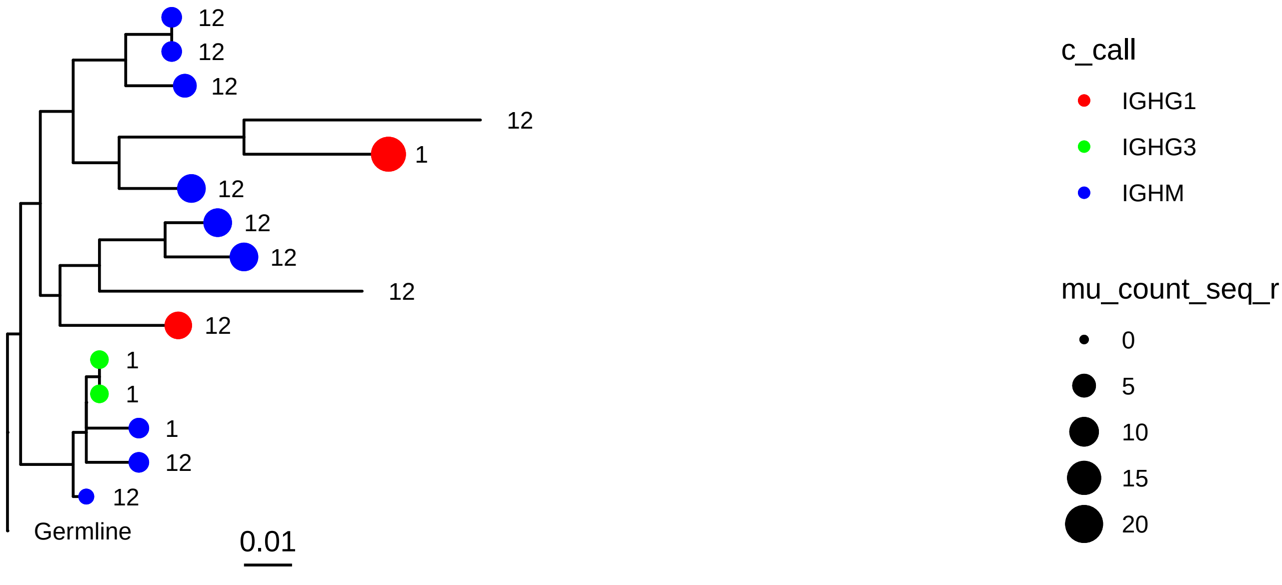
12
12
12
12
1
12
12
12
12
12
1
1
1
12
12
Germline
0.01
c_call
IGHG1
IGHG3
IGHM
mu_count_seq_r
0
5
10
15
20

5041

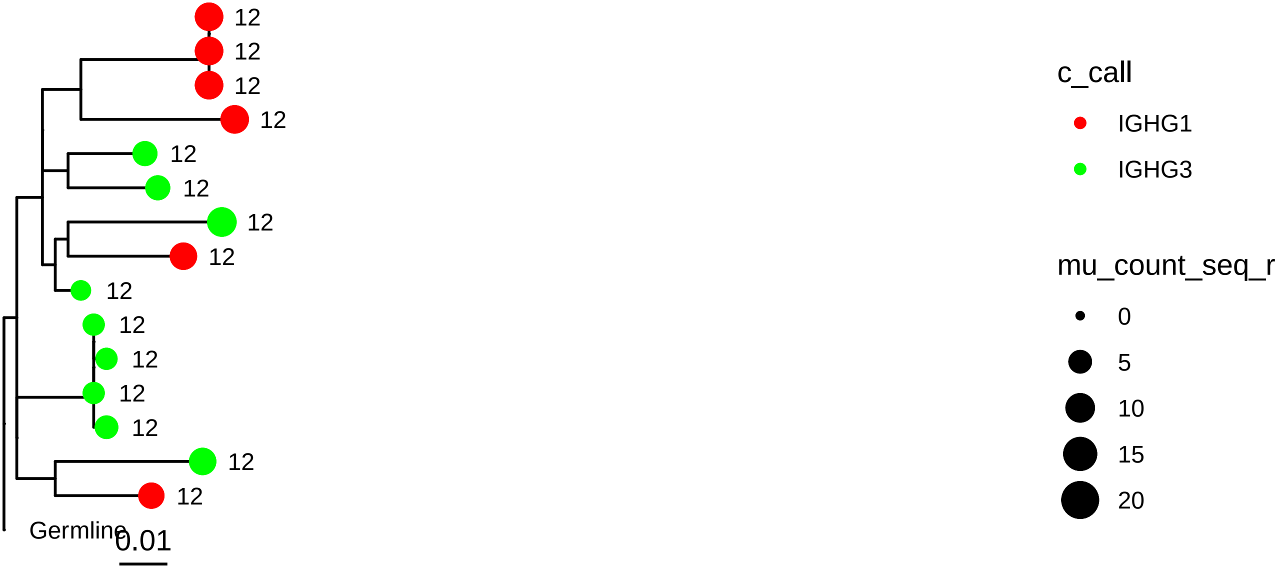

12
12
12
12
12
12
12
12
12
12
12
12
12
12
12
Germline
0.01
c_call
IGHG1
IGHG3
mu_count_seq_r
0
5
10
15
20


4366

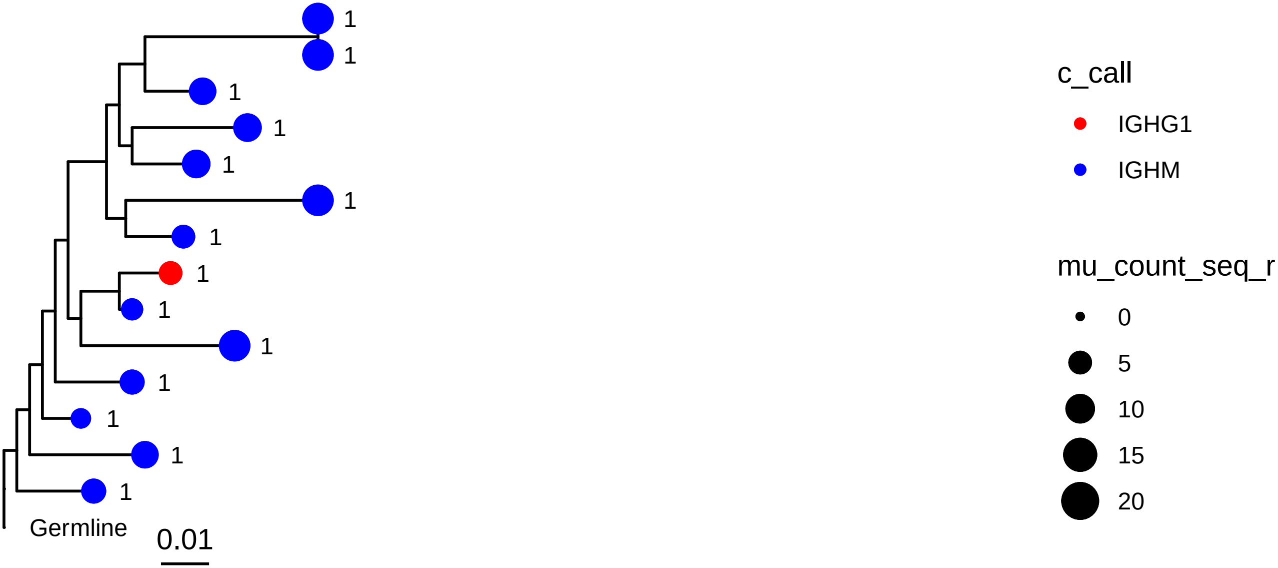

1
1
1
1
1
1
1
1
1
1
1
1
1
1
Germline
0.01
c_call
IGHG1
IGHM
mu_count_seq_r
0
5
10
15
20

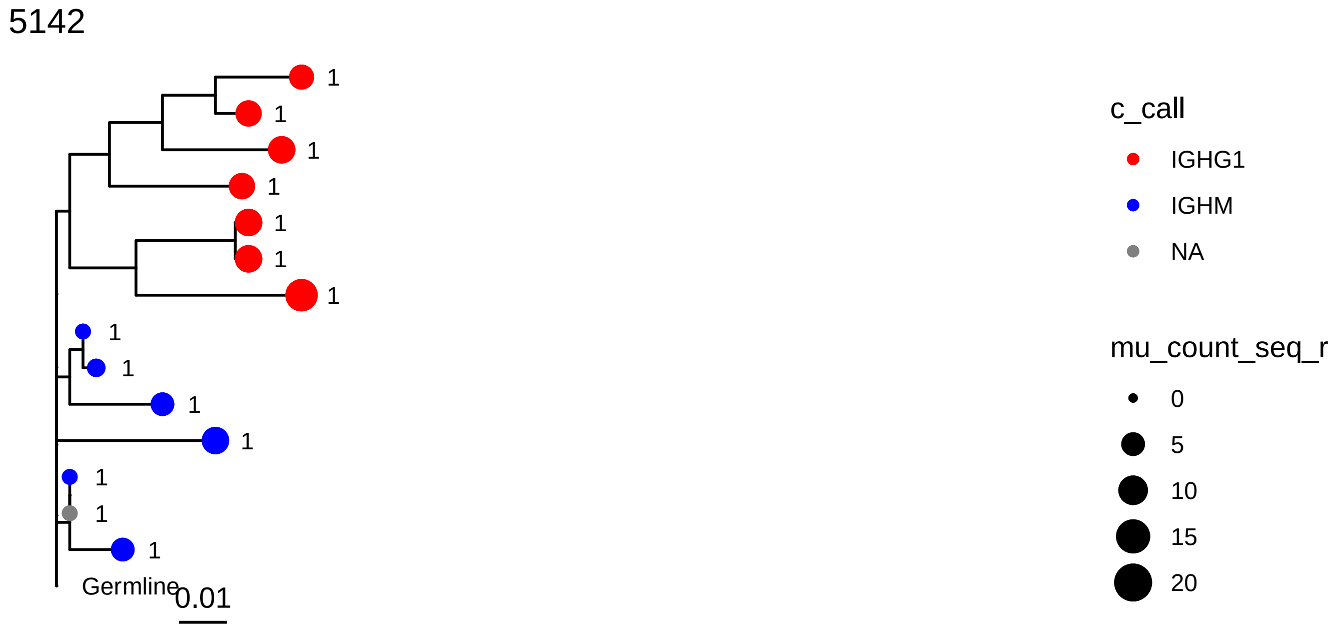
5142
1
1
1
1
1
1
1
1
1
1
1
1
1
1
Germline
0.01
c_call
IGHG1
IGHM
NA
mu_count_seq_r
0
5
10
15
20

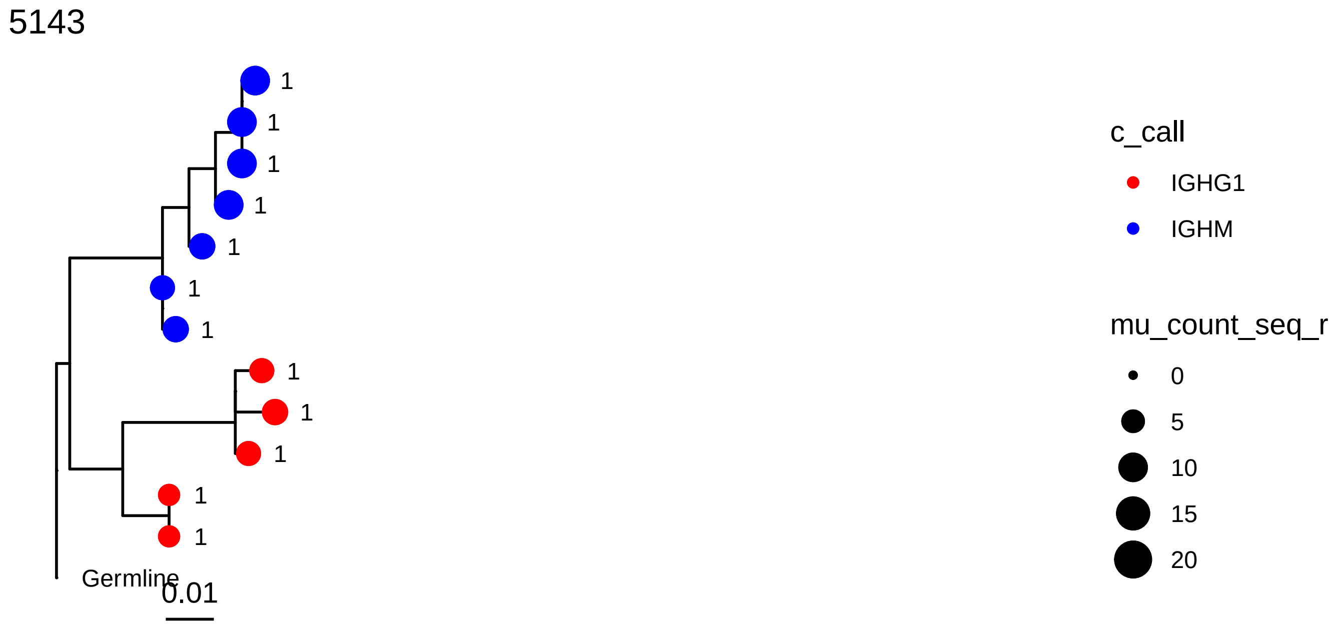
5143
1
1
1
1
1
1
1
1
1
1
1
1
Germline
0.01
c_call
IGHG1
IGHM
mu_count_seq_r
0
5
10
15
20

1650

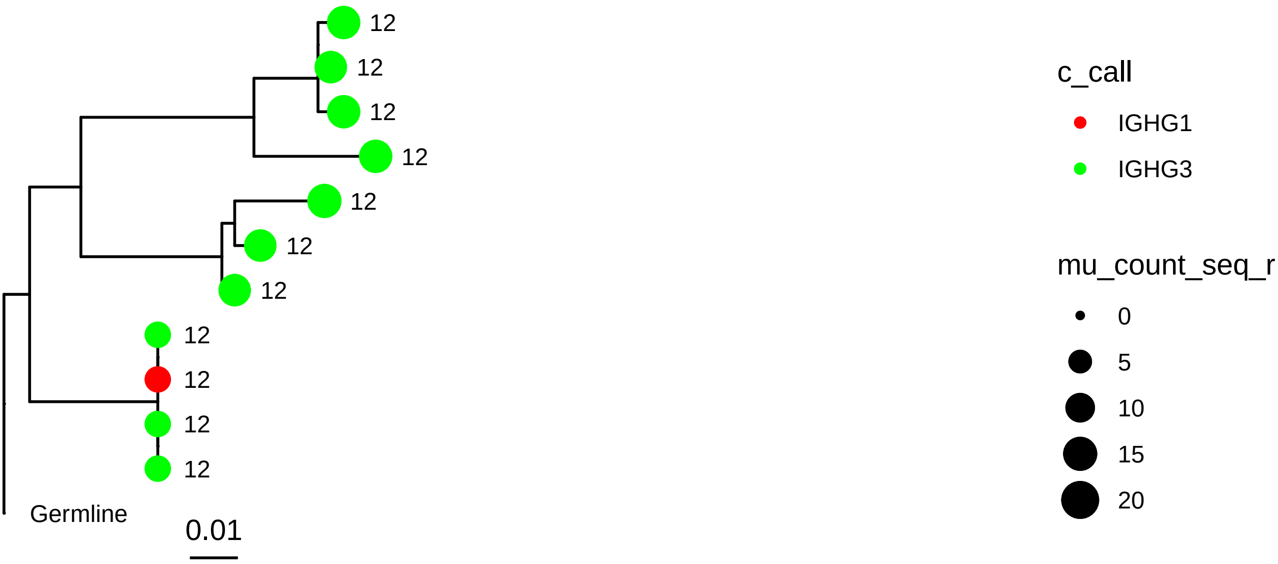
12
12
12
12
12
12
12
12
12
12
12
Germline
0.01
c_call
IGHG1
IGHG3
mu_count_seq_r
0
5
10
15
20

3879

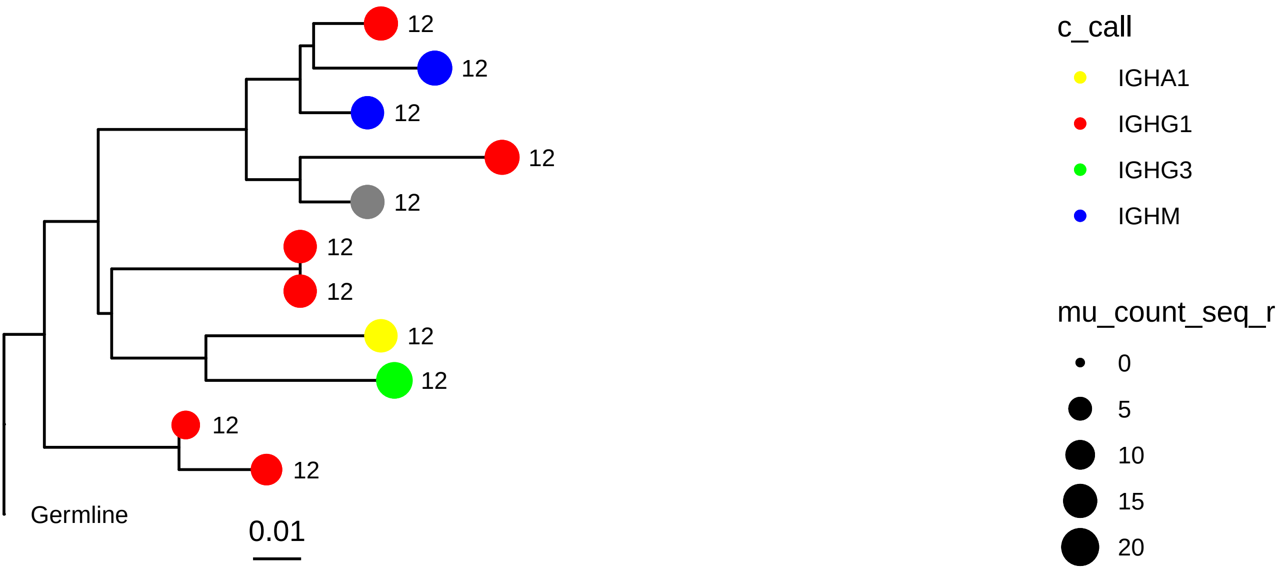
12
12
12
12
12
12
12
12
12
12
12
Germline
0.01
c_call
IGHA1
IGHG1
IGHG3
IGHM
mu_count_seq_r
0
5
10
15
20

5091

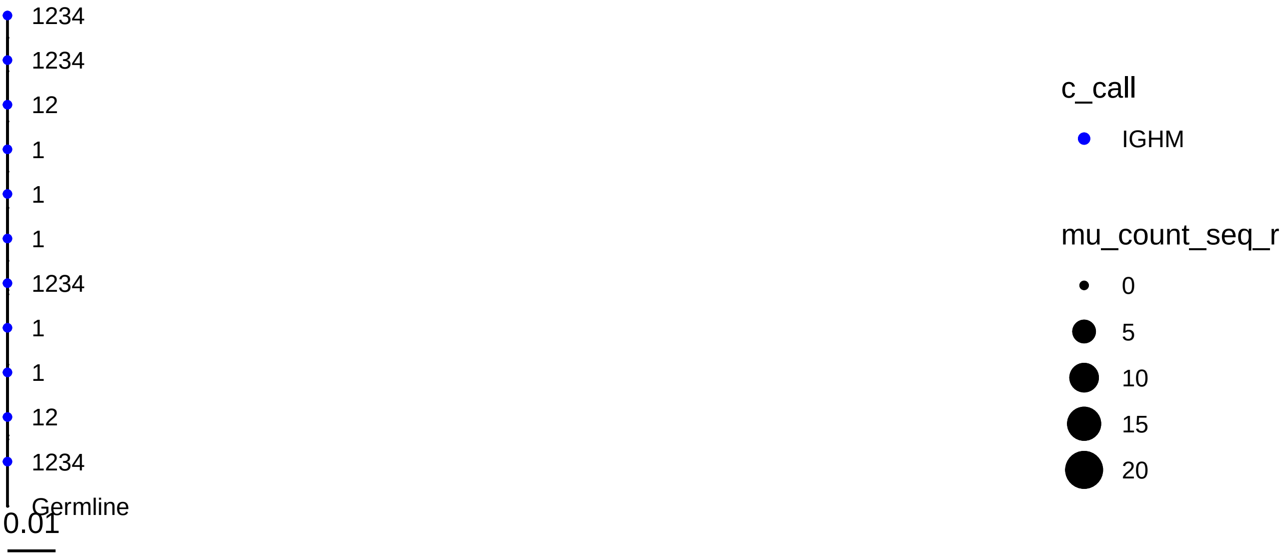
1234
1234
12
1
1
1
1234
1
1
12
1234
Germline
0.01
c_call
IGHM
mu_count_seq_r
0
5
10
15
20

156

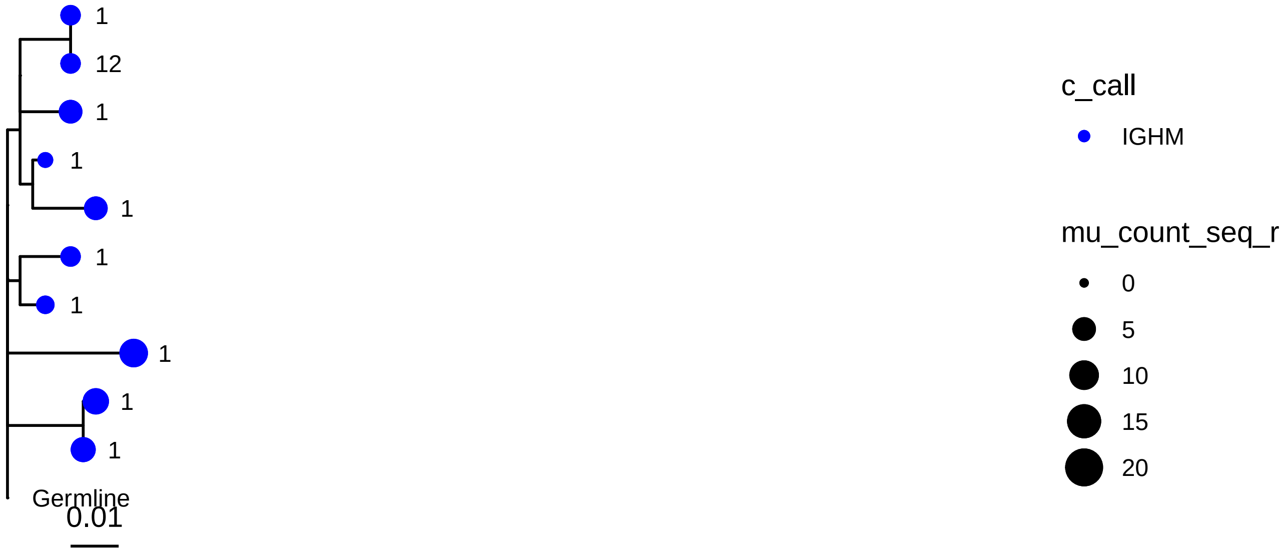
1
12
1
1
1
1
1
1
1
1
Germline
0.01
c_call
IGHM
mu_count_seq_r
0
5
10
15
20

1372

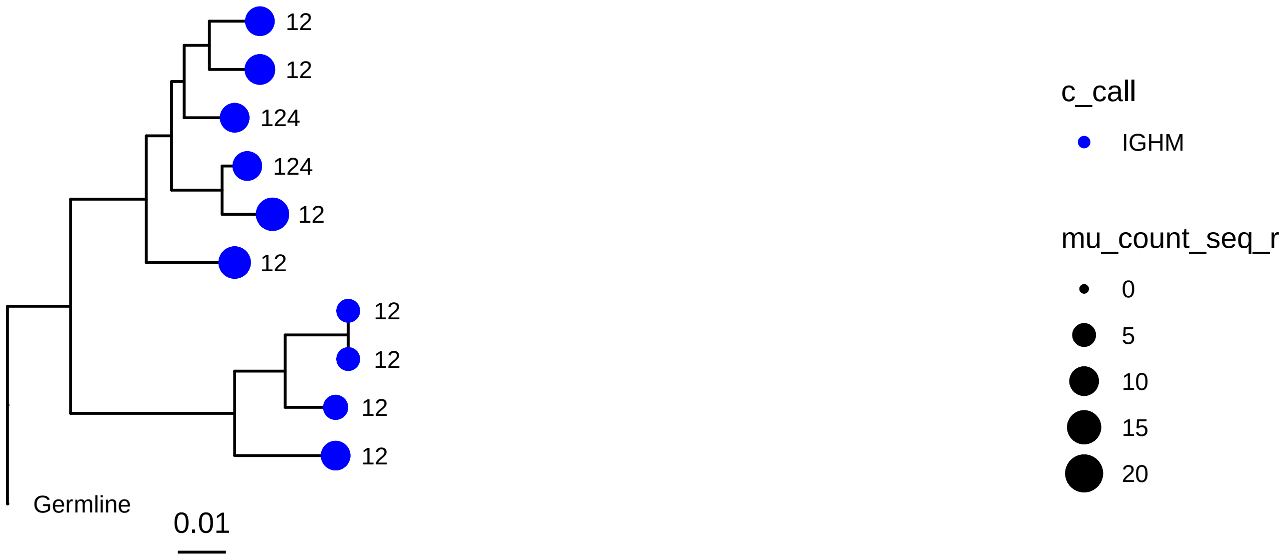
12
12
124
124
12
12
12
12
12
12
Germline
0.01
c_call
IGHM
mu_count_seq_r
0
5
10
15
20

2791

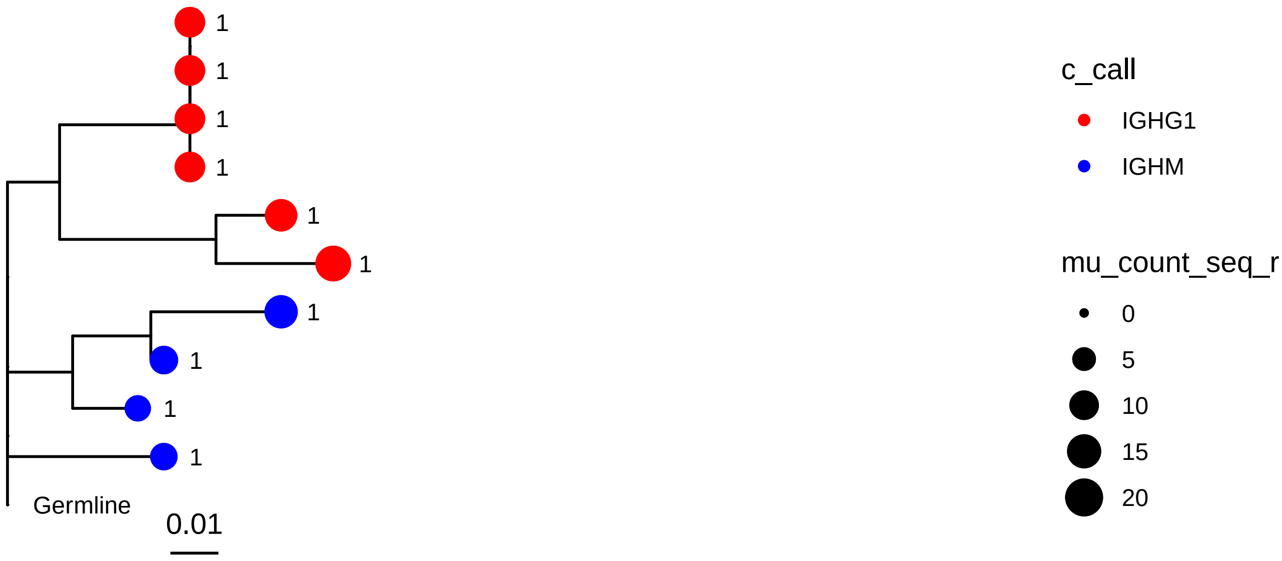
1
1
1
1
1
1
1
1
1
1
Germline
0.01
c_call
IGHG1
IGHM
mu_count_seq_r
0
5
10
15
20

1697

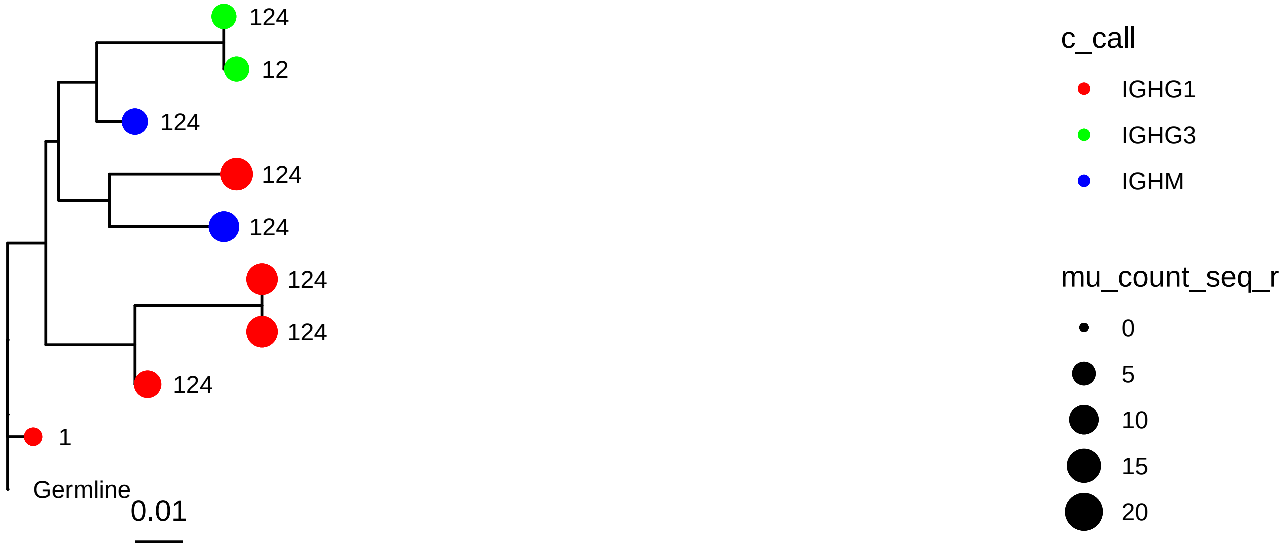
124
12
124
124
124
124
124
124
1
Germline
0.01
c_call
IGHG1
IGHG3
IGHM
mu_count_seq_r
0
5
10
15
20

2671

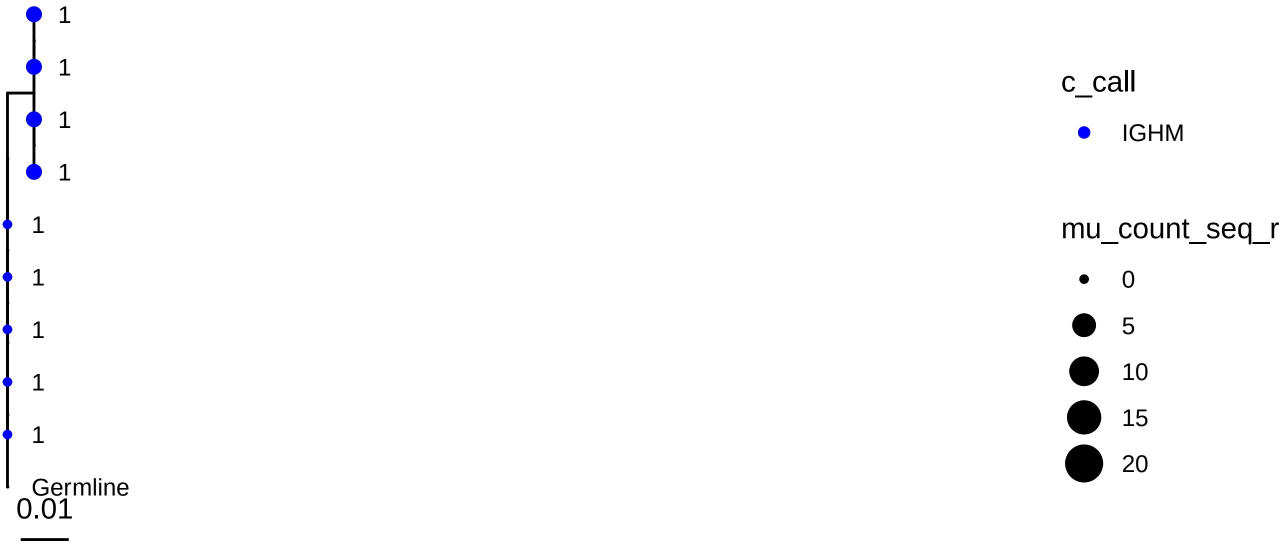
1
1
1
1
1
1
1
1
1
Germline
0.01
c_call
IGHM
mu_count_seq_r
0
5
10
15
20

2956

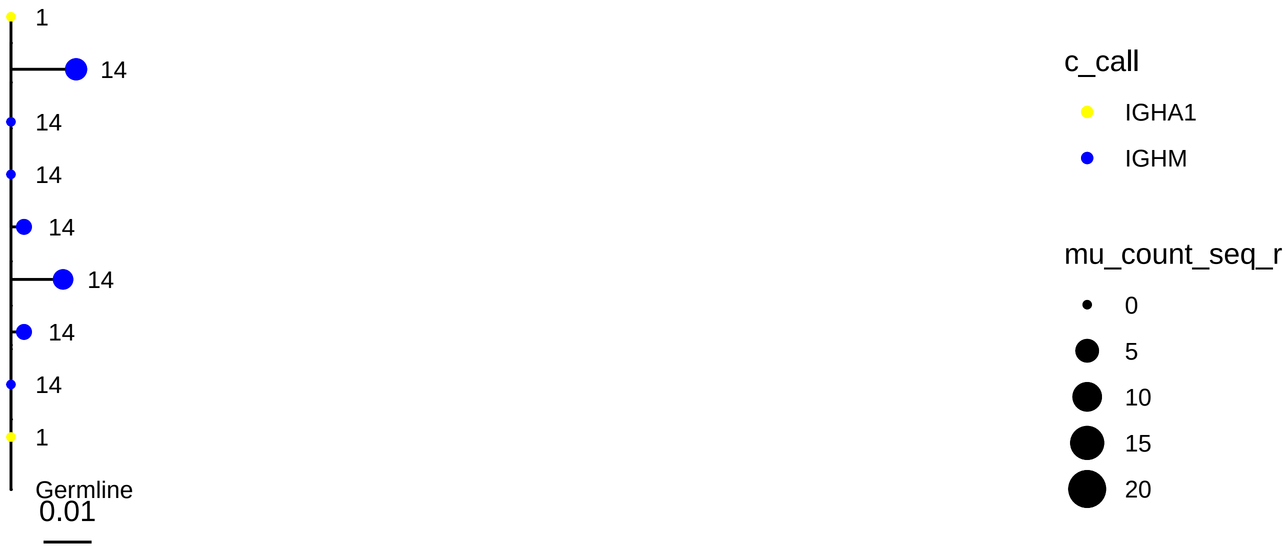
1
14
14
14
14
14
14
14
1
Germline
0.01
c_call
IGHA1
IGHM
mu_count_seq_r
0
5
10
15
20

3233

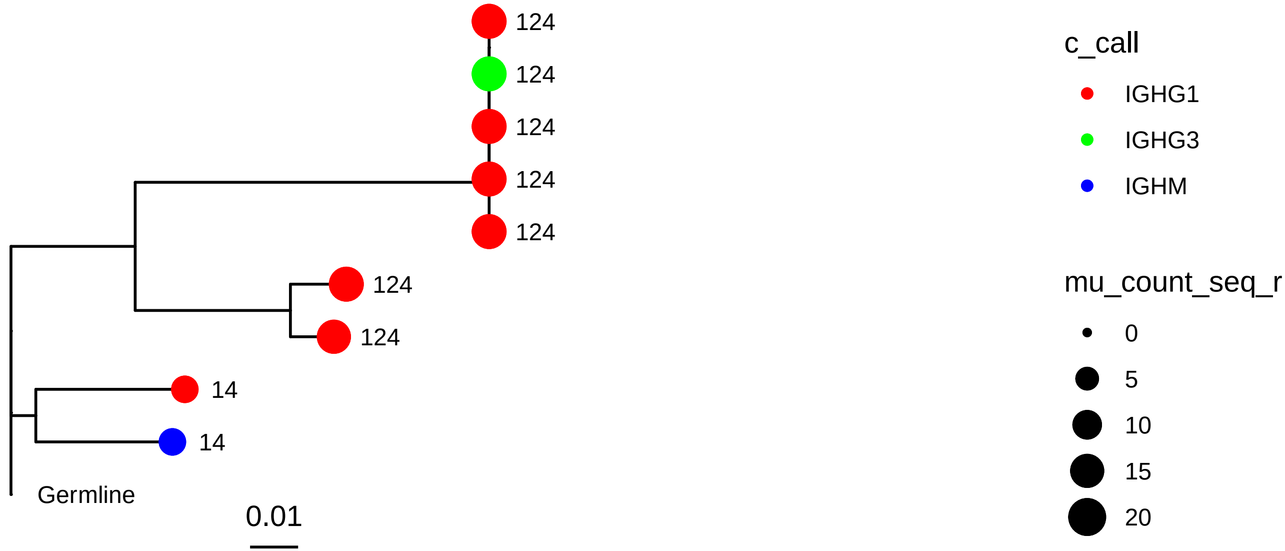
124
124
124
124
124
124
124
14
14
Germline
0.01
c_call
IGHG1
IGHG3
IGHM
mu_count_seq_r
0
5
10
15
20

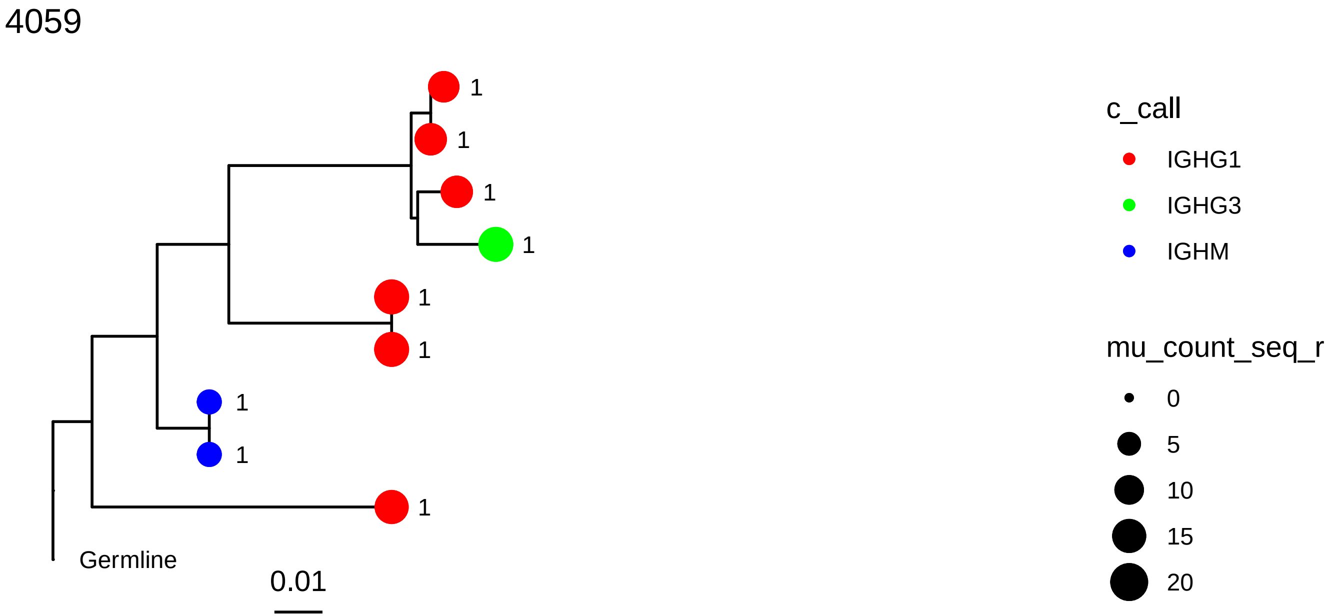
4059
1
1
1
1
1
1
1
1
1
Germline
0.01
c_call
IGHG1
IGHG3
IGHM
mu_count_seq_r
0
5
10
15
20

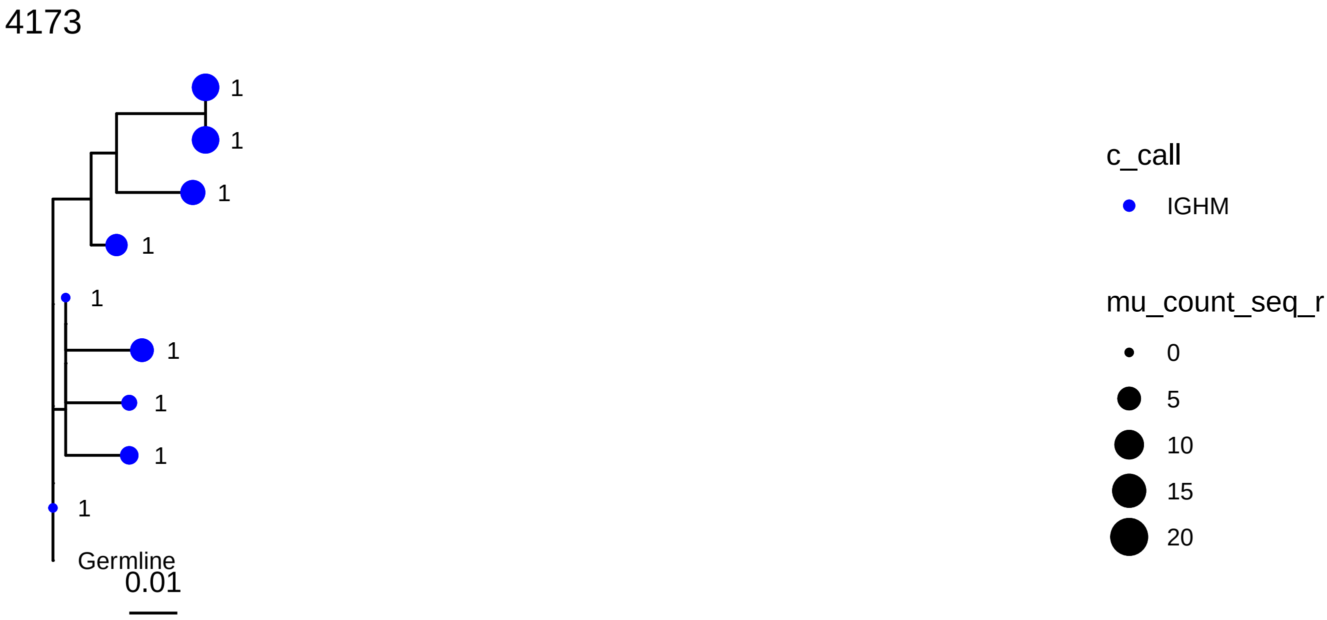
4173
1
1
1
1
1
1
1
1
1
Germline
0.01
c_call
IGHM
mu_count_seq_r
0
5
10
15
20

4417

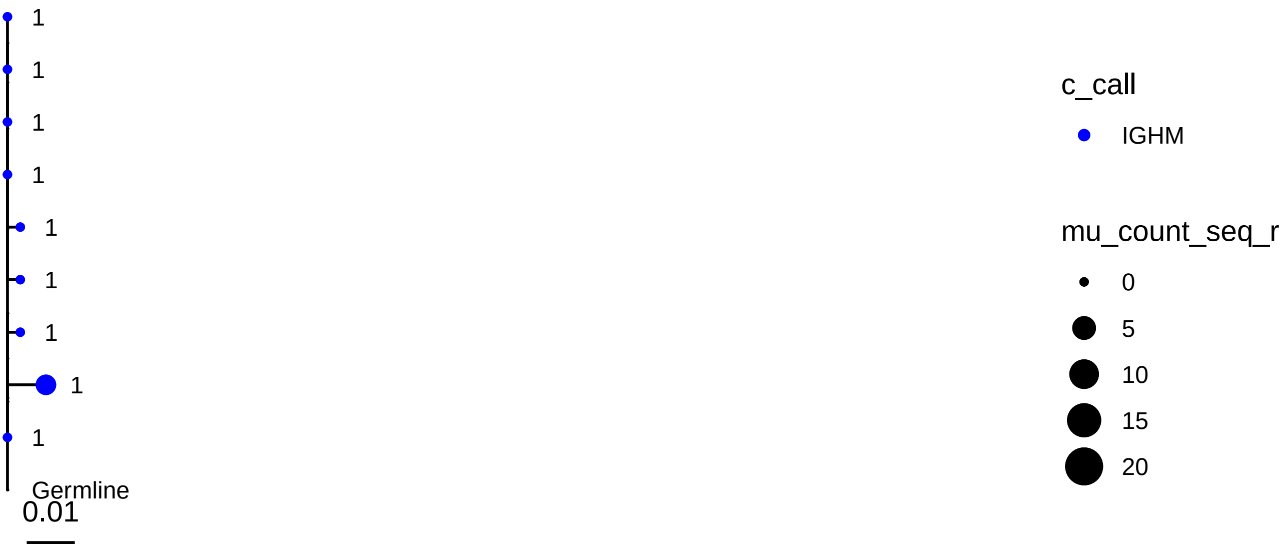
1
1
1
1
1
1
1
1
1
Germline
0.01
c_call
IGHM
mu_count_seq_r
0
5
10
15
20

5013

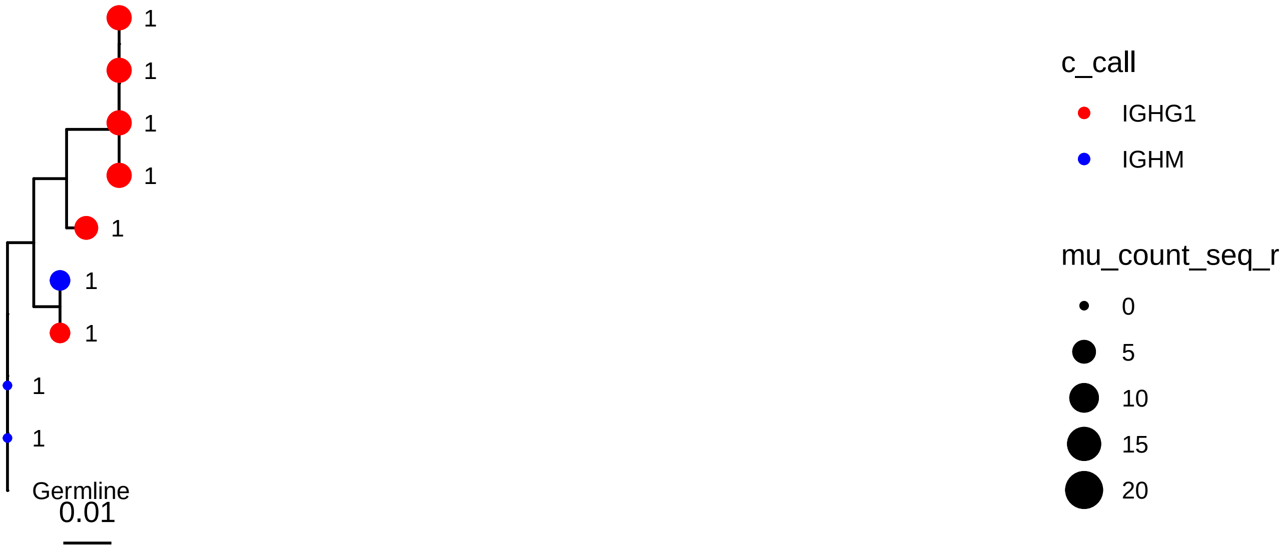
1
1
1
1
1
1
1
1
1
Germline
0.01
c_call
IGHG1
IGHM
mu_count_seq_r
0
5
10
15
20

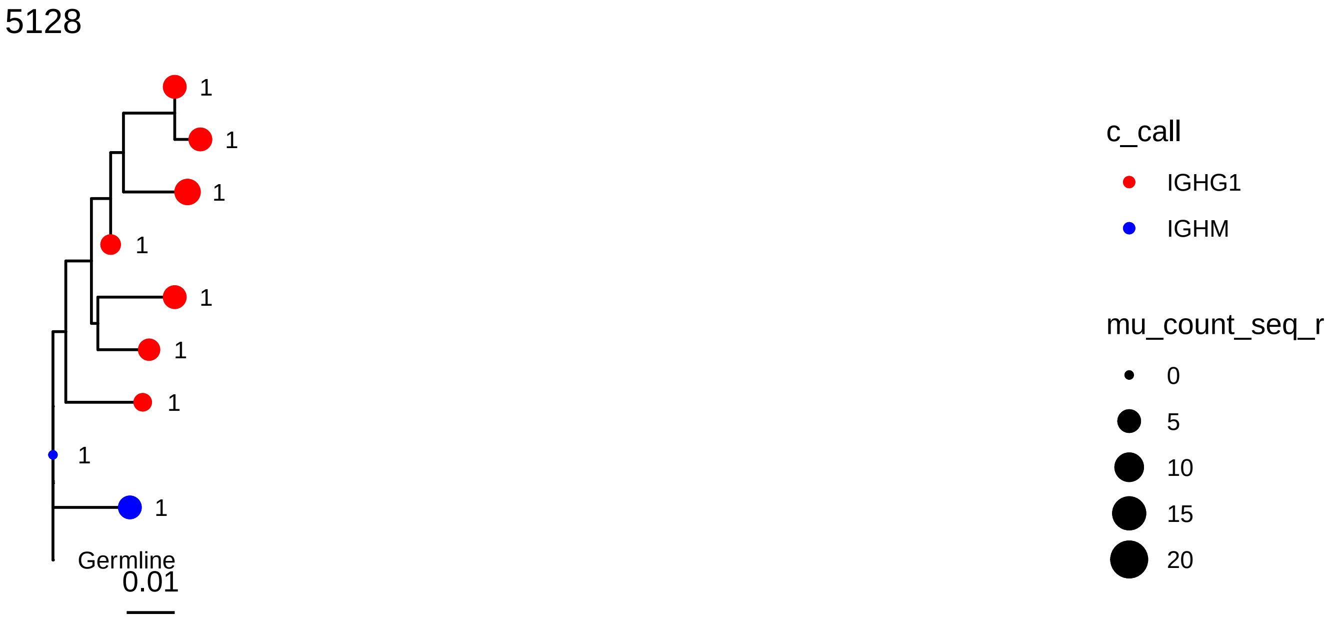
5128
1
1
1
1
1
1
1
1
1
Germline
0.01
c_call
IGHG1
IGHM
mu_count_seq_r
0
5
10
15
20

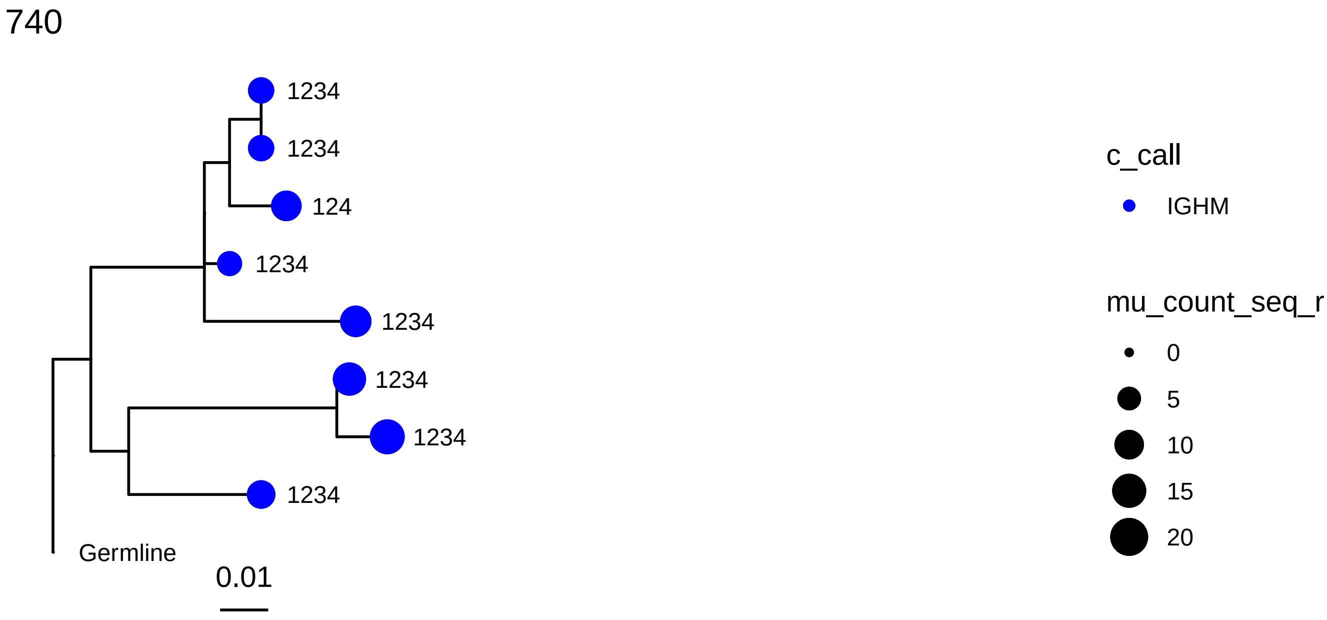
740
1234
1234
124
1234
1234
1234
1234
1234
Germline
0.01
c_call
IGHM
mu_count_seq_r
0
5
10
15
20

1862

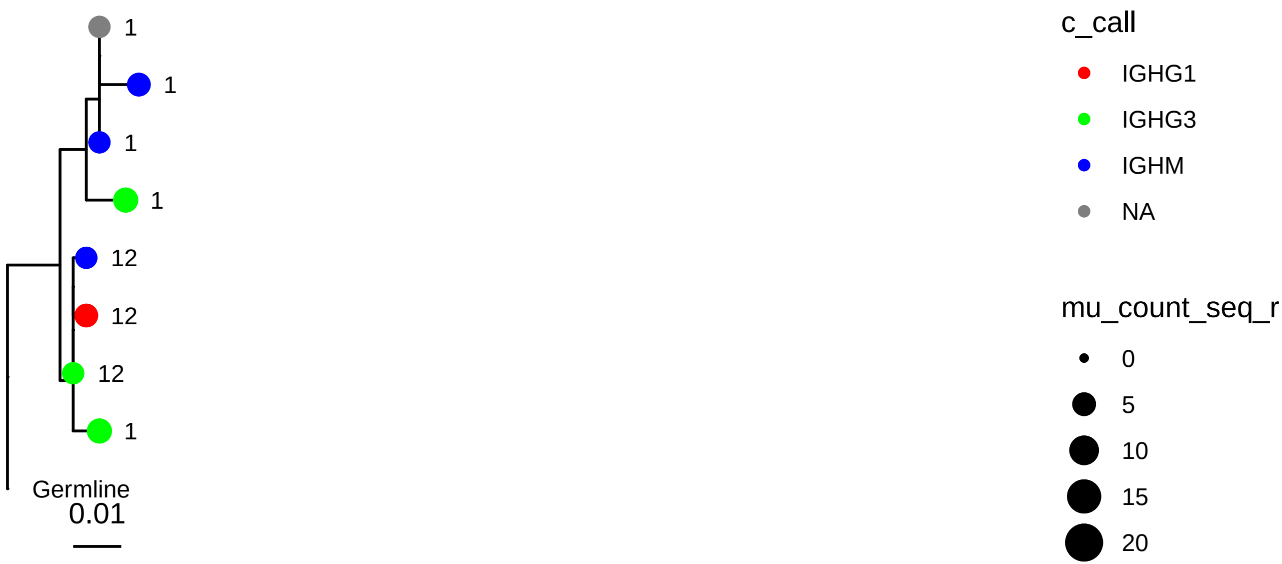
1
1
1
1
12
12
12
1
Germline
0.01
c_call
IGHG1
IGHG3
IGHM
NA
mu_count_seq_r
0
5
10
15
20

3321

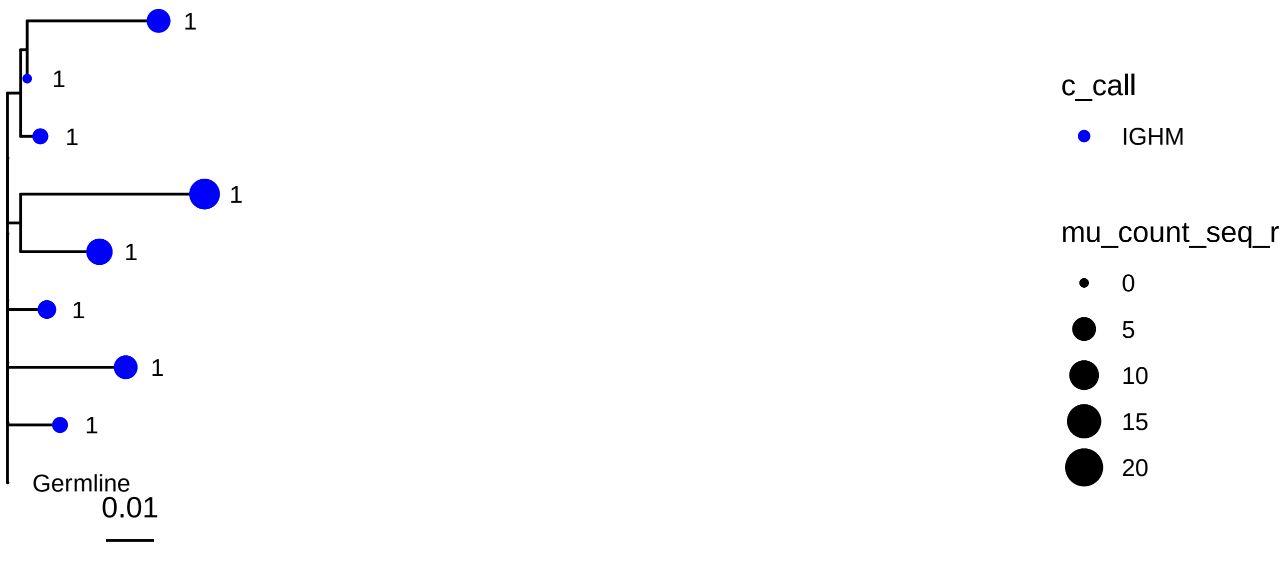
1
1
1
1
1
1
1
1
Germline
0.01
c_call
IGHM
mu_count_seq_r
0
5
10
15
20

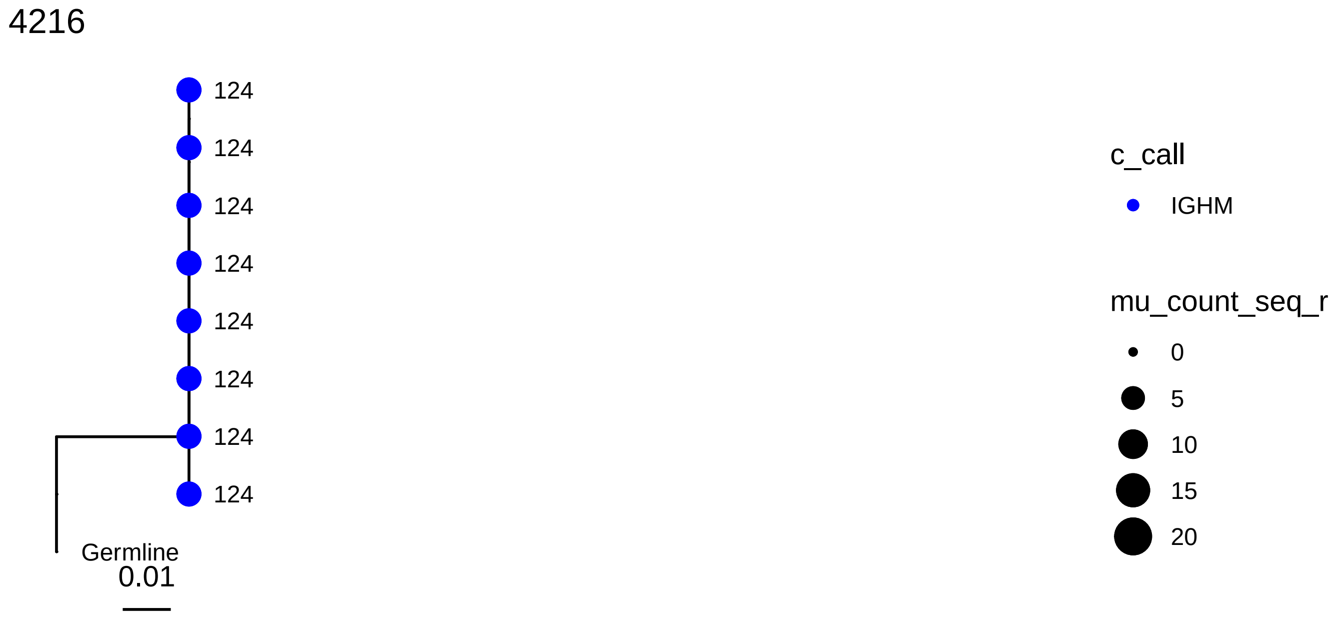
4216
124
124
124
124
124
124
124
124
Germline
0.01
c_call
IGHM
mu_count_seq_r
0
5
10
15
20

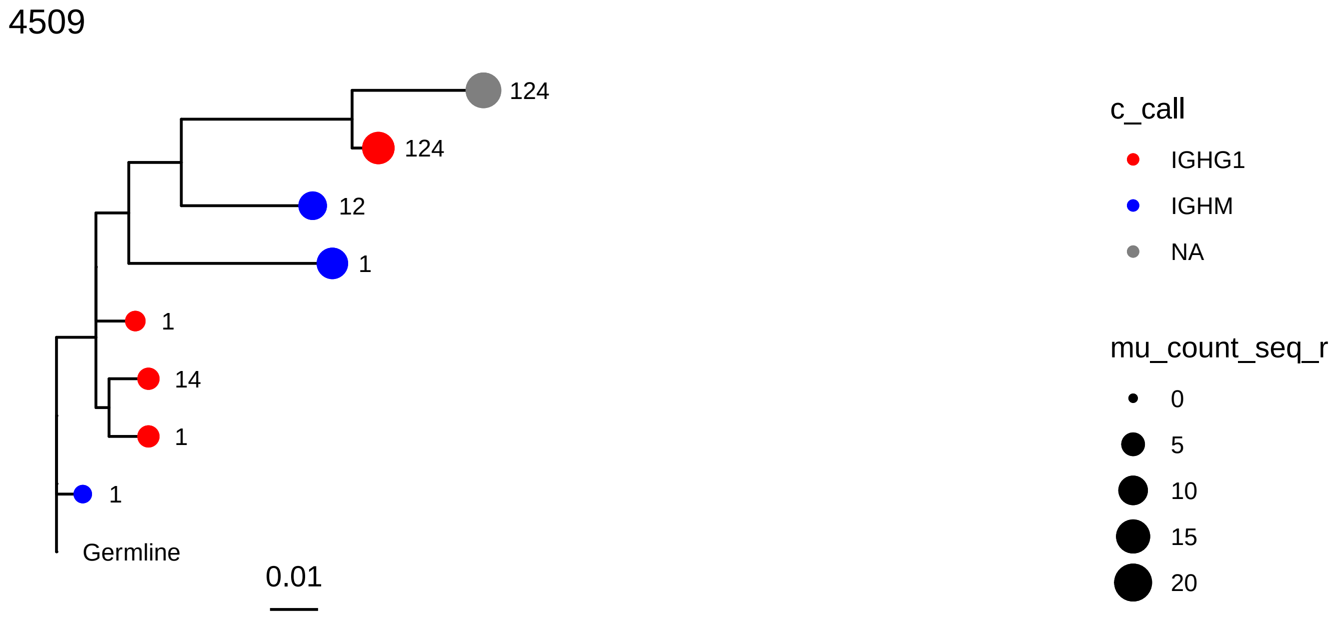
4509
124
124
12
1
1
14
1
1
Germline
0.01
c_call
IGHG1
IGHM
NA
mu_count_seq_r
0
5
10
15
20

5156

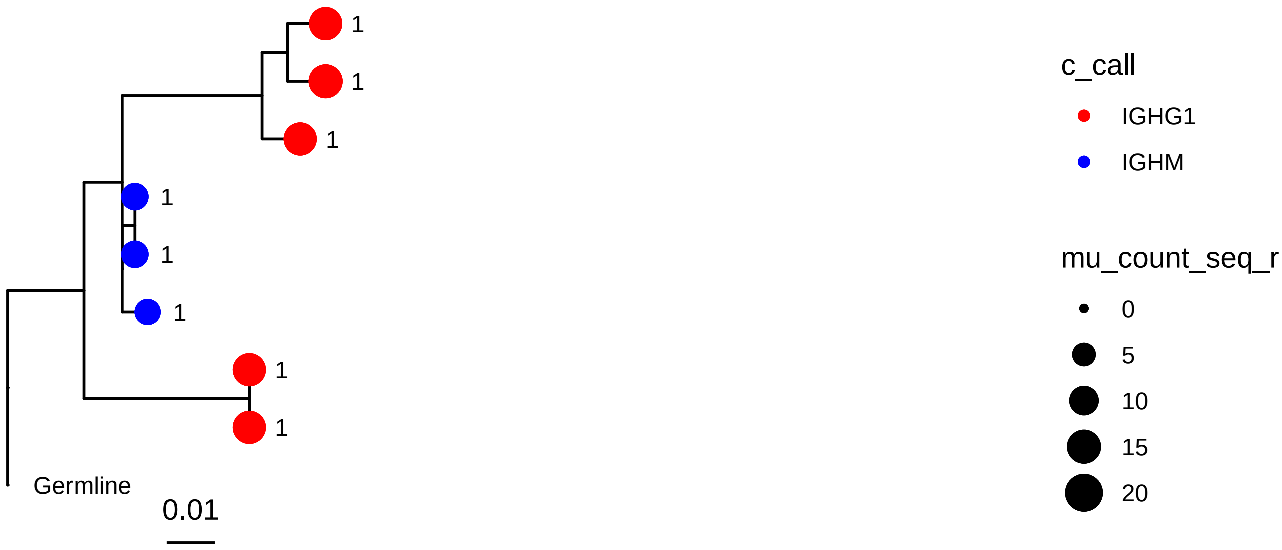
Germline
0.01
c_call
IGHG1
IGHM
mu_count_seq_r
0
5
10
15
20

5684

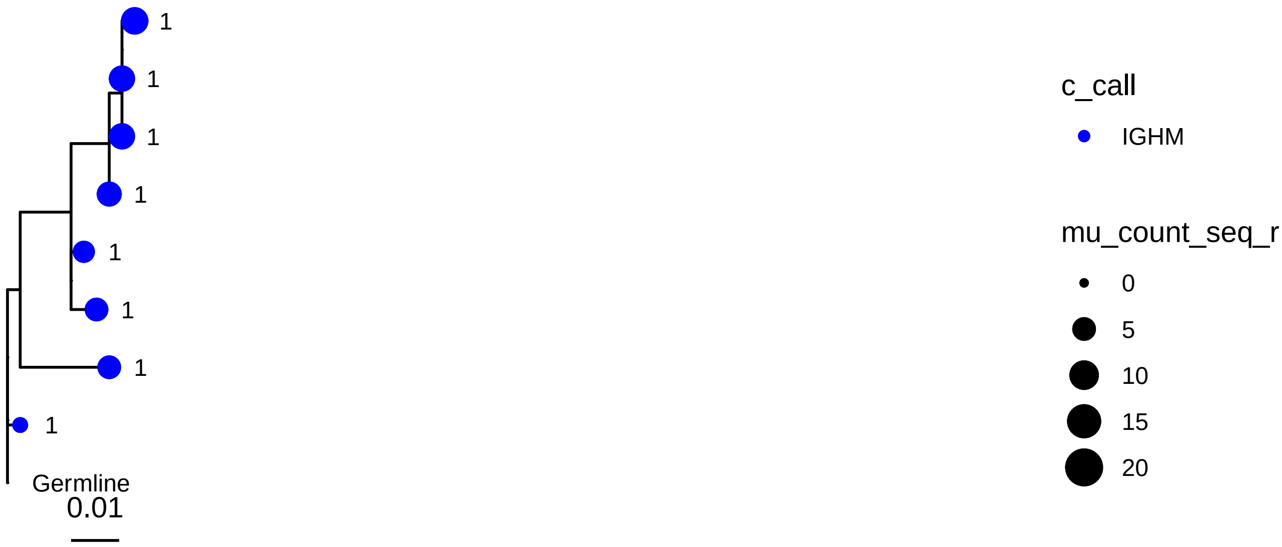
Germline
0.01
c_call
IGHM
mu_count_seq_r
0
5
10
15
20

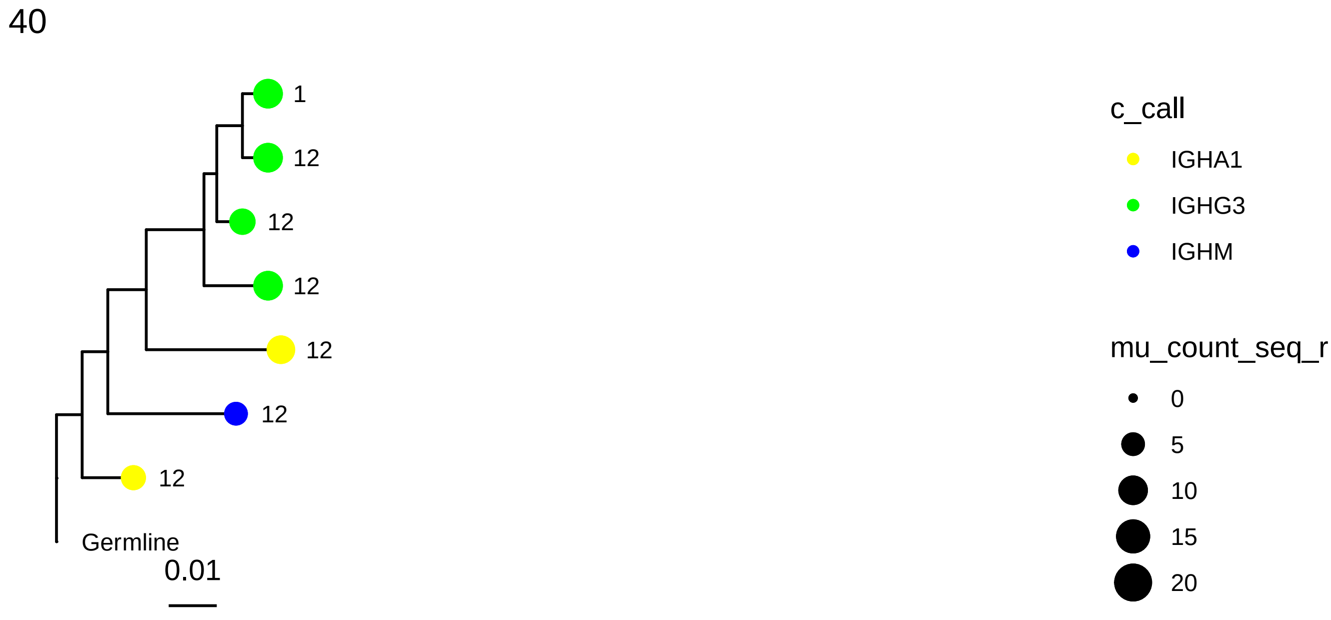
40
1
12
12
12
12
12
12
Germline
0.01
c_call
IGHA1
IGHG3
IGHM
mu_count_seq_r
0
5
10
15
20

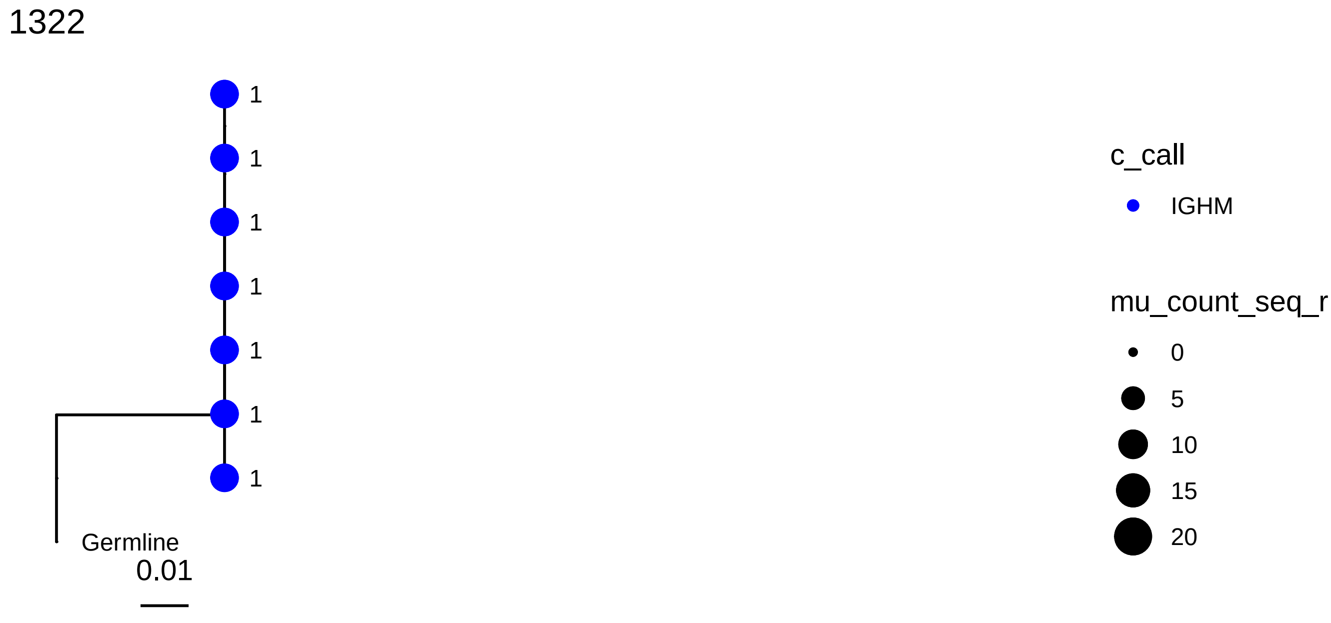
1322
1
1
1
1
1
1
1
Germline
0.01
c_call
IGHM
mu_count_seq_r
0
5
10
15
20

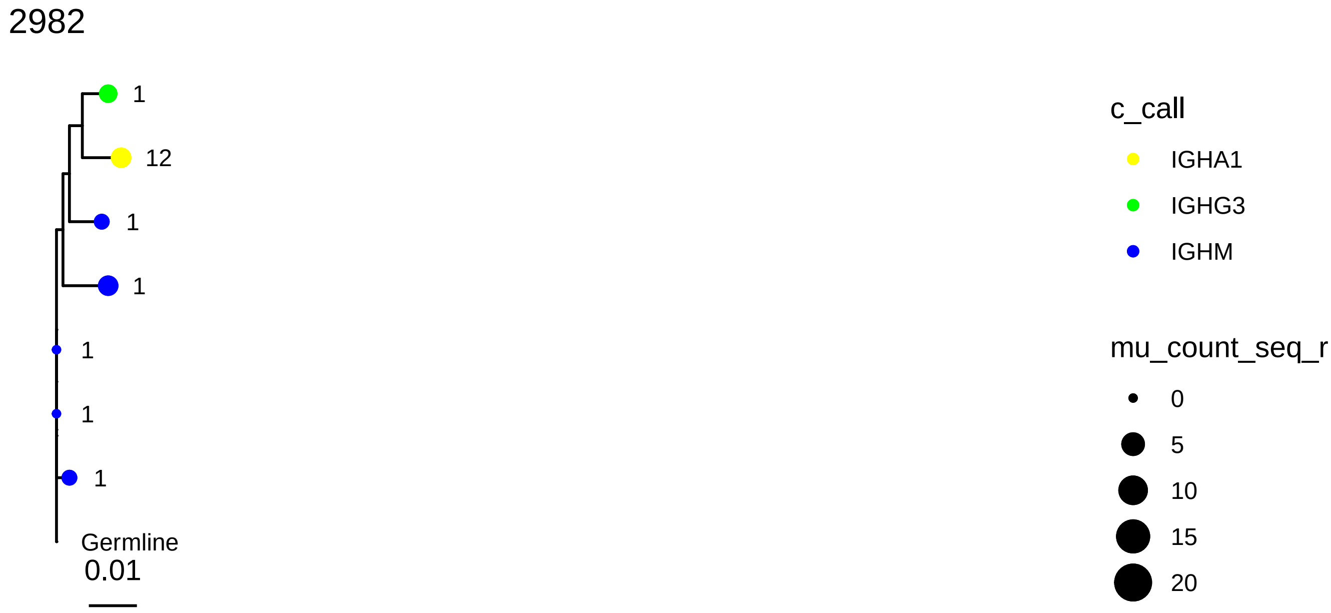

2982
1
12
1
1
1
1
1
Germline
0.01
c_call
IGHA1
IGHG3
IGHM
mu_count_seq_r
0
5
10
15
20


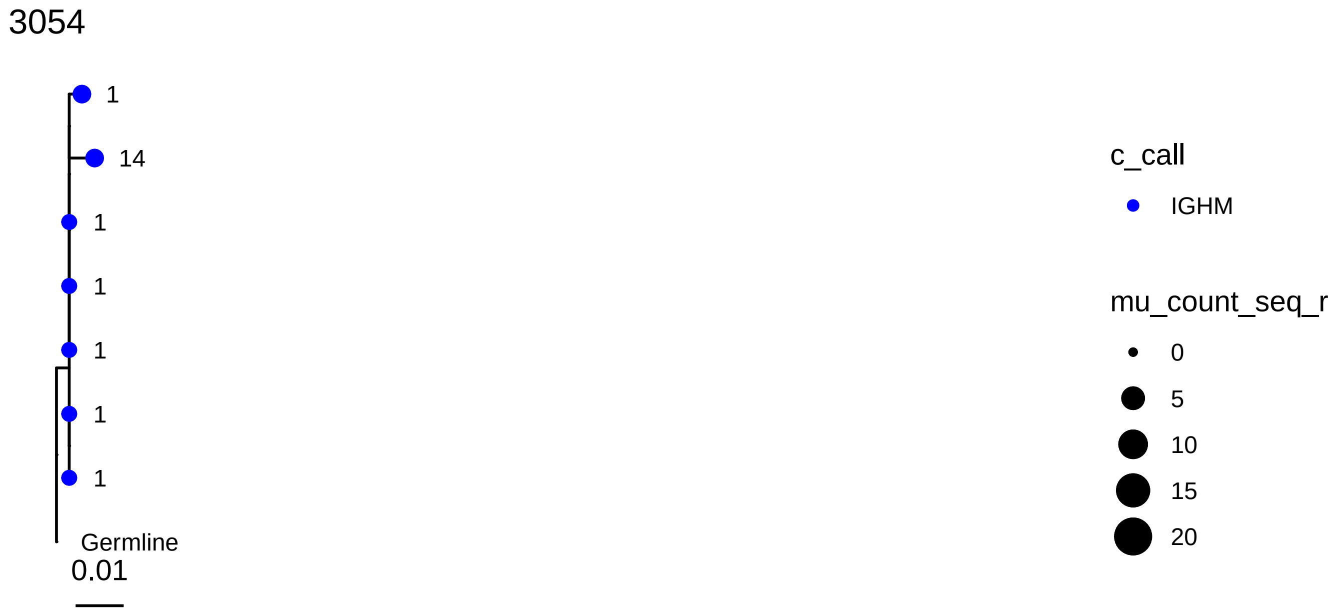

3054
1
14
1
1
1
1
1
Germline
0.01
c_call
IGHM
mu_count_seq_r
0
5
10
15
20

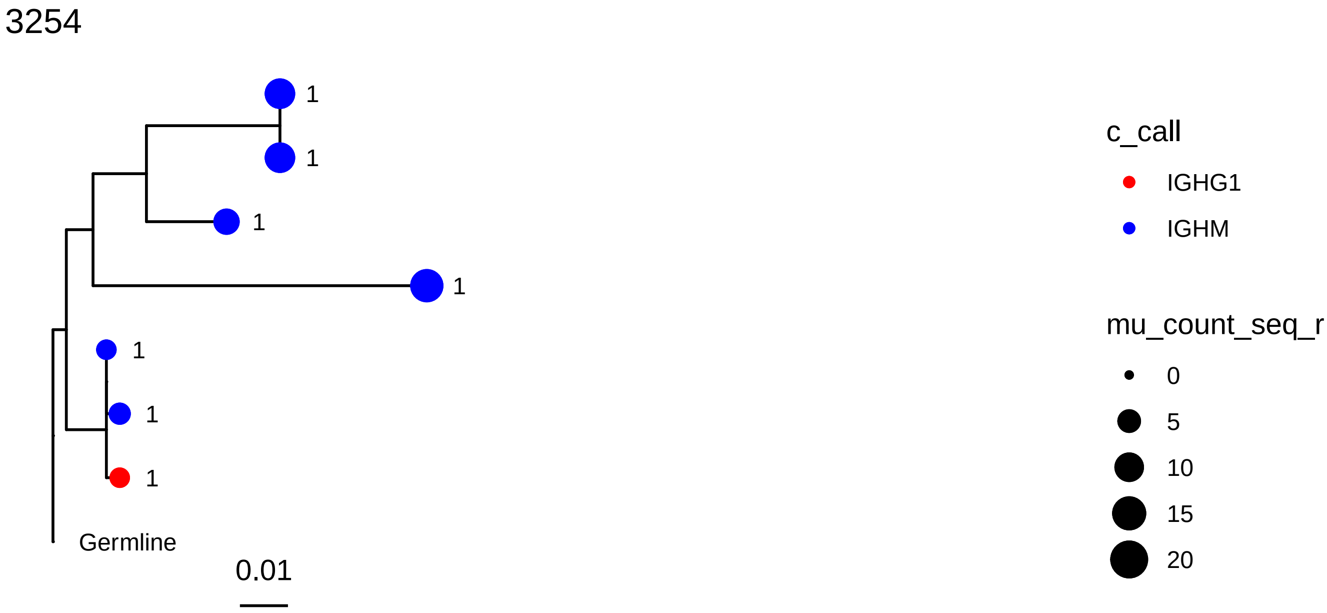
3254
1
1
1
1
1
1
1
Germline
0.01
c_call
IGHG1
IGHM
mu_count_seq_r
0
5
10
15
20

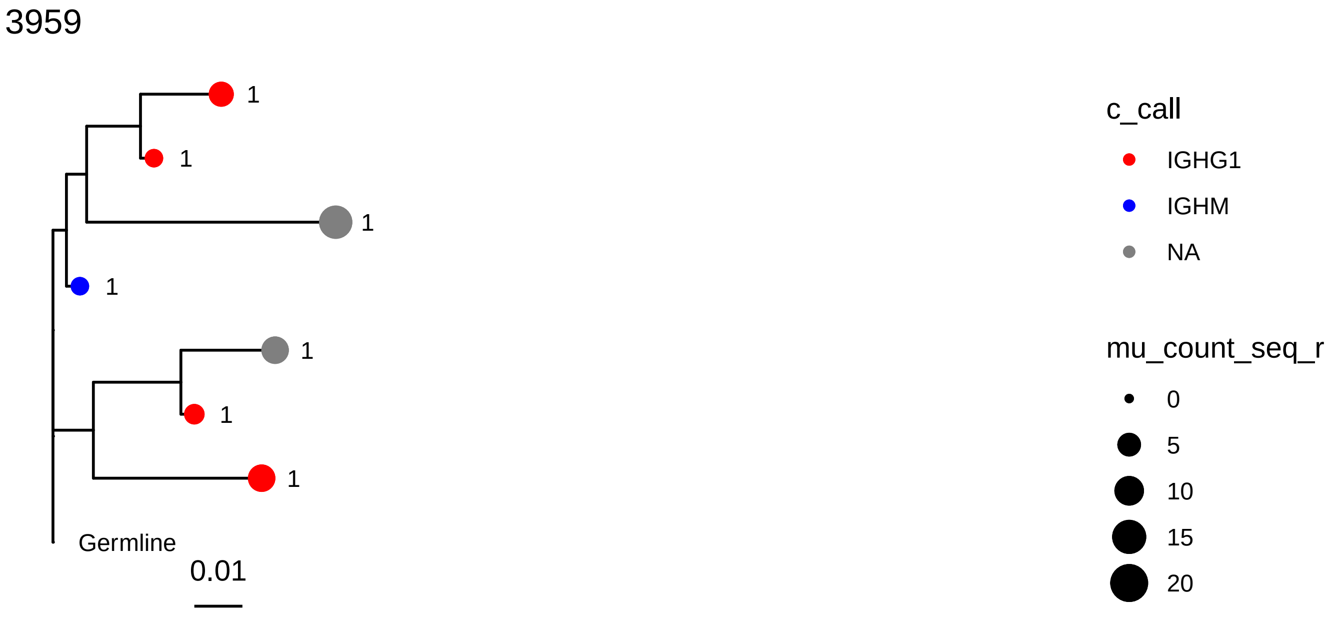
3959
1
1
1
1
1
1
1
Germline
0.01
c_call
IGHG1
IGHM
NA
mu_count_seq_r
0
5
10
15
20

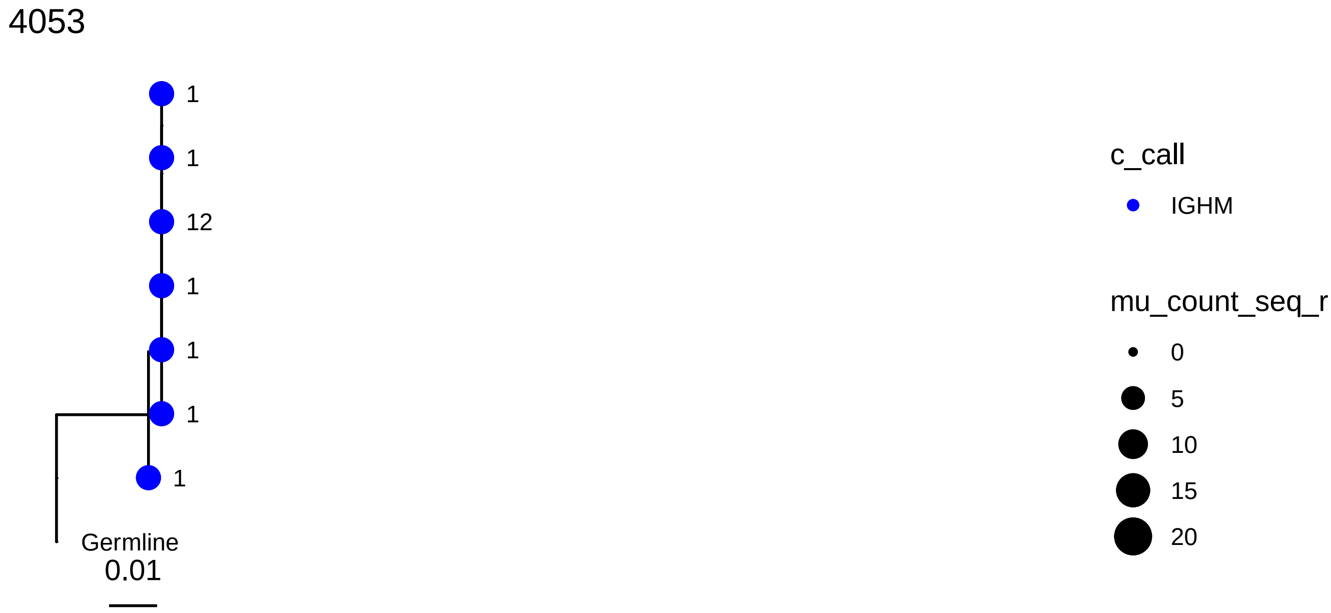
4053
1
1
12
1
1
1
1
Germline
0.01
c_call
IGHM
mu_count_seq_r
0
5
10
15
20

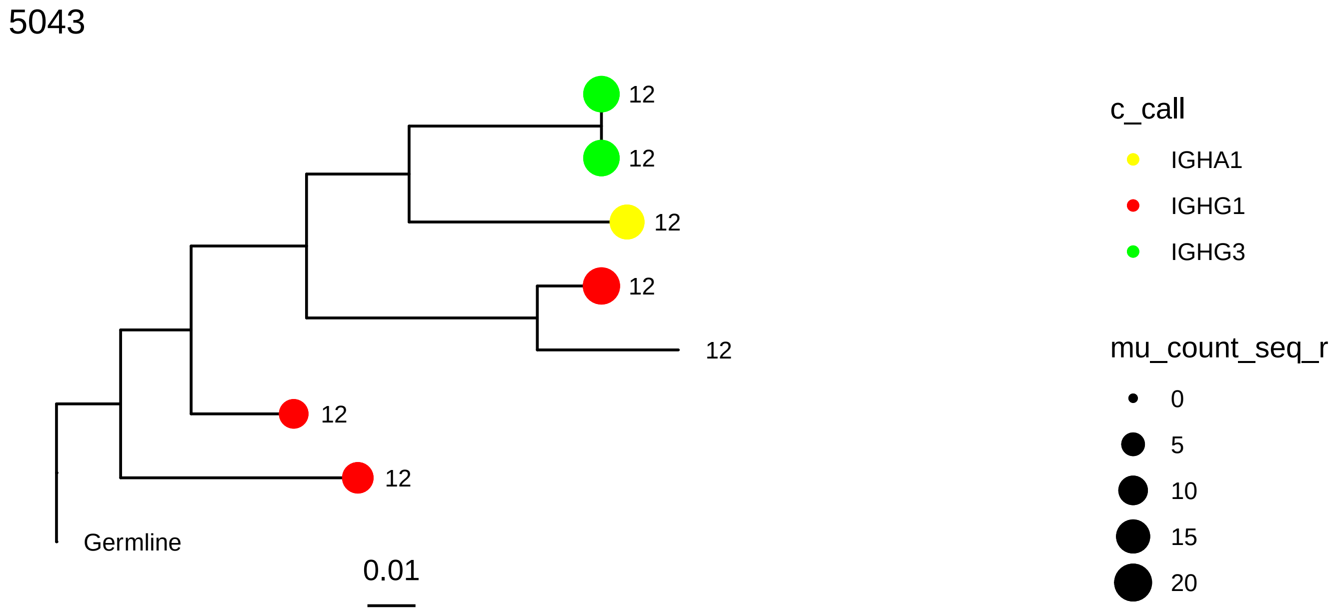
5043
12
12
12
12
12
12
12
Germline
0.01
c_call
IGHA1
IGHG1
IGHG3
mu_count_seq_r
0
5
10
15
20

5248

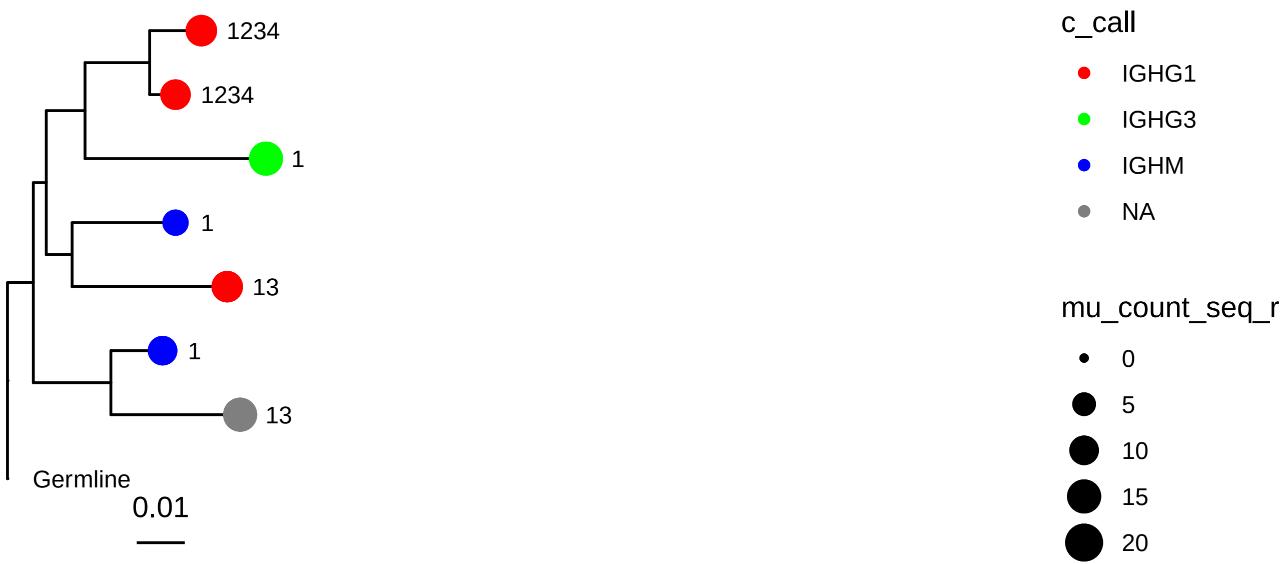
1234
1234
1
1
13
1
13
Germline
0.01
c_call
IGHG1
IGHG3
IGHM
NA
mu_count_seq_r
0
5
10
15
20

5348

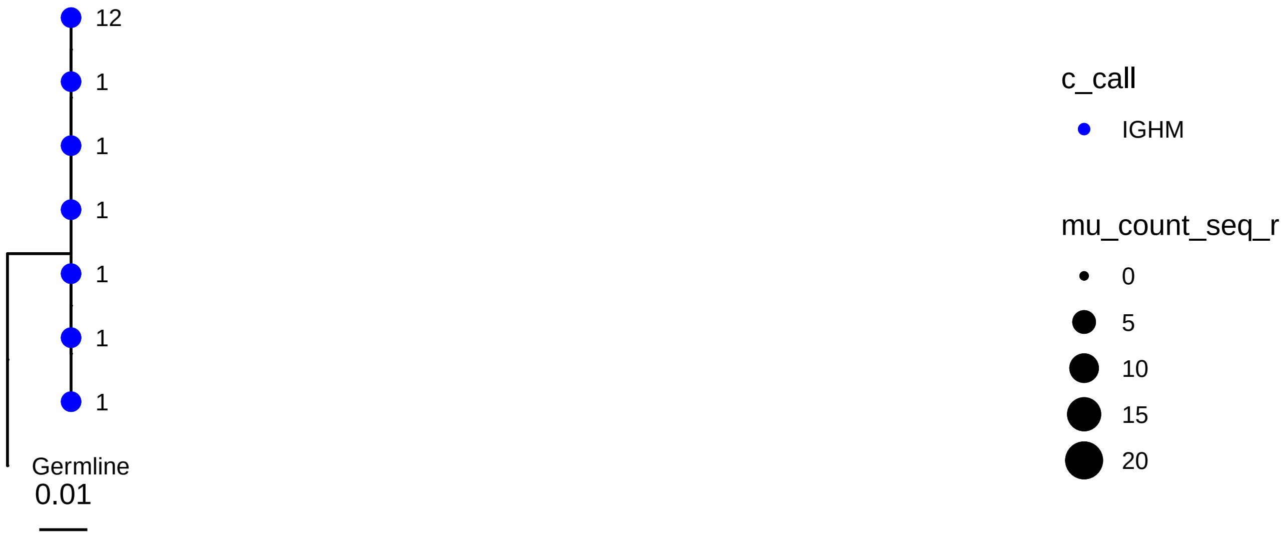
12
1
1
1
1
1
1
Germline
0.01
c_call
IGHM
mu_count_seq_r
0
5
10
15
20

61

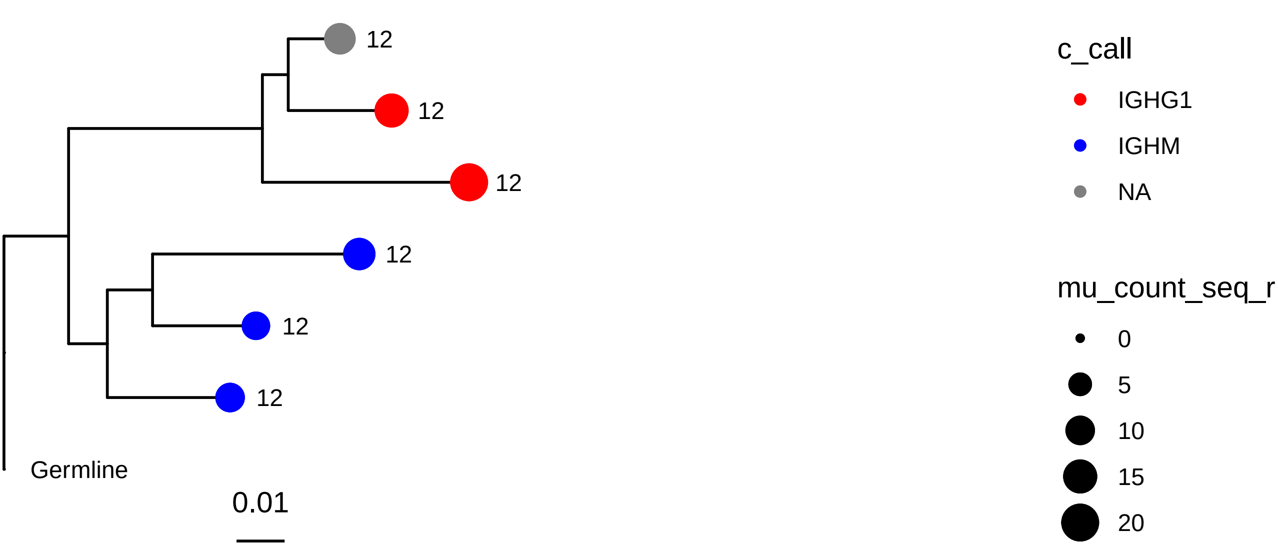
12
12
12
12
12
12
Germline
0.01
c_call
IGHG1
IGHM
NA
mu_count_seq_r
0
5
10
15
20

188

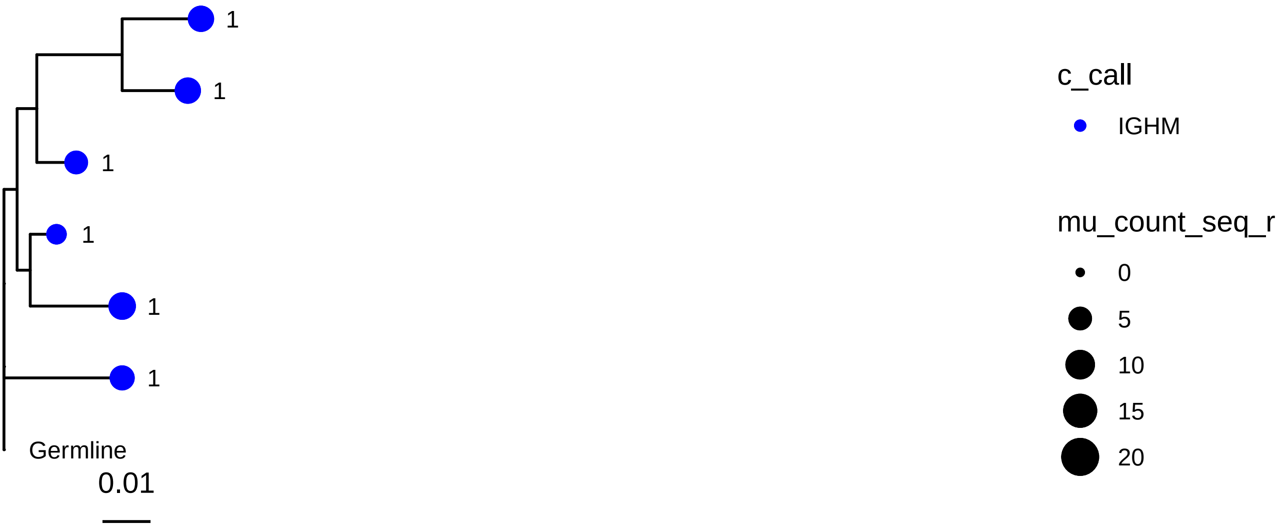
1
1
1
1
1
1
Germline
0.01
c_call
IGHM
mu_count_seq_r
0
5
10
15
20

613

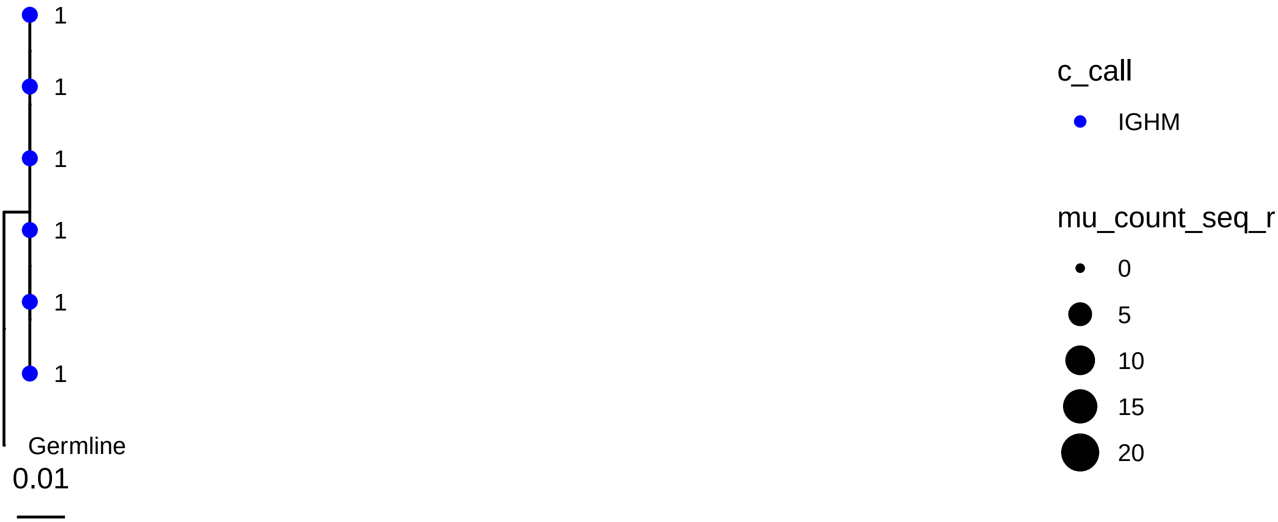
1
1
1
1
1
1
Germline
0.01
c_call
IGHM
mu_count_seq_r
0
5
10
15
20

728

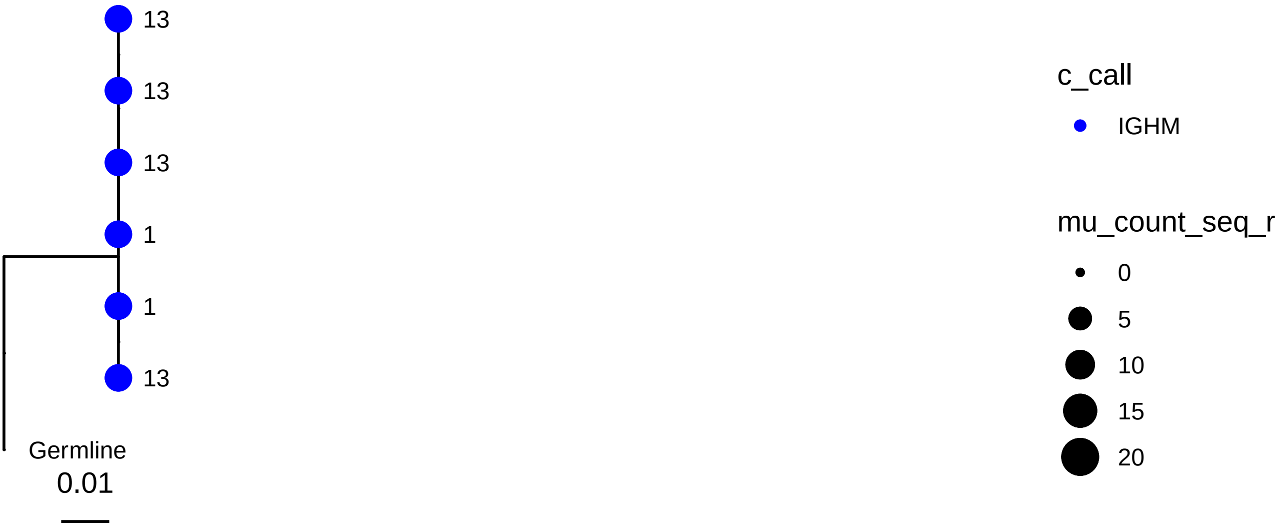
13
13
13
1
1
13
Germline
0.01
c_call
IGHM
mu_count_seq_r
0
5
10
15
20

898

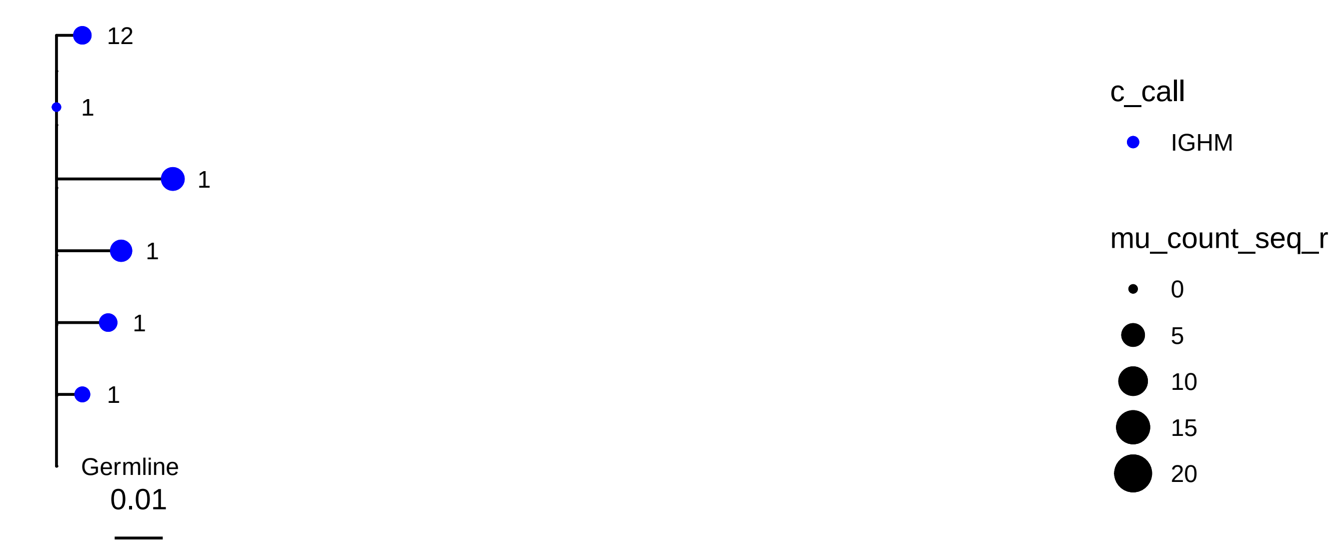
12
1
1
1
1
1
Germline
0.01
c_call
IGHM
mu_count_seq_r
0
5
10
15
20

950

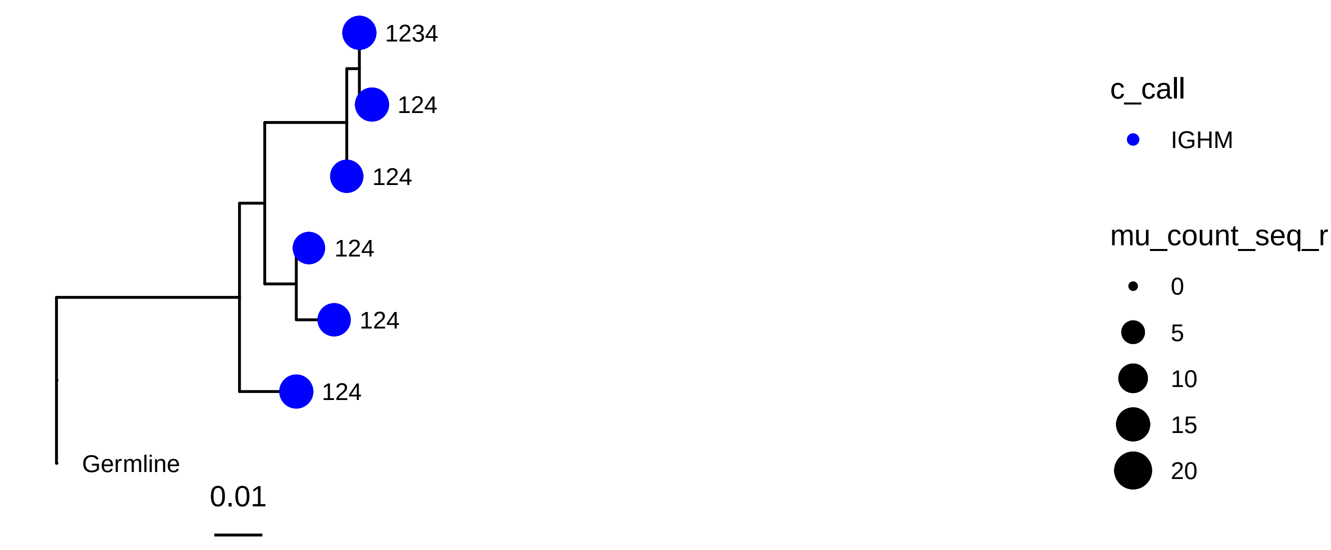
1234
124
124
124
124
124
Germline
0.01
c_call
IGHM
mu_count_seq_r
0
5
10
15
20

1278

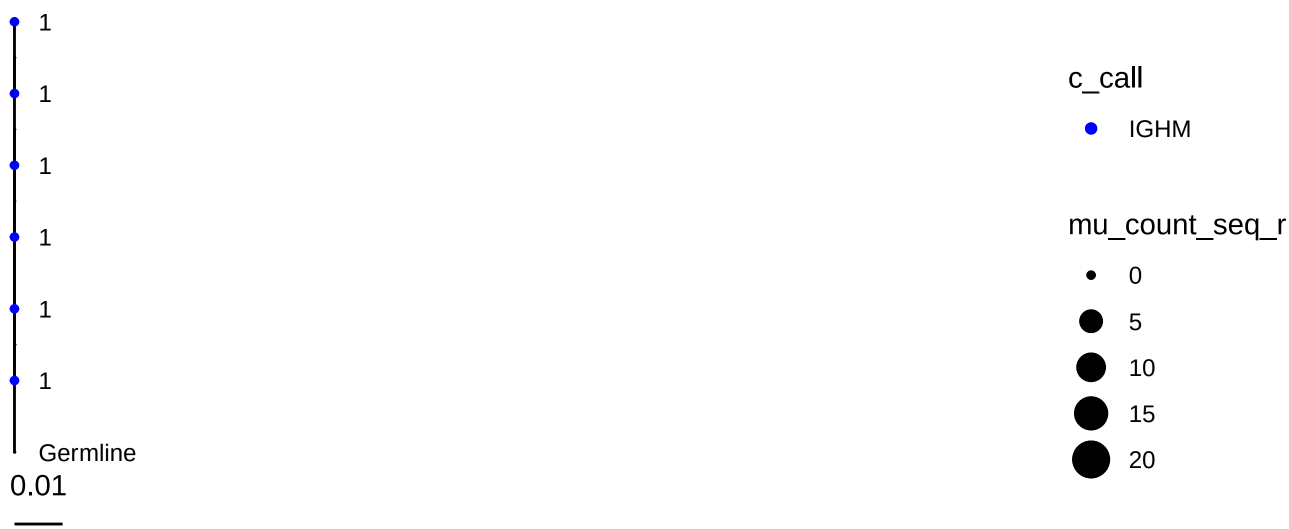
1
1
1
1
1
1
Germline
0.01
c_call
IGHM
mu_count_seq_r
0
5
10
15
20

1449

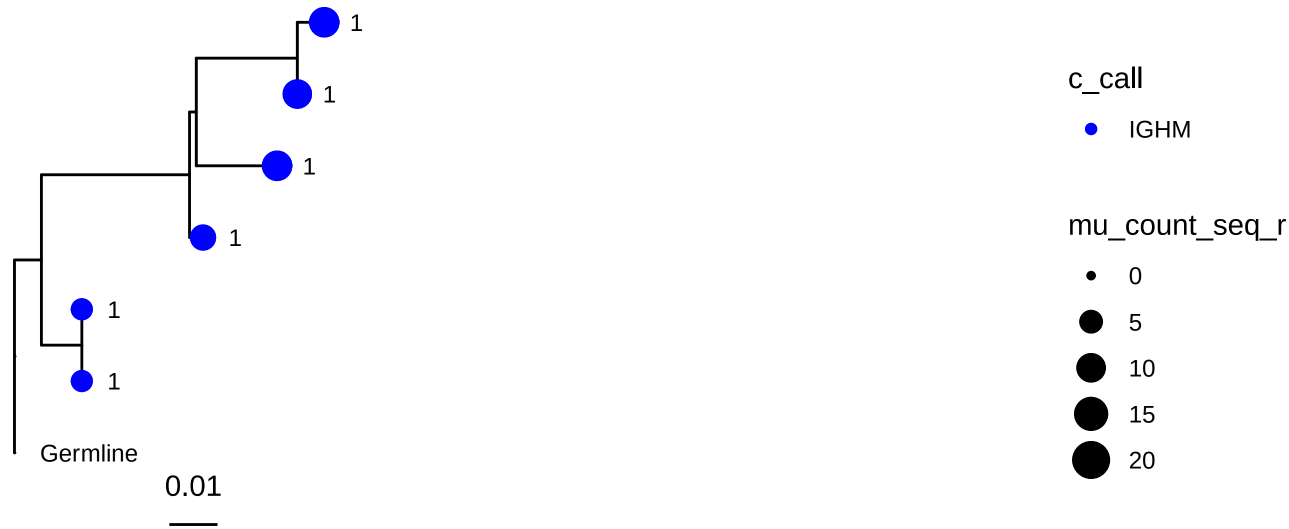
1
1
1
1
1
1
Germline
0.01
c_call
IGHM
mu_count_seq_r
0
5
10
15
20

1633

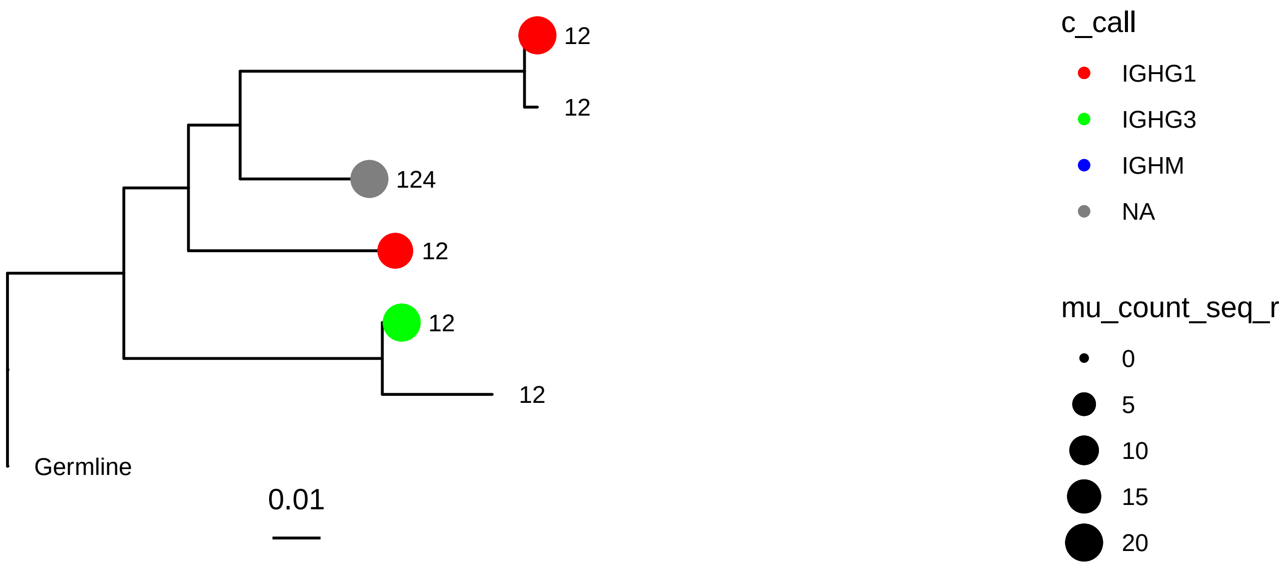
12
12
124
12
12
12
Germline
0.01
c_call
IGHG1
IGHG3
IGHM
NA
mu_count_seq_r
0
5
10
15
20

2177

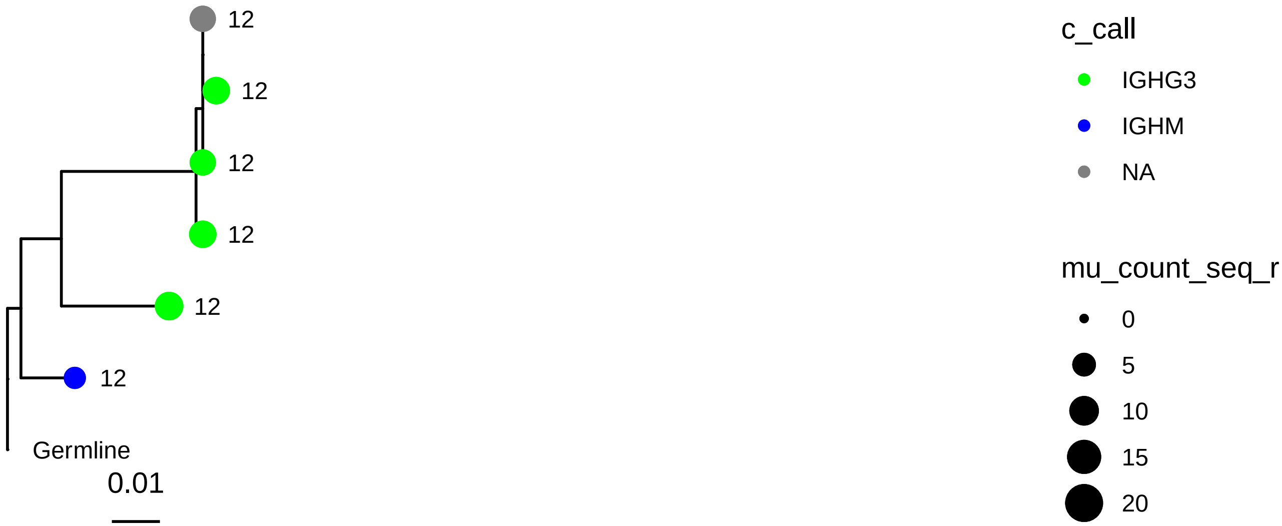
12
12
12
12
12
12
Germline
0.01
c_call
IGHG3
IGHM
NA
mu_count_seq_r
0
5
10
15
20

2240

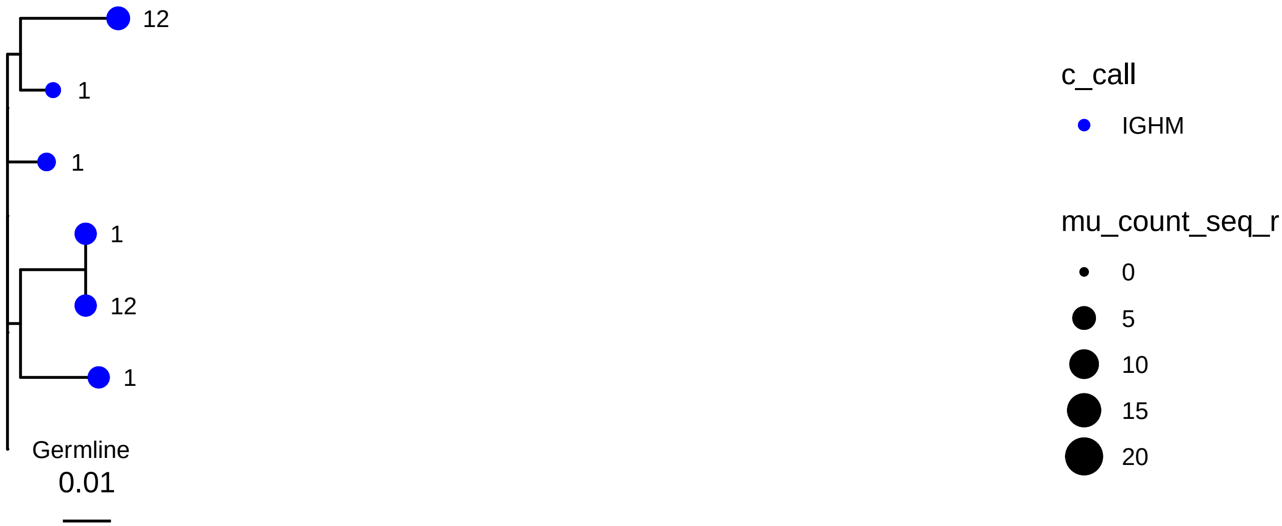
12
1
1
1
12
1
Germline
0.01
c_call
IGHM
mu_count_seq_r
0
5
10
15
20

2364

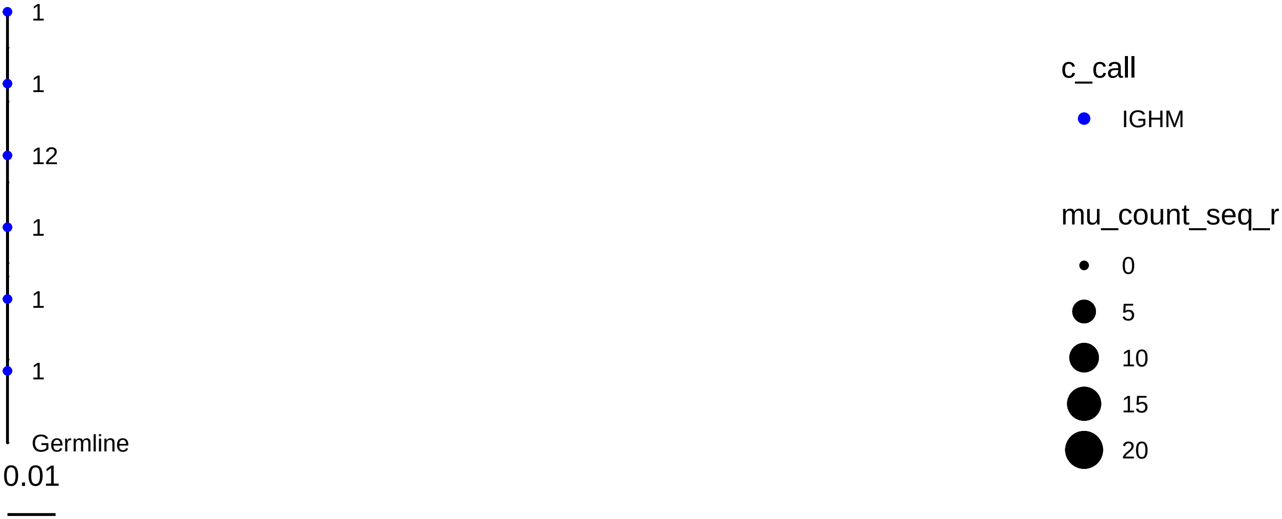
1
1
12
1
1
1
Germline
0.01
c_call
IGHM
mu_count_seq_r
0
5
10
15
20

2597

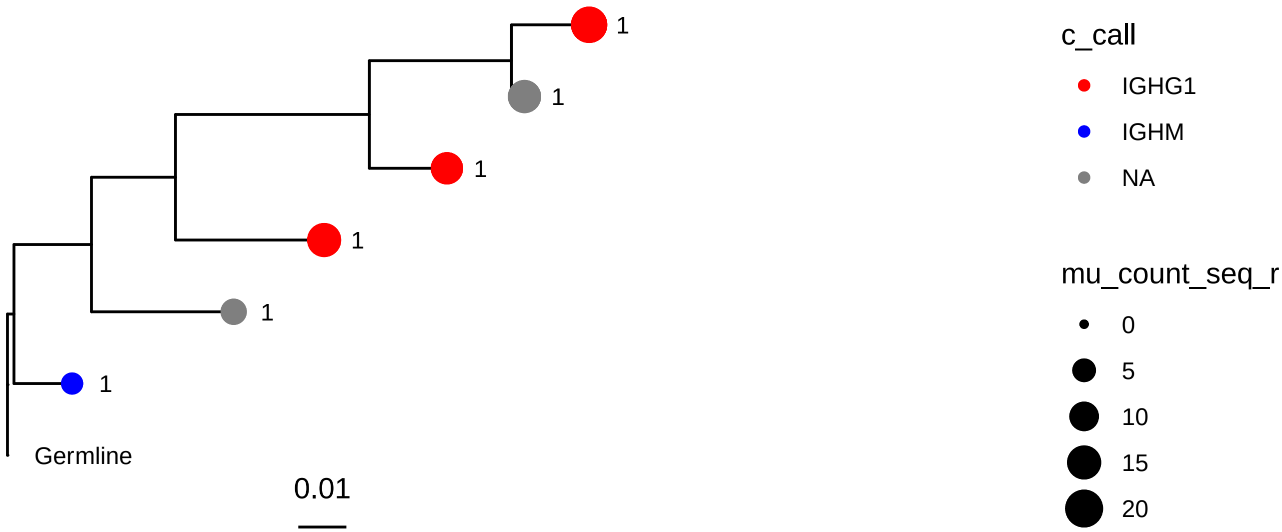
1
1
1
1
1
1
Germline
0.01
c_call
IGHG1
IGHM
NA
mu_count_seq_r
0
5
10
15
20

3393

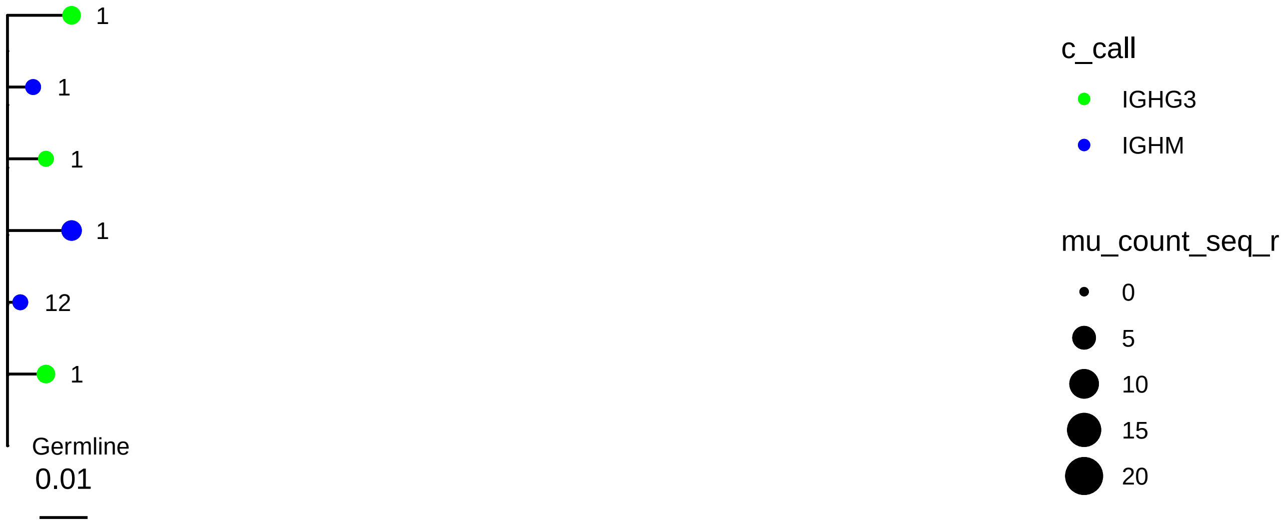
1
1
1
1
12
1
Germline
0.01
c_call
IGHG3
IGHM
mu_count_seq_r
0
5
10
15
20

3498

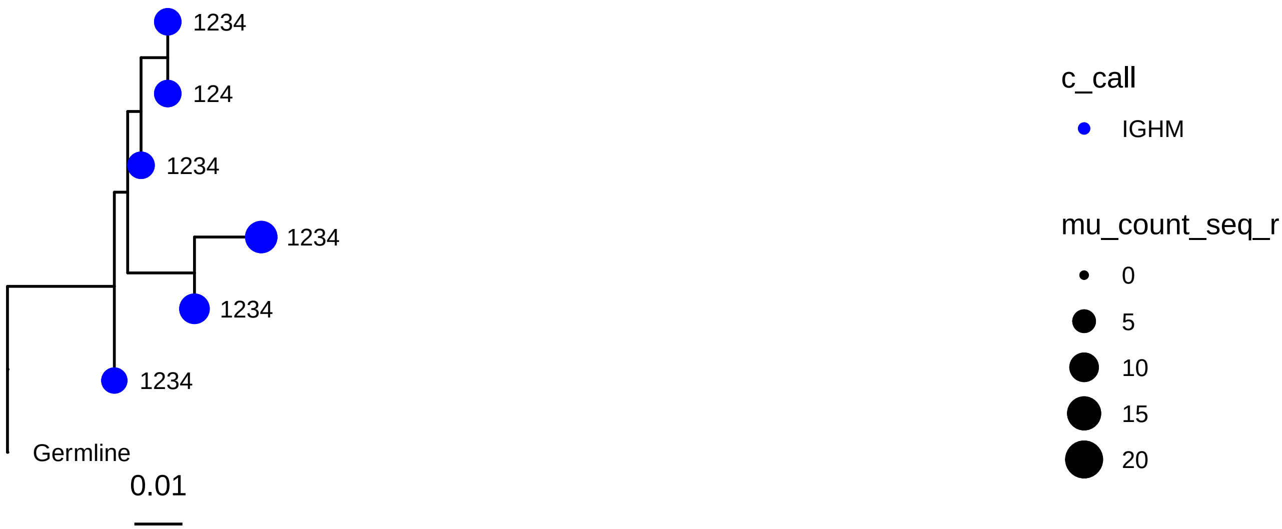
1234
124
1234
1234
1234
1234
Germline
0.01
c_call
IGHM
mu_count_seq_r
0
5
10
15
20

3607

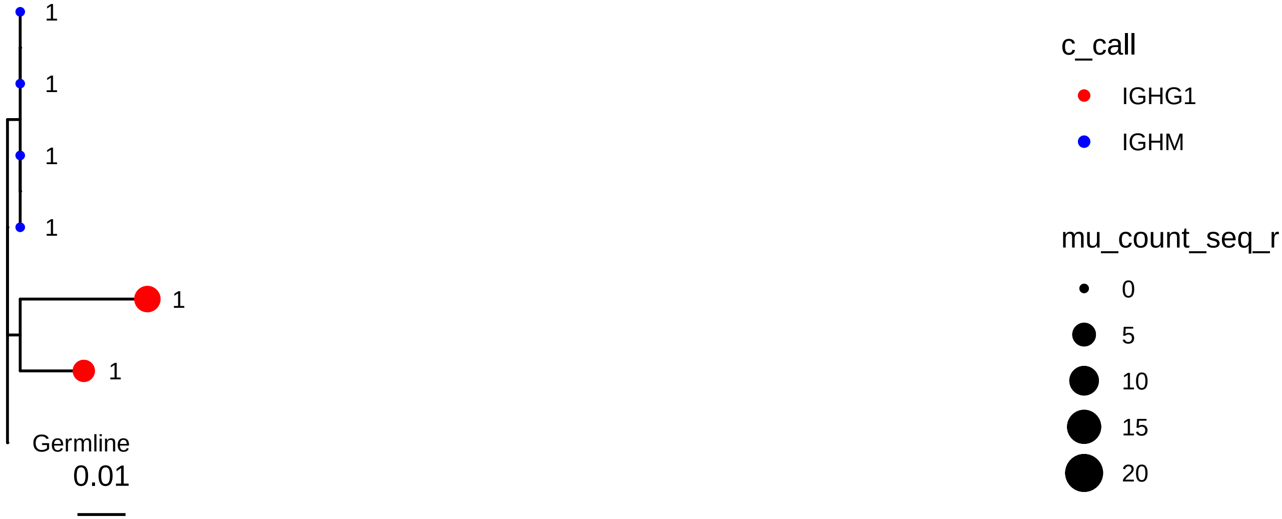
1
1
1
1
1
1
Germline
0.01
c_call
IGHG1
IGHM
mu_count_seq_r
0
5
10
15
20

3689

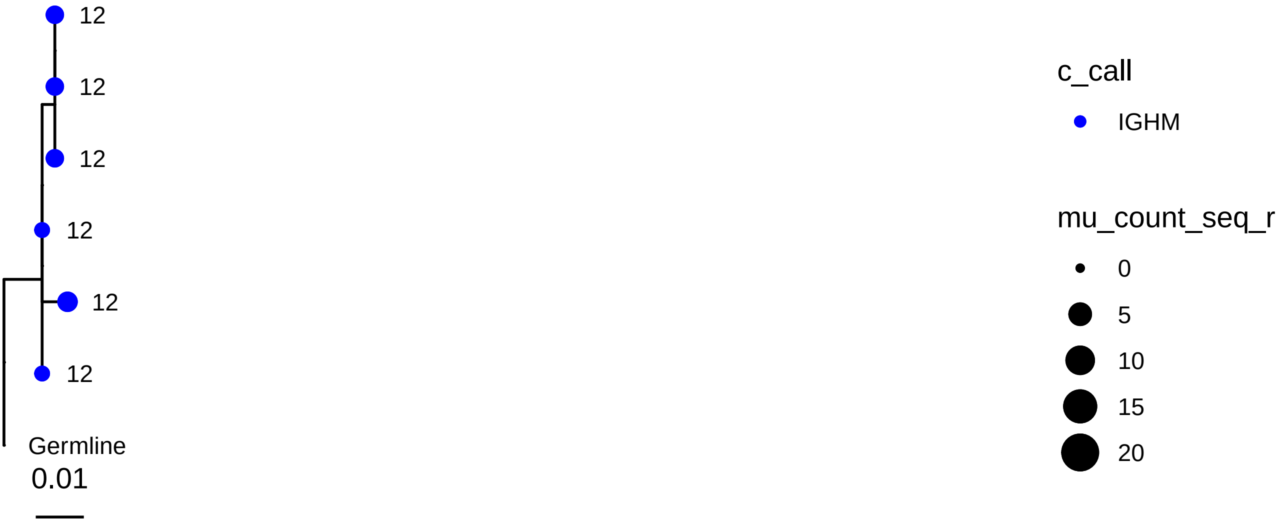
12
12
12
12
12
12
Germline
0.01
c_call
IGHM
mu_count_seq_r
0
5
10
15
20

3872

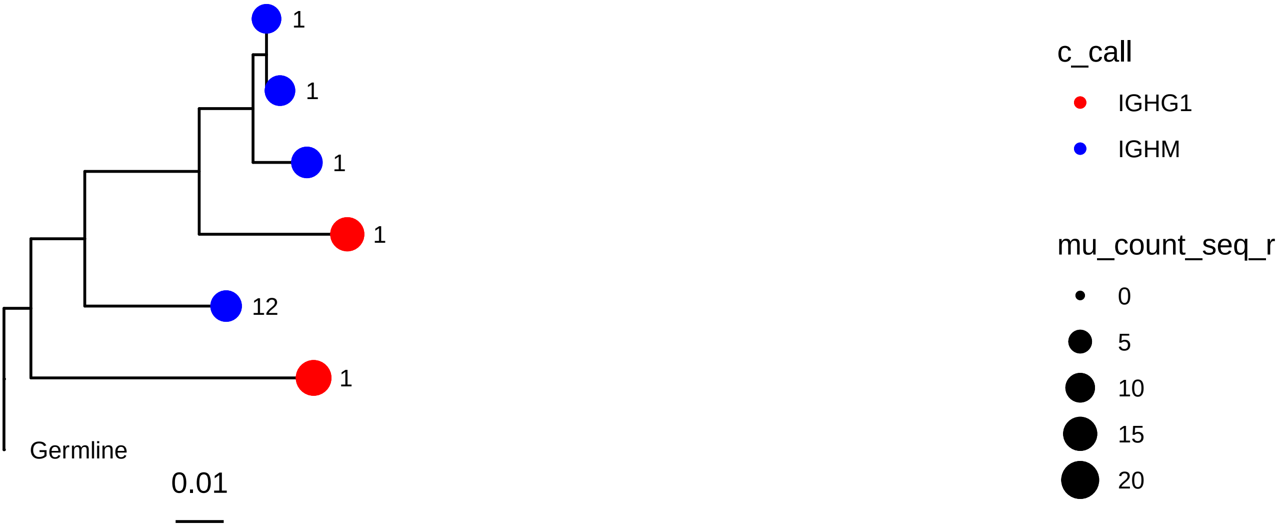
1
1
1
1
12
1
Germline
0.01
c_call
IGHG1
IGHM
mu_count_seq_r
0
5
10
15
20

4117

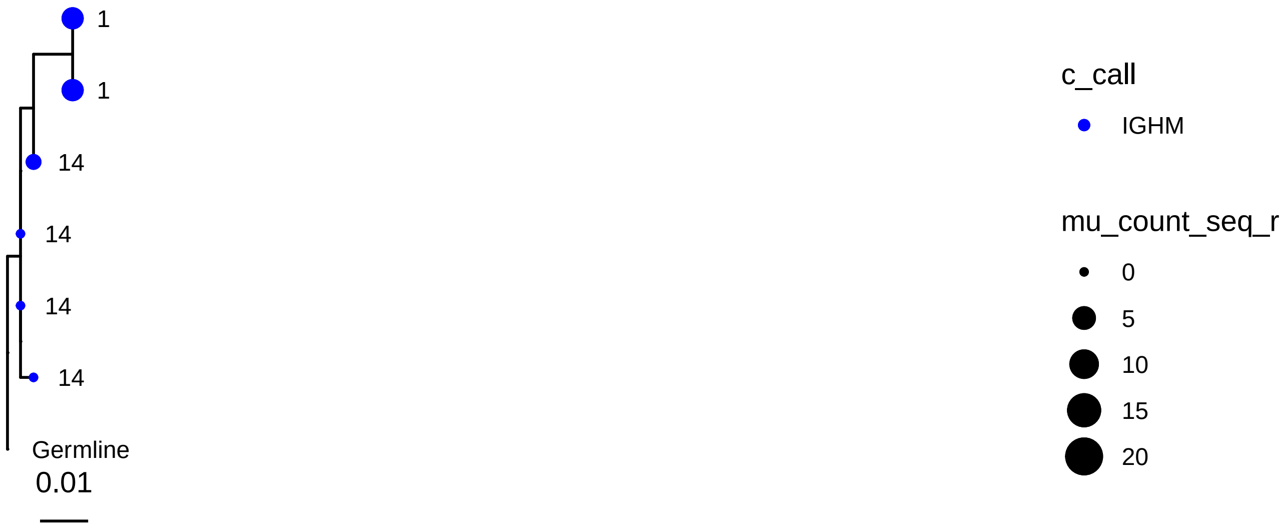
1
1
14
14
14
14
Germline
0.01
c_call
IGHM
mu_count_seq_r
0
5
10
15
20

4196

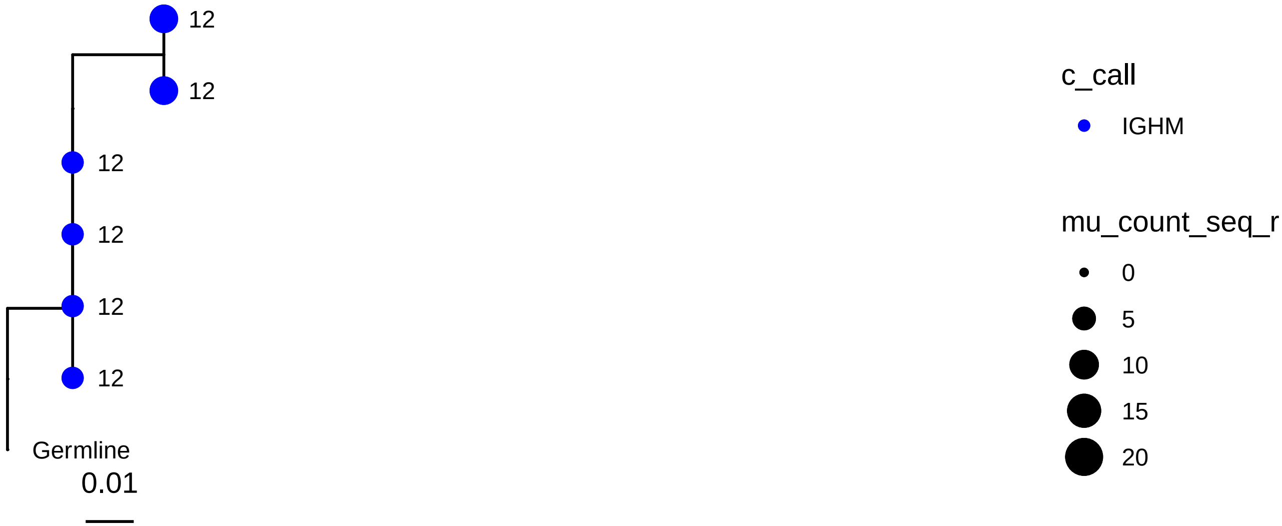
12
12
12
12
12
12
Germline
0.01
c_call
IGHM
mu_count_seq_r
0
5
10
15
20

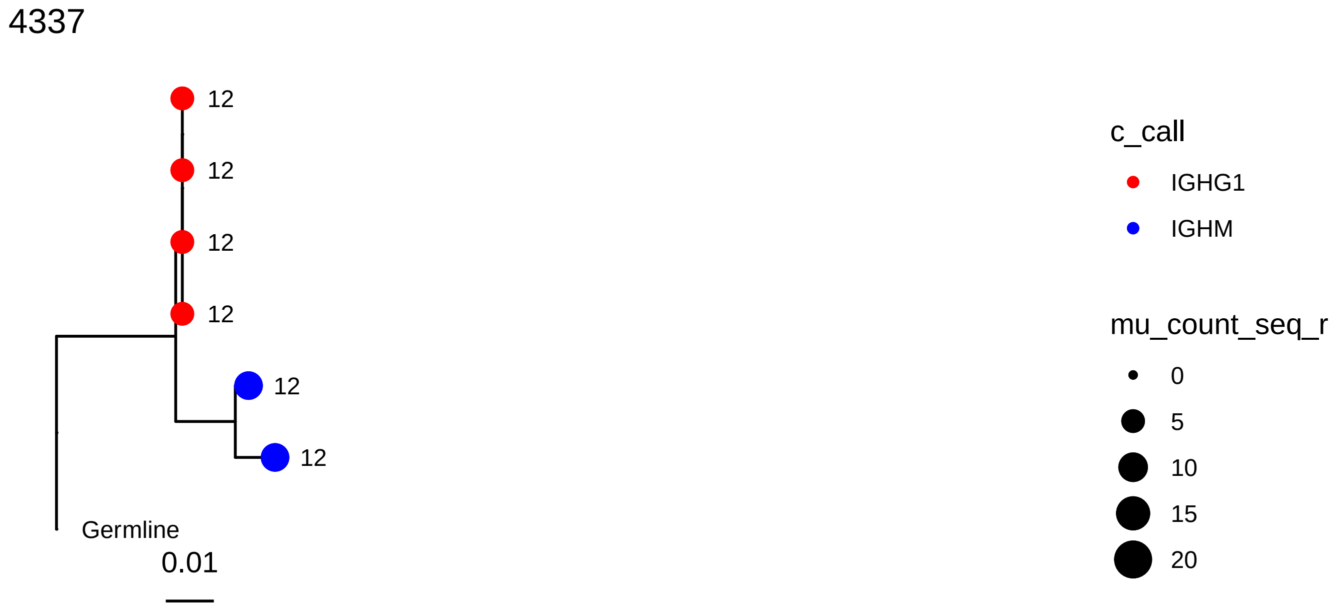
4337
12
12
12
12
12
12
Germline
0.01
c_call
IGHG1
IGHM
mu_count_seq_r
0
5
10
15
20

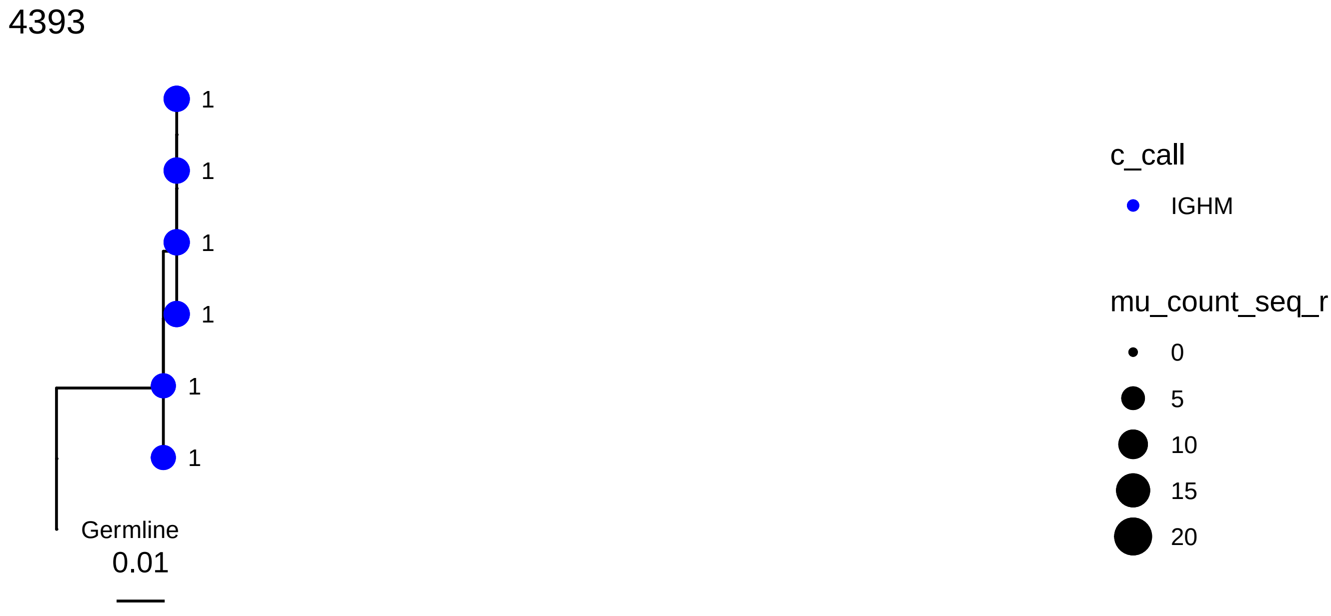
4393
1
1
1
1
1
1
Germline
0.01
c_call
IGHM
mu_count_seq_r
0
5
10
15
20

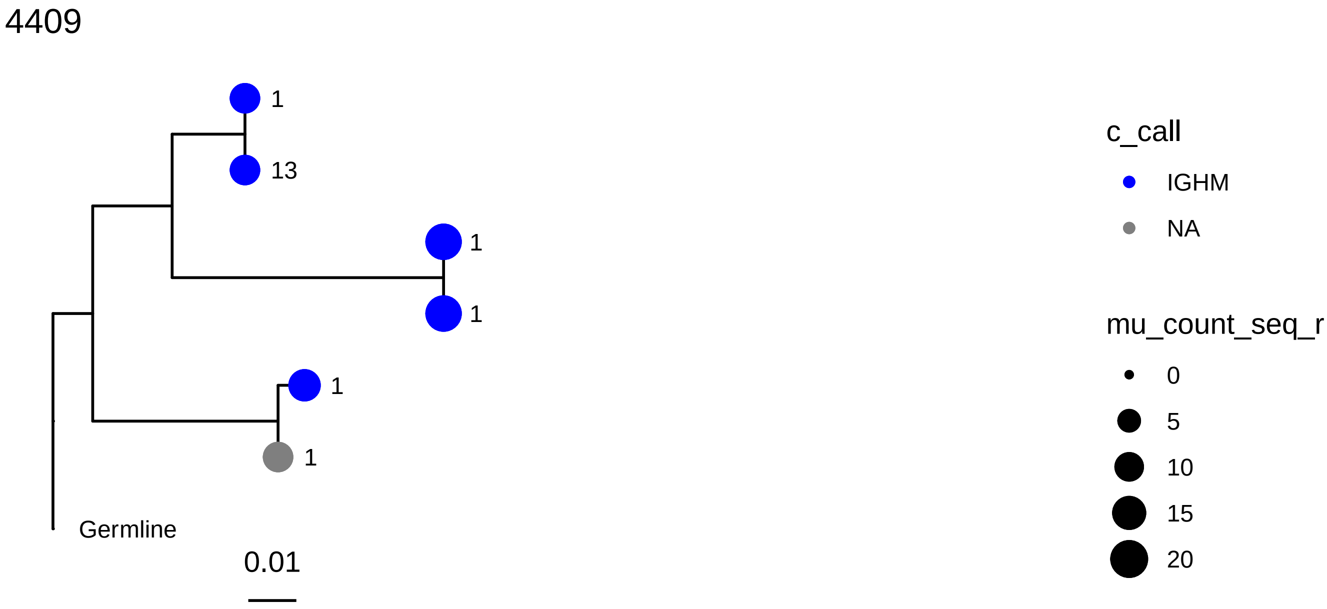
4409
1
13
1
1
1
1
Germline
0.01
c_call
IGHM
NA
mu_count_seq_r
0
5
10
15
20

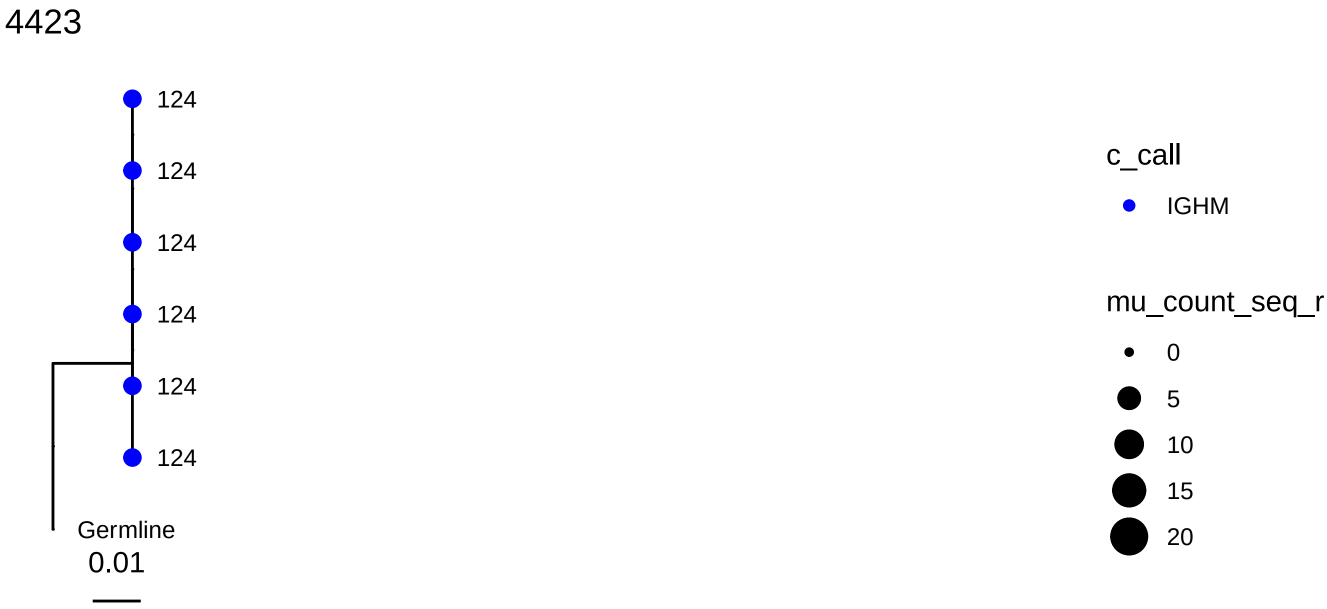
4423
124
124
124
124
124
124
Germline
0.01
c_call
IGHM
mu_count_seq_r
0
5
10
15
20

4663

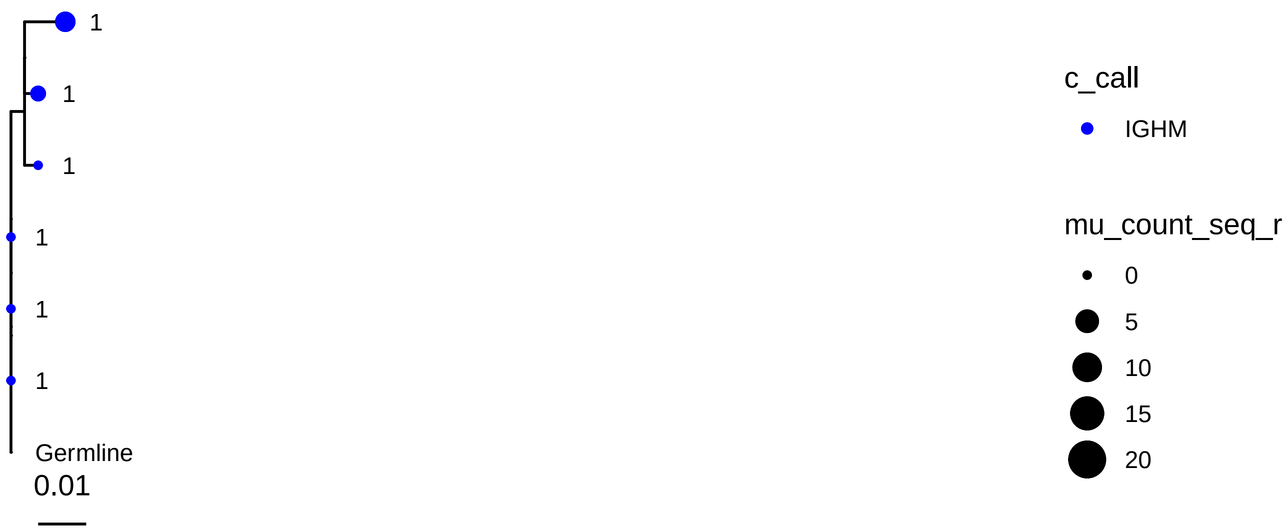
1
1
1
1
1
1
Germline
0.01
c_call
IGHM
mu_count_seq_r
0
5
10
15
20

4956

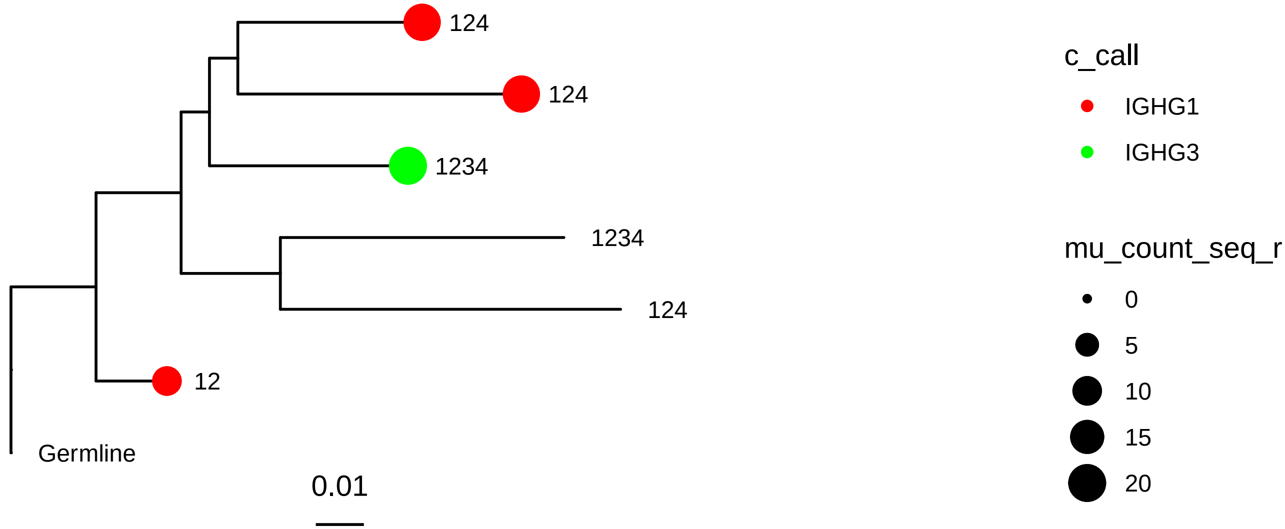
124
124
1234
1234
124
12
Germline
0.01
c_call
IGHG1
IGHG3
mu_count_seq_r
0
5
10
15
20

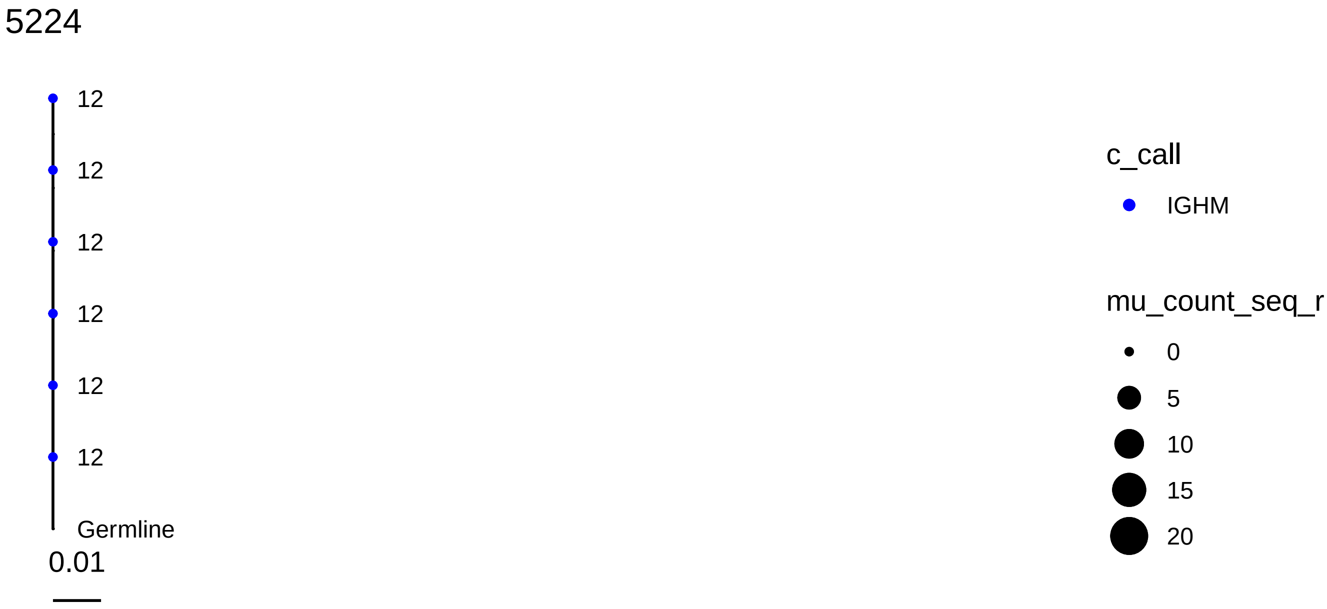
5224
12
12
12
12
12
12
Germline
0.01
c_call
IGHM
mu_count_seq_r
0
5
10
15
20

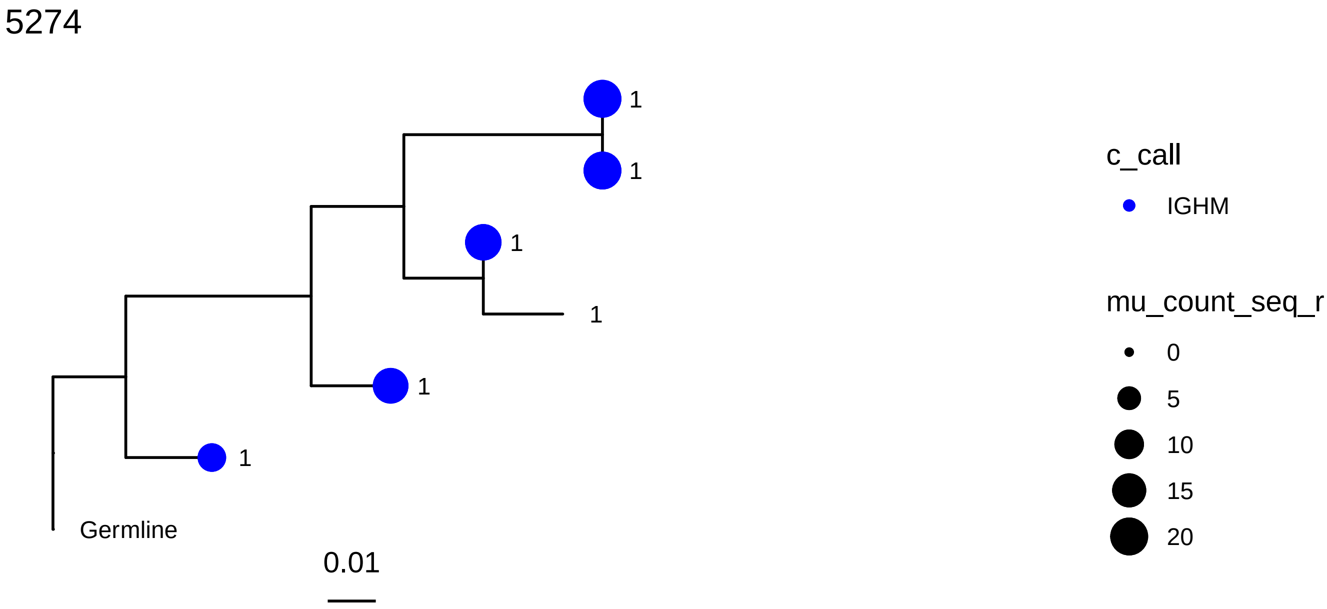
5274
1
1
1
1
1
1
Germline
0.01
c_call
IGHM
mu_count_seq_r
0
5
10
15
20

202

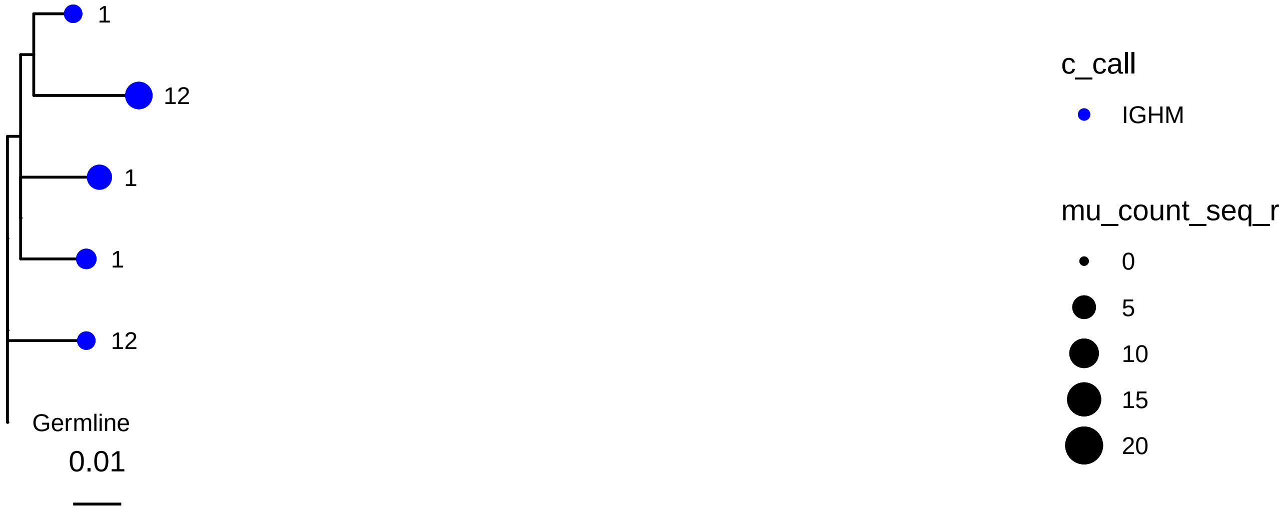
1
12
1
1
12
Germline
0.01
c_call
IGHM
mu_count_seq_r
0
5
10
15
20

264

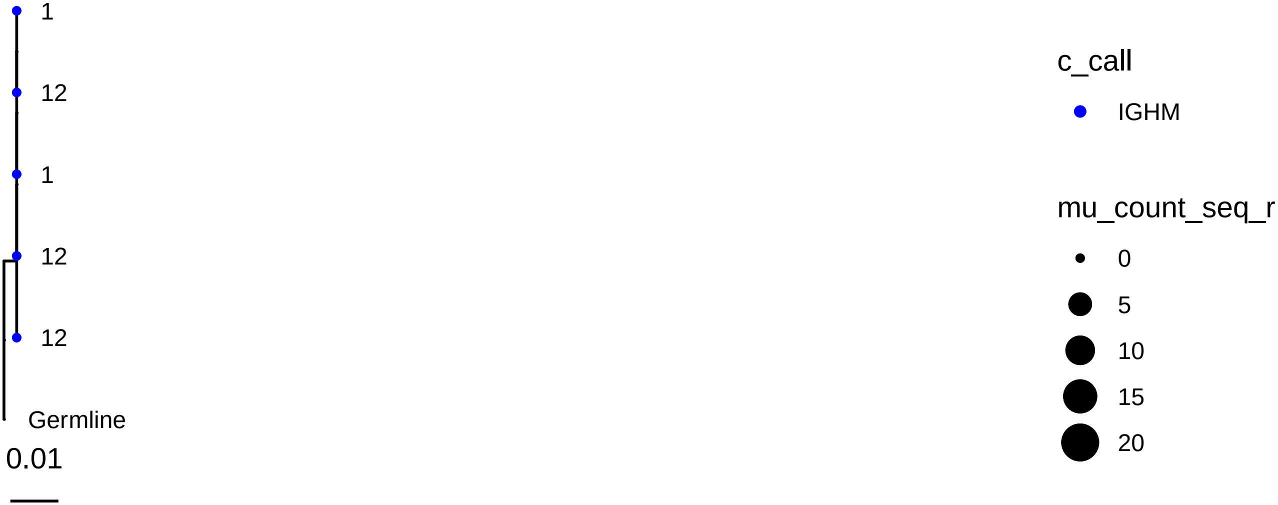
1
12
1
12
12
Germline
0.01
c_call
IGHM
mu_count_seq_r
0
5
10
15
20

433

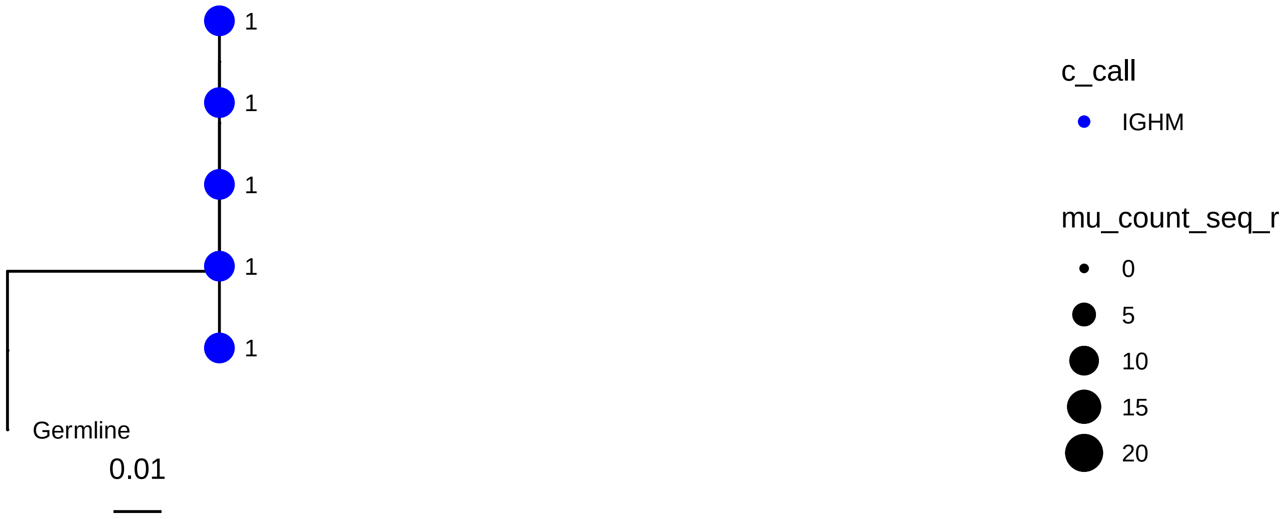
1
1
1
1
1
Germline
0.01
c_call
IGHM
mu_count_seq_r
0
5
10
15
20

504

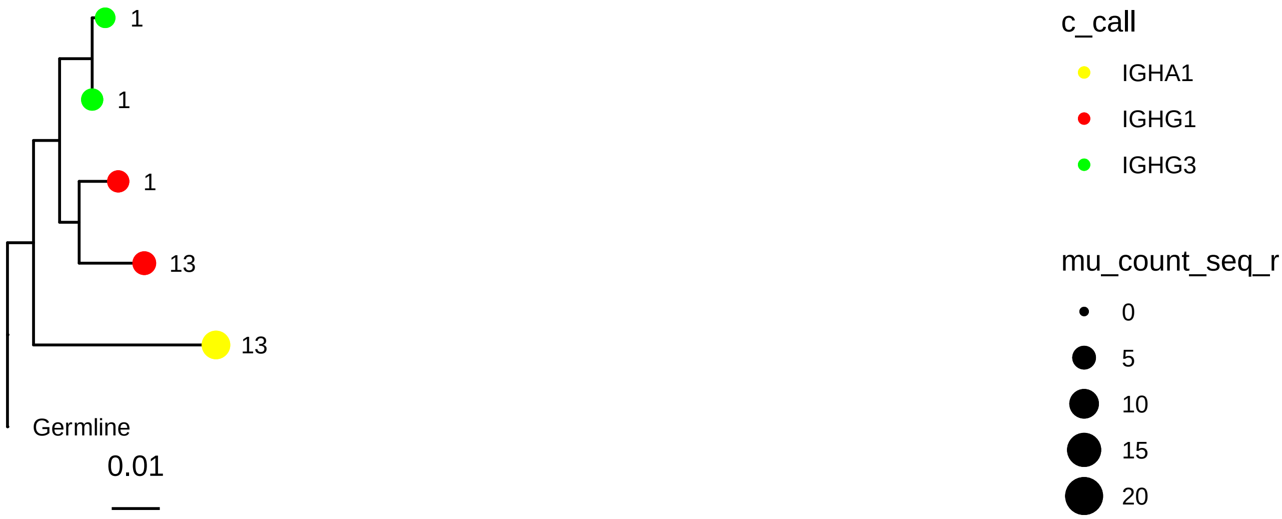
1
1
1
13
13
Germline
0.01
c_call
IGHA1
IGHG1
IGHG3
mu_count_seq_r
0
5
10
15
20

506

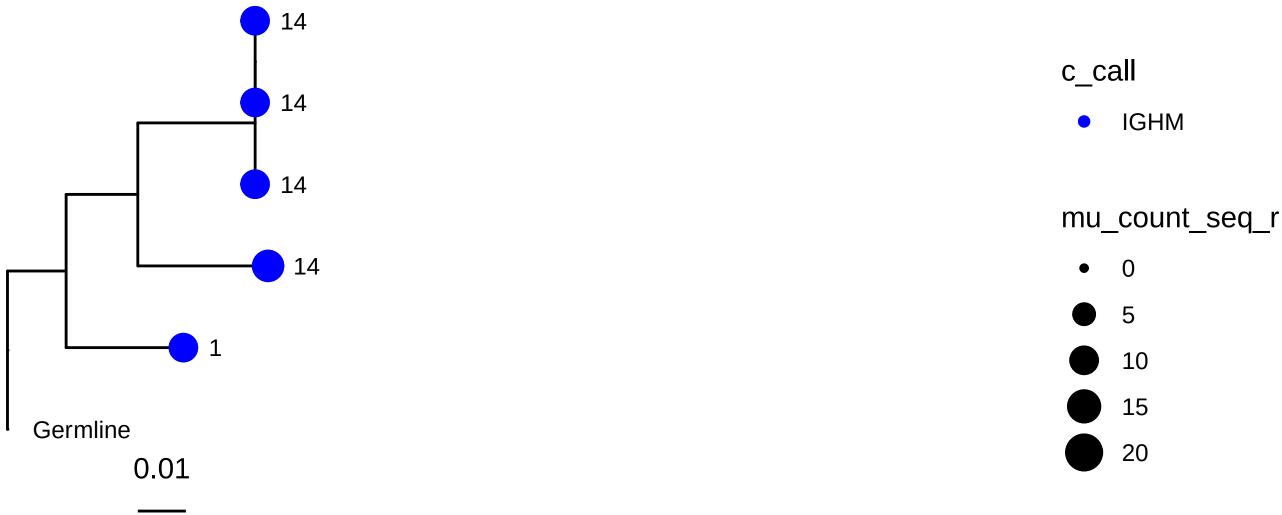

14
14
14
14
1
Germline
0.01
c_call
IGHM
mu_count_seq_r
0
5
10
15
20


746

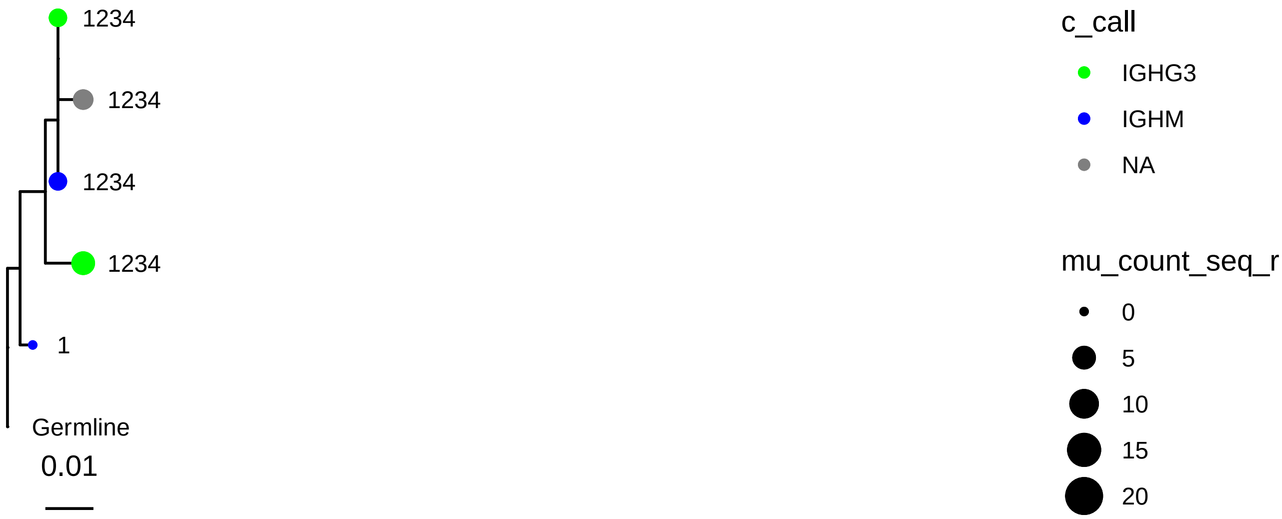

1234
1234
1234
1234
1
Germline
0.01
c_call
IGHG3
IGHM
NA
mu_count_seq_r
0
5
10
15
20

800

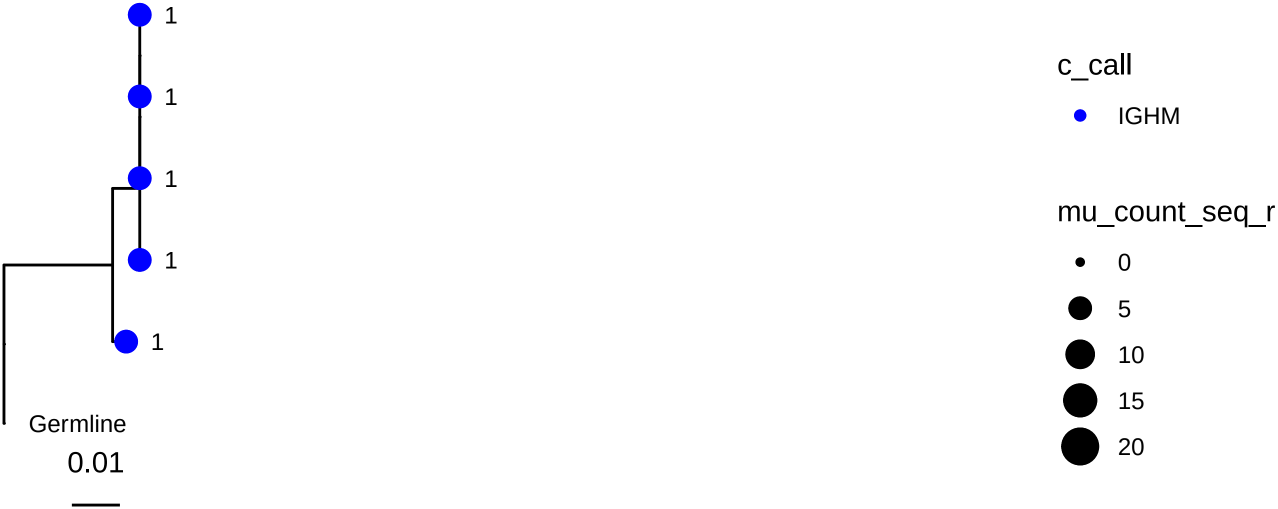
1
1
1
1
1
Germline
0.01
c_call
IGHM
mu_count_seq_r
0
5
10
15
20

936

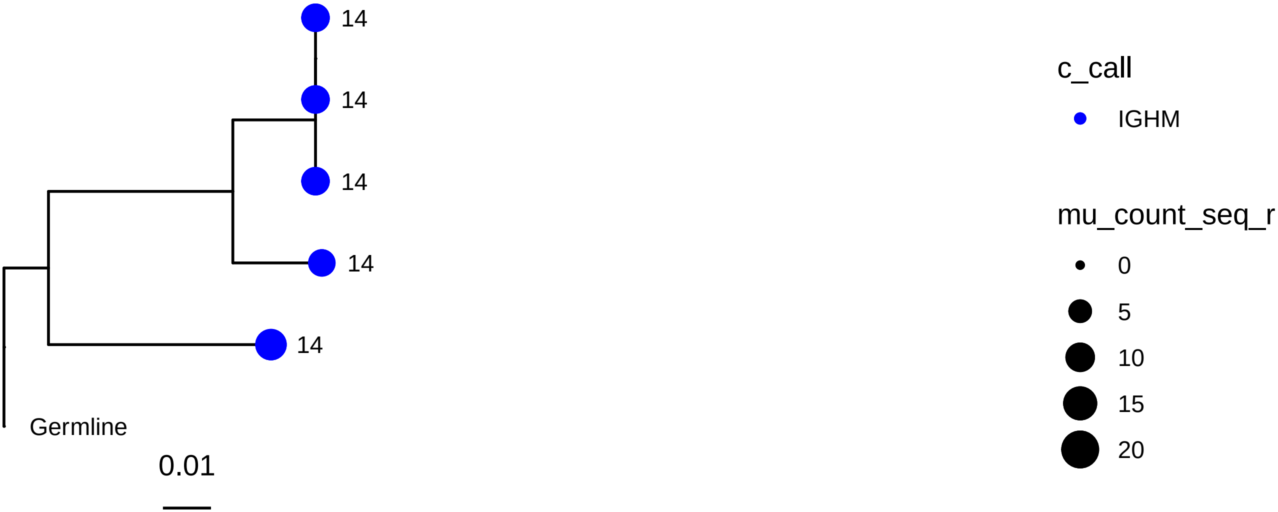
14
14
14
14
14
Germline
0.01
c_call
IGHM
mu_count_seq_r
0
5
10
15
20

1114

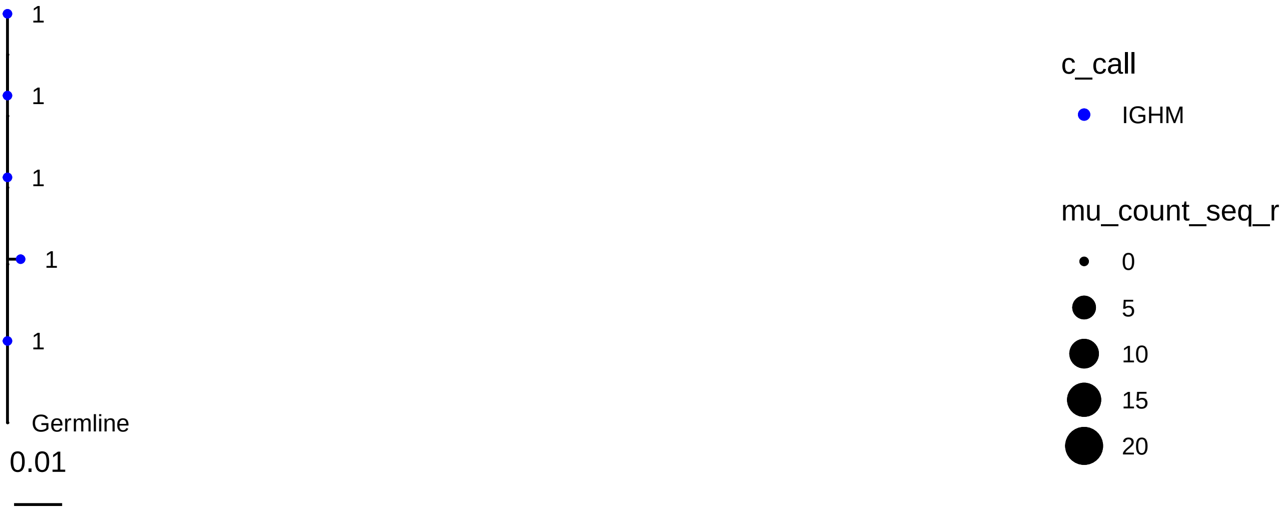
1
1
1
1
1
Germline
0.01
c_call
IGHM
mu_count_seq_r
0
5
10
15
20

1419

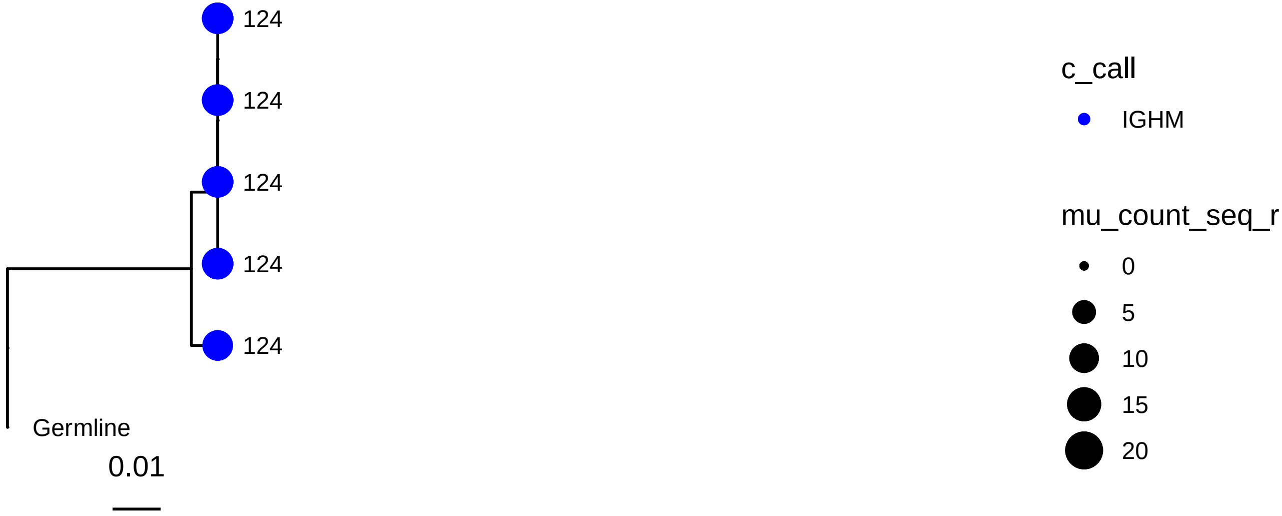
124
124
124
124
124
Germline
0.01
c_call
IGHM
mu_count_seq_r
0
5
10
15
20

1648

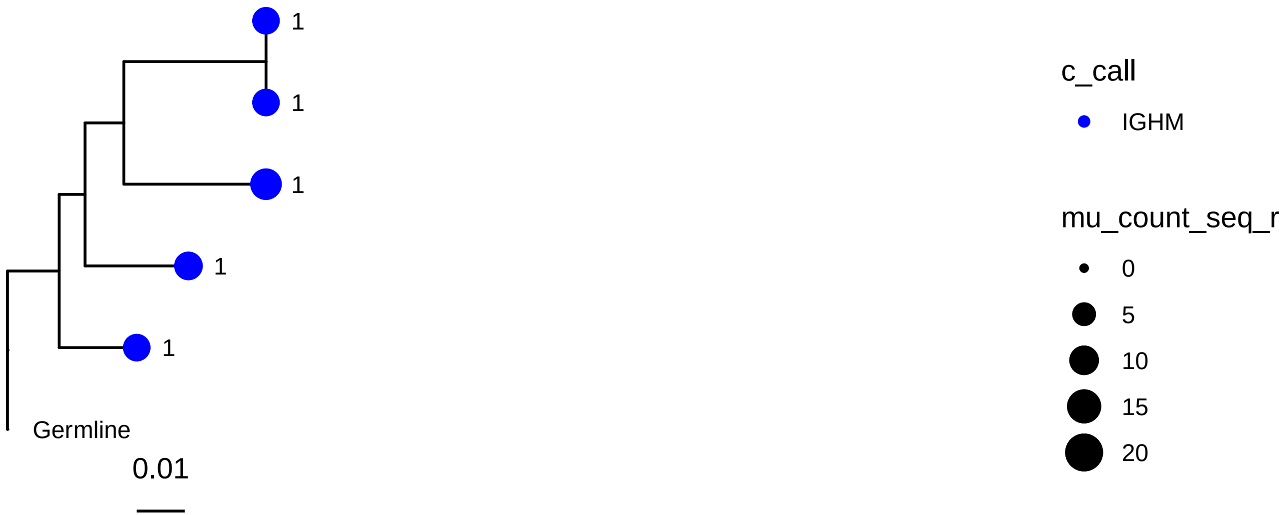
1
1
1
1
1
Germline
0.01
c_call
IGHM
mu_count_seq_r
0
5
10
15
20

1699

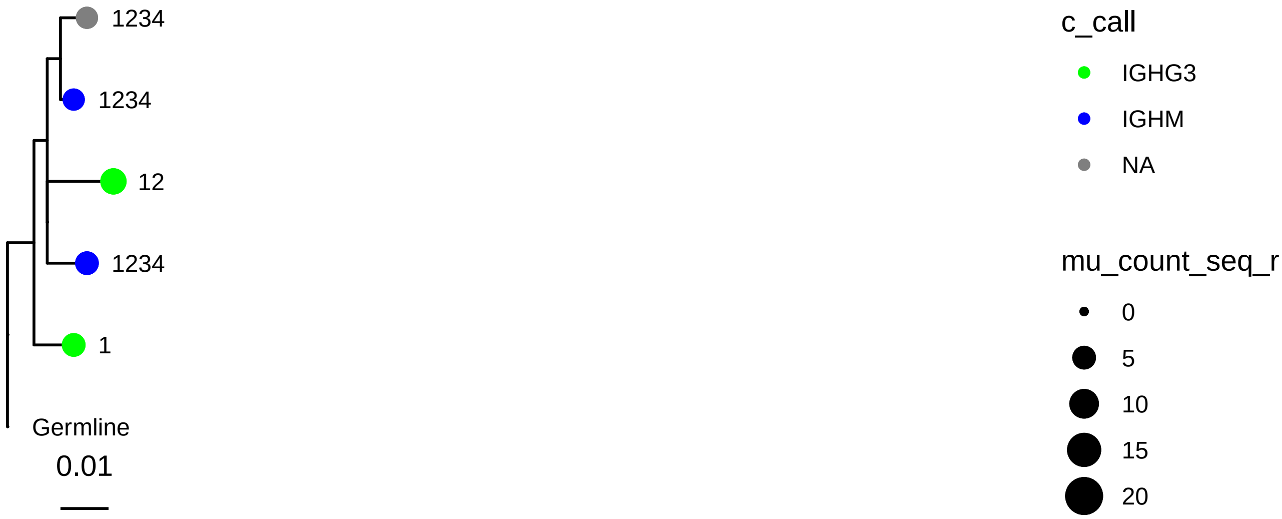
1234
1234
12
1234
1
Germline
0.01
c_call
IGHG3
IGHM
NA
mu_count_seq_r
0
5
10
15
20

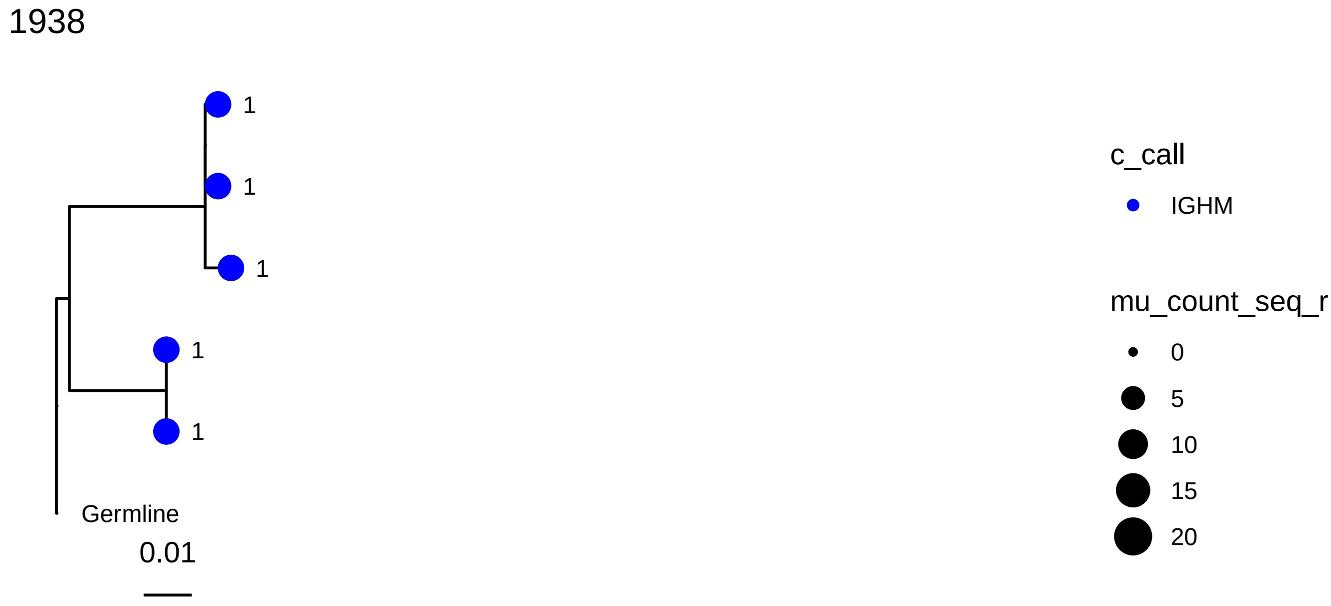
1938
1
1
1
1
1
Germline
0.01
c_call
IGHM
mu_count_seq_r
0
5
10
15
20

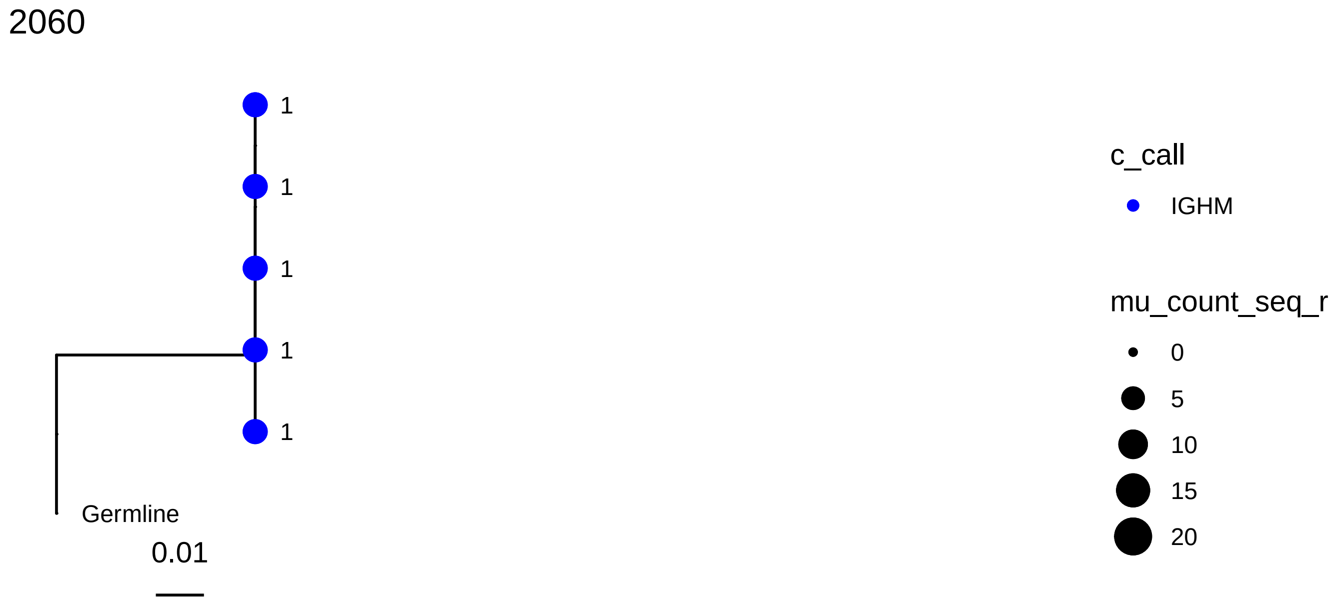
2060
1
1
1
1
1
Germline
0.01
c_call
IGHM
mu_count_seq_r
0
5
10
15
20

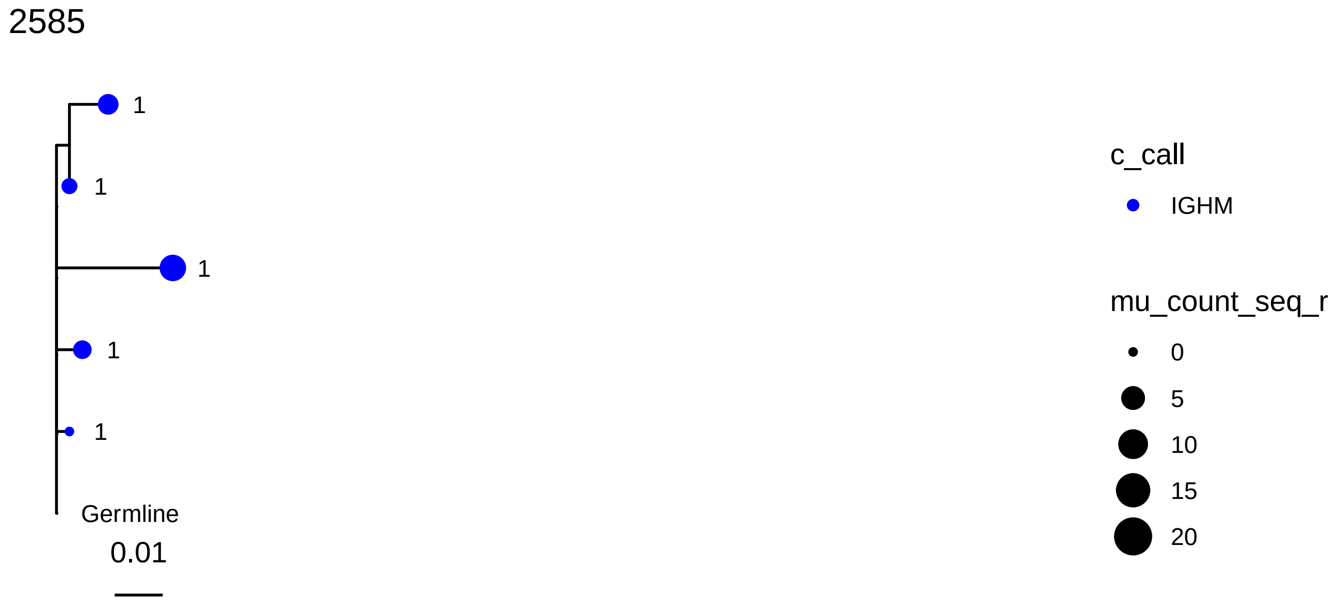
2585
1
1
1
1
1
Germline
0.01
c_call
IGHM
mu_count_seq_r
0
5
10
15
20

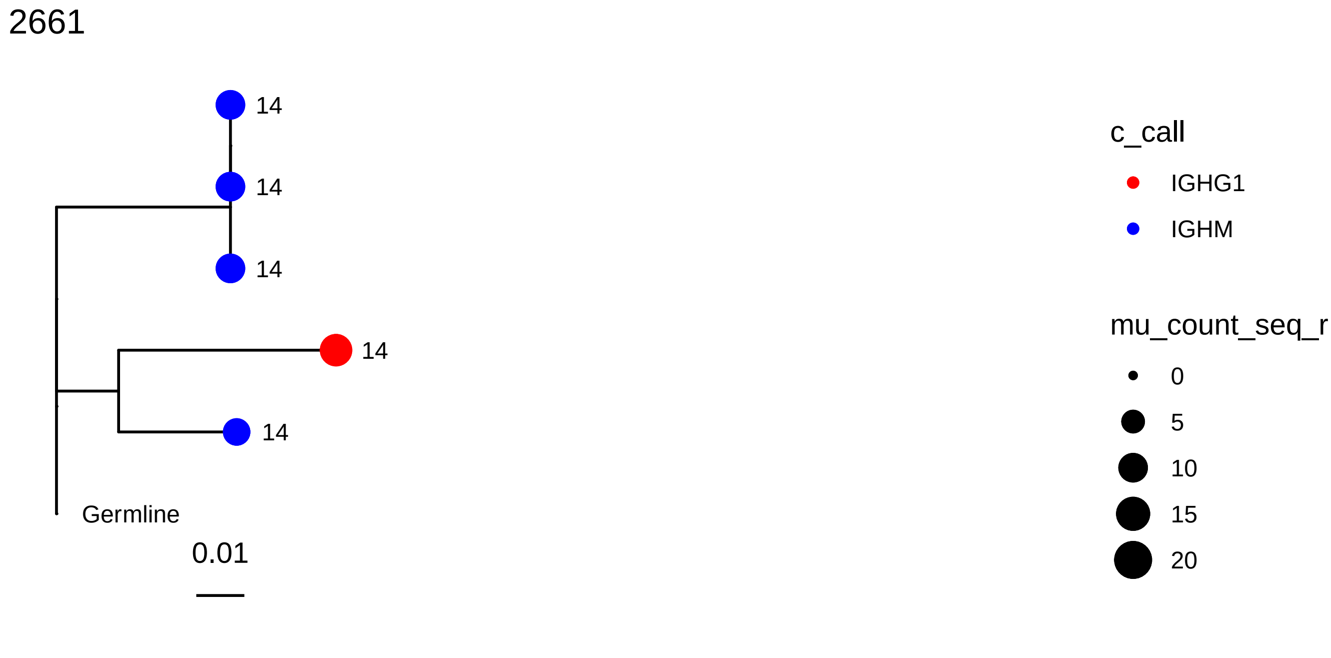
2661
14
14
14
14
14
Germline
0.01
c_call
IGHG1
IGHM
mu_count_seq_r
0
5
10
15
20

2741

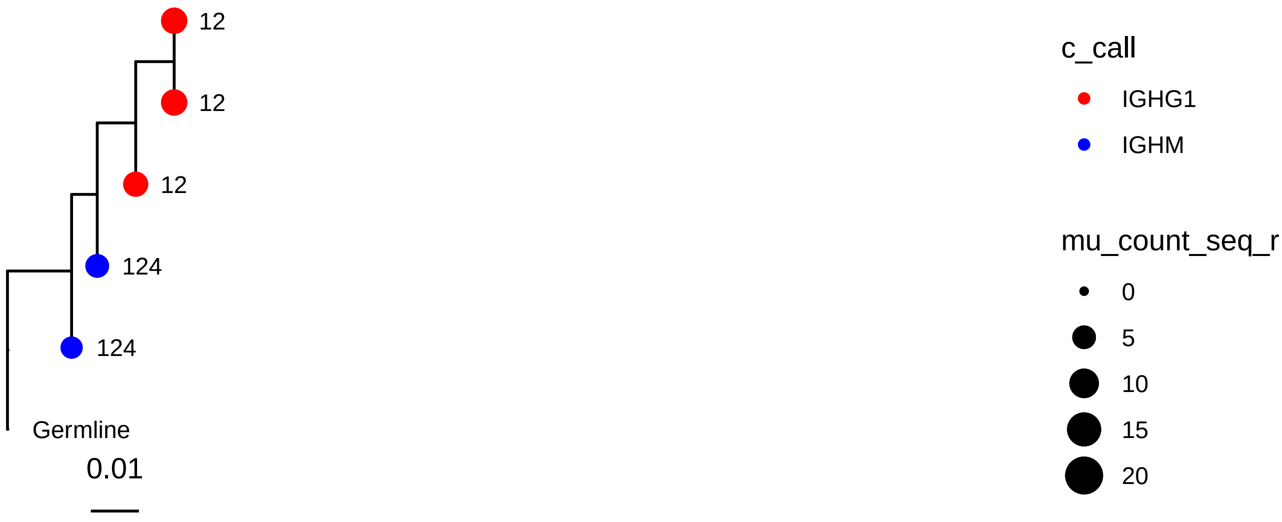
12
12
12
124
124
Germline
0.01
c_call
IGHG1
IGHM
mu_count_seq_r
0
5
10
15
20

2766

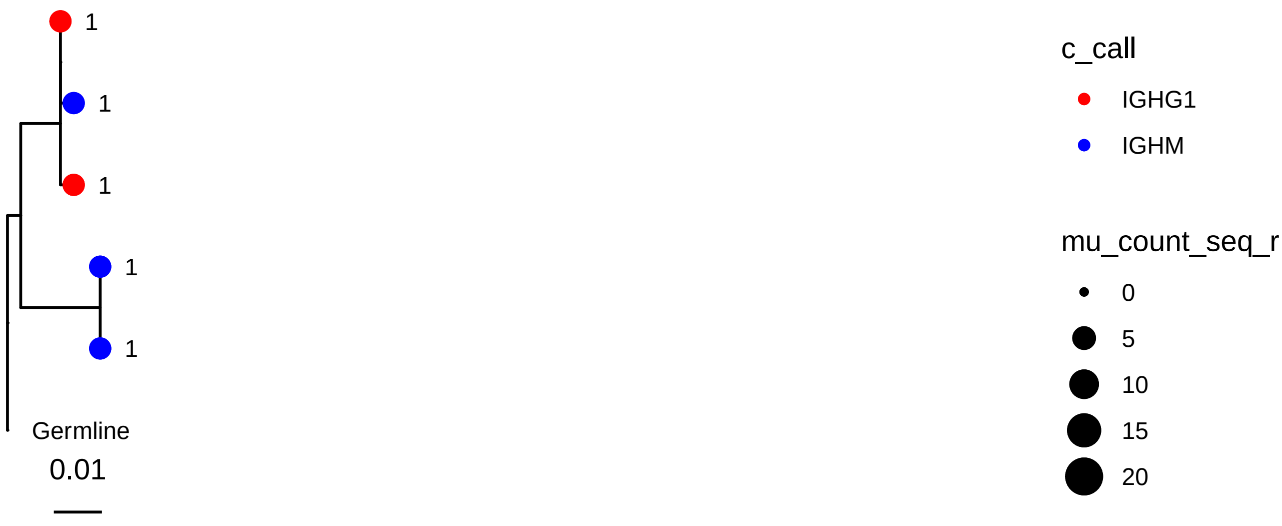
1
1
1
1
1
Germline
0.01
c_call
IGHG1
IGHM
mu_count_seq_r
0
5
10
15
20

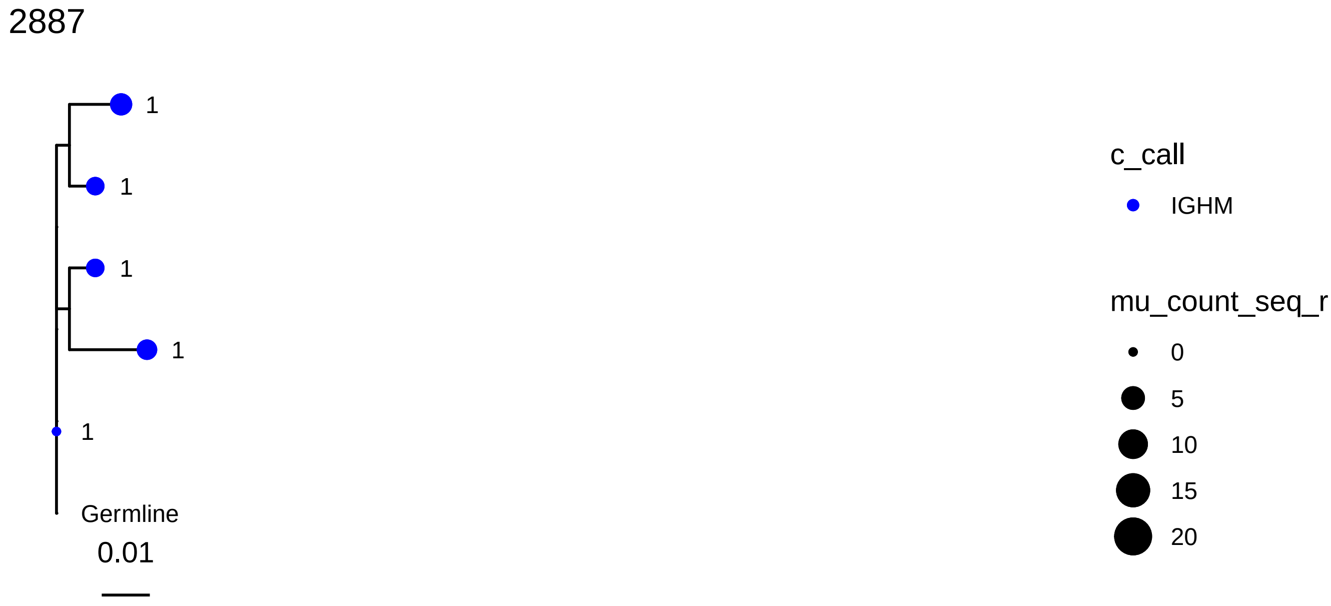
2887
1
1
1
1
1
Germline
0.01
c_call
IGHM
mu_count_seq_r
0
5
10
15
20

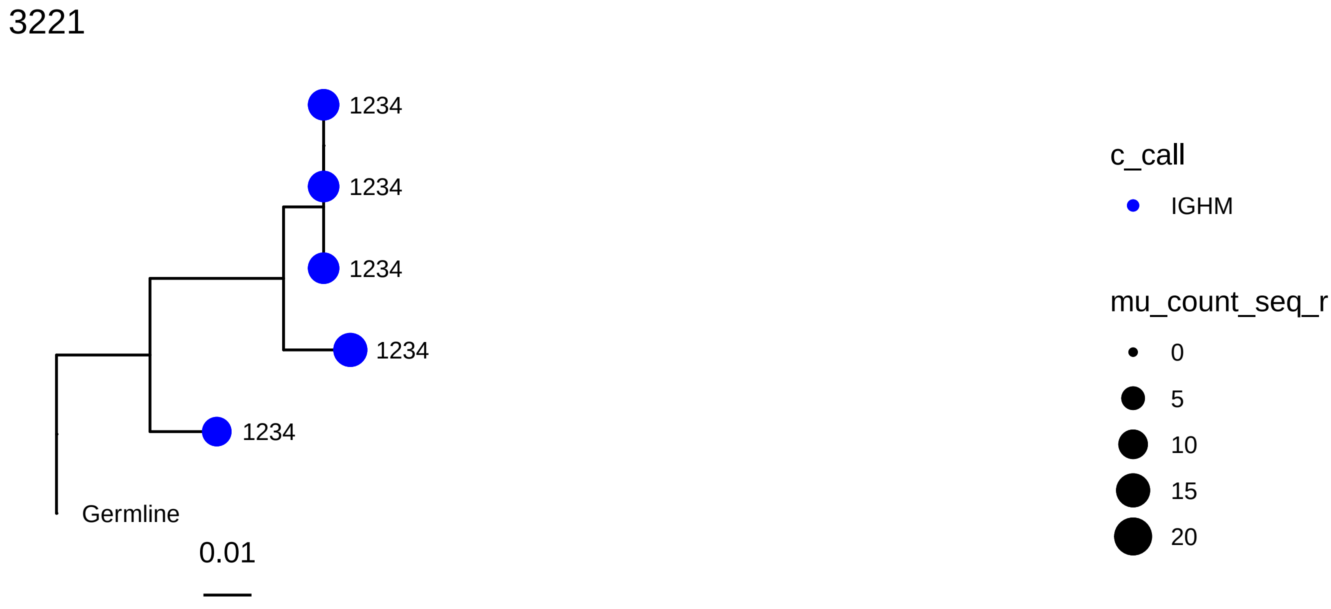
3221
1234
1234
1234
1234
1234
Germline
0.01
c_call
IGHM
mu_count_seq_r
0
5
10
15
20

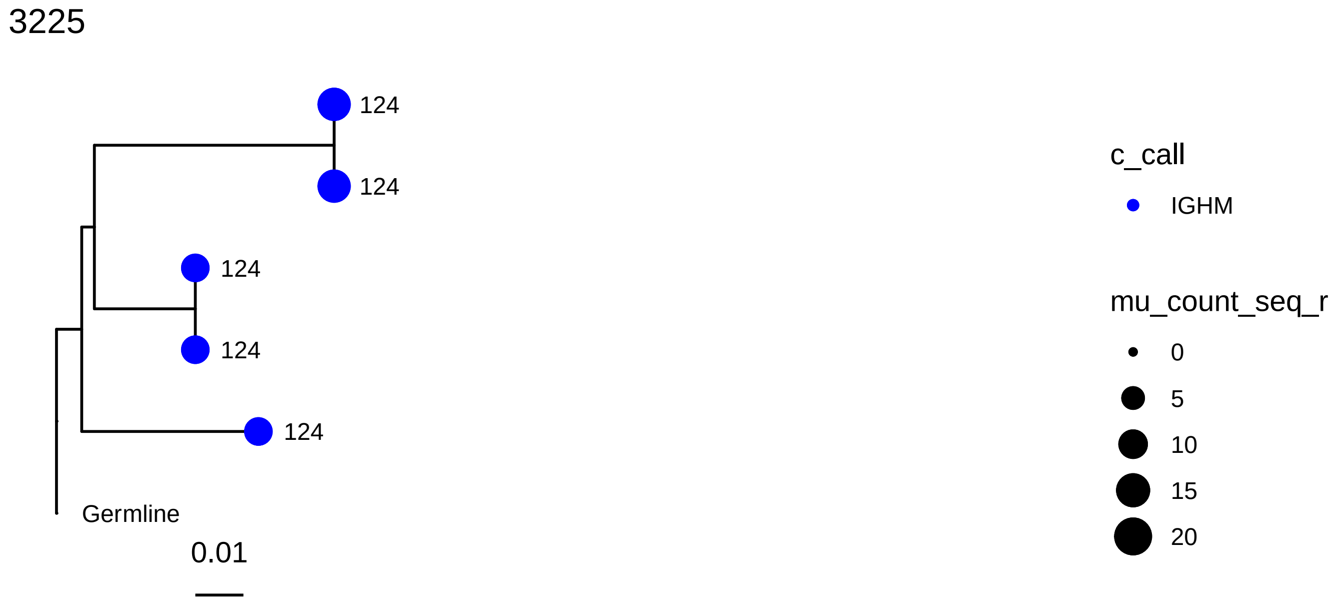
3225
124
124
124
124
124
Germline
0.01
c_call
IGHM
mu_count_seq_r
0
5
10
15
20

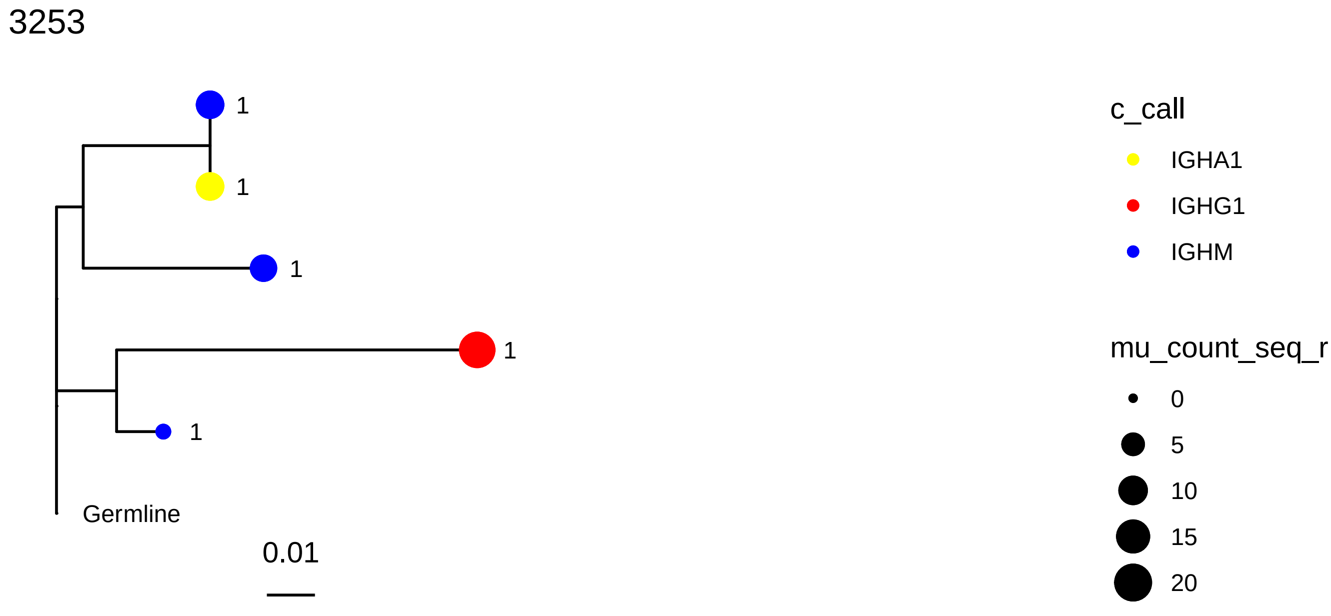
3253
1
1
1
1
1
Germline
0.01
c_call
IGHA1
IGHG1
IGHM
mu_count_seq_r
0
5
10
15
20

3427

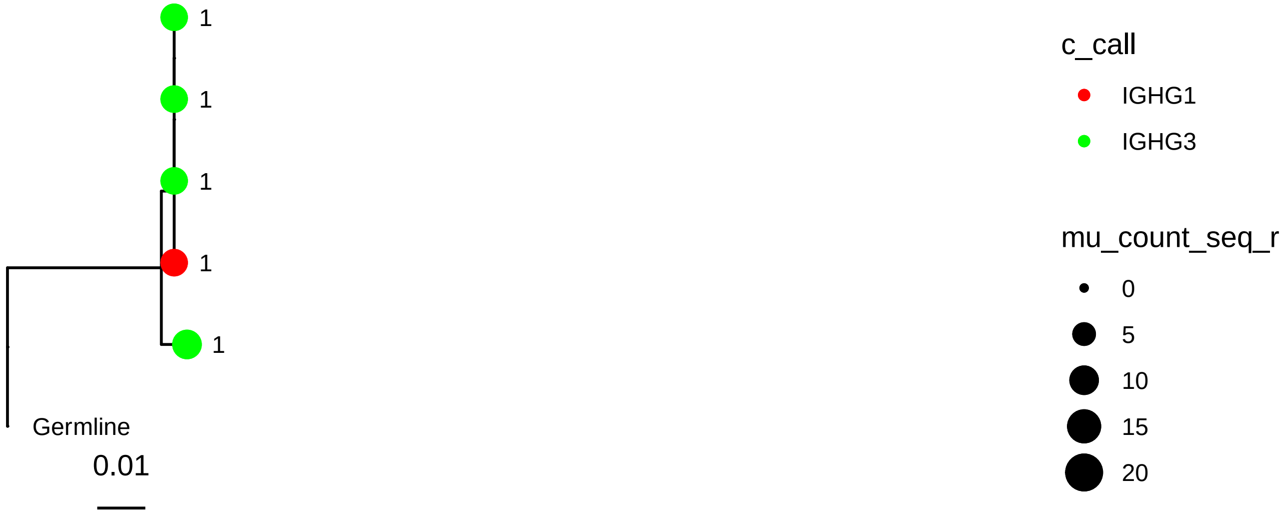
1
1
1
1
1
Germline
0.01
c_call
IGHG1
IGHG3
mu_count_seq_r
0
5
10
15
20

3569

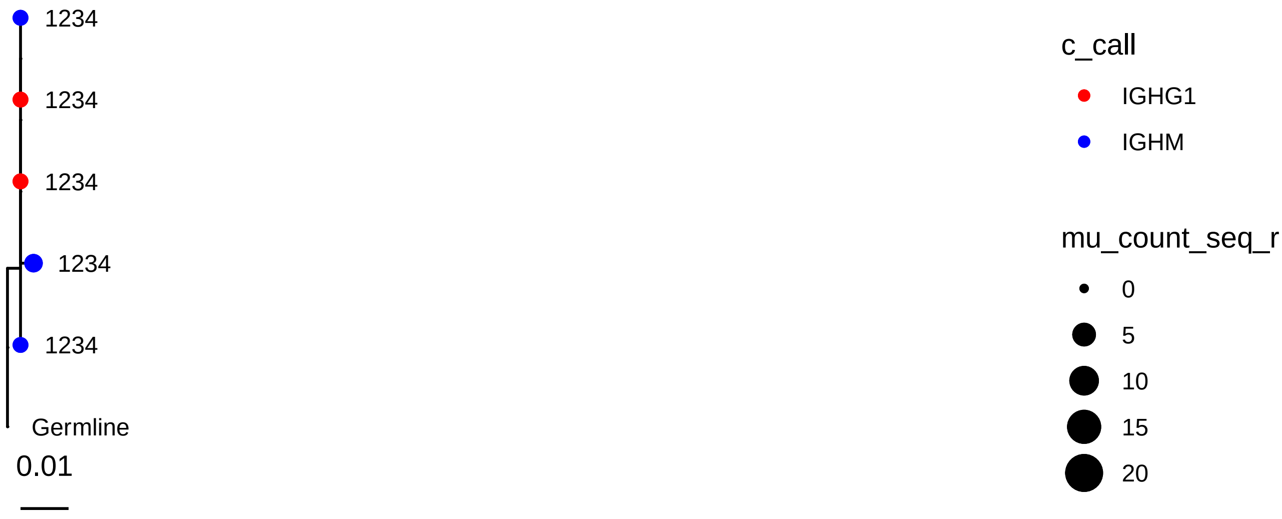
1234
1234
1234
1234
1234
Germline
0.01
c_call
IGHG1
IGHM
mu_count_seq_r
0
5
10
15
20

3585

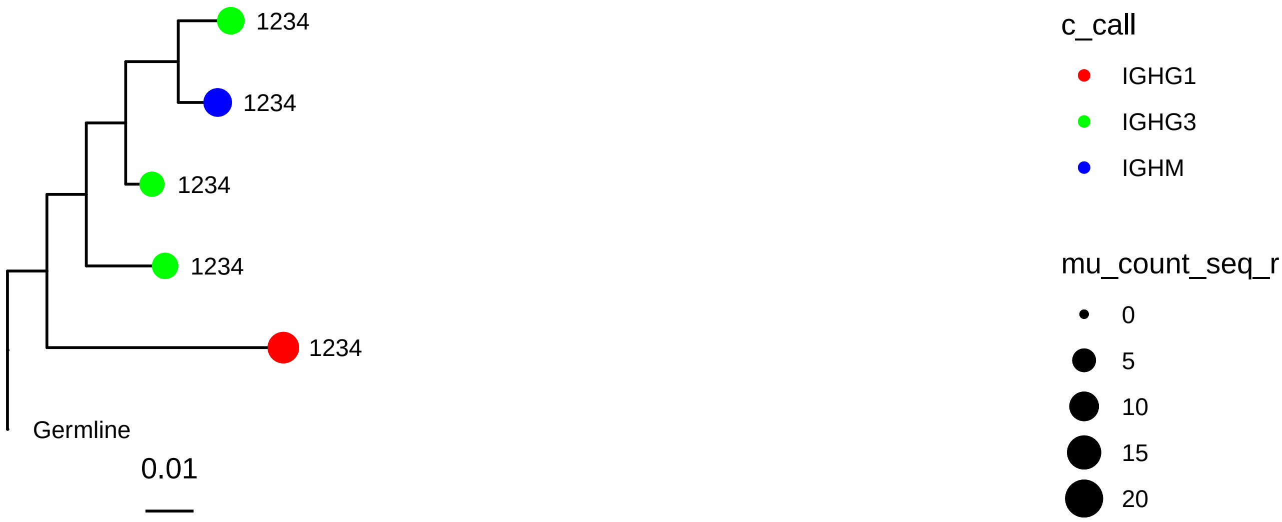
1234
1234
1234
1234
1234
Germline
0.01
c_call
IGHG1
IGHG3
IGHM
mu_count_seq_r
0
5
10
15
20

3762

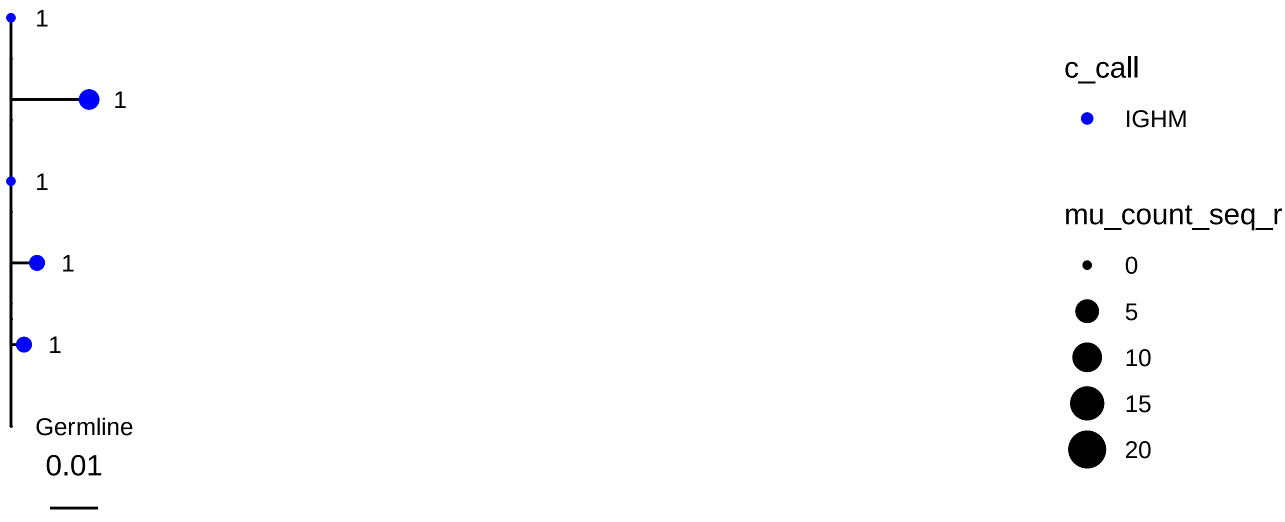
1
1
1
1
1
Germline
0.01
c_call
IGHM
mu_count_seq_r
0
5
10
15
20

3969

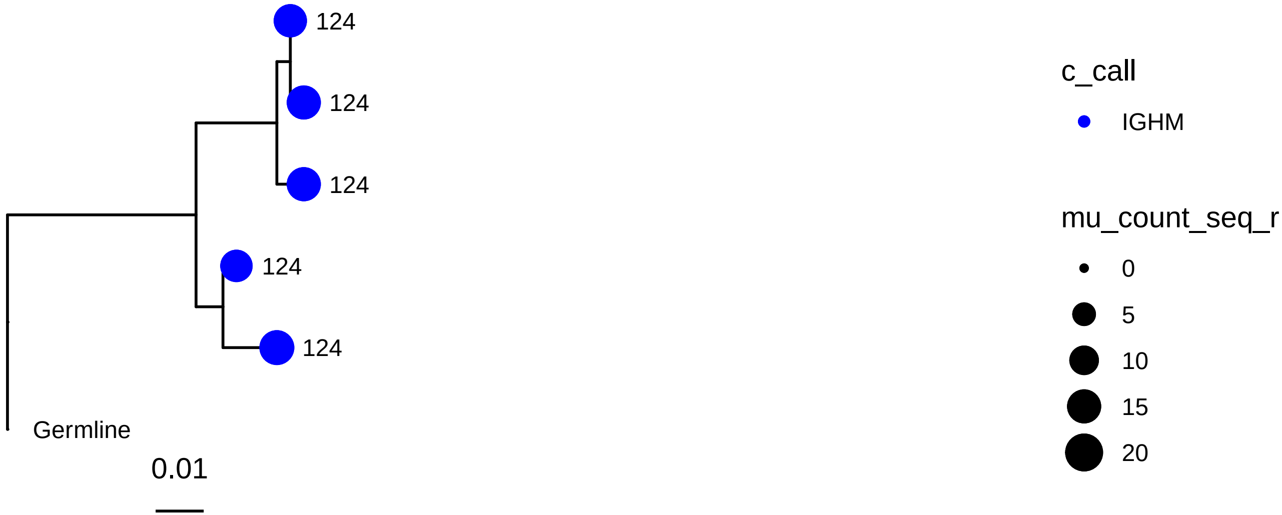
124
124
124
124
124
Germline
0.01
c_call
IGHM
mu_count_seq_r
0
5
10
15
20

4063

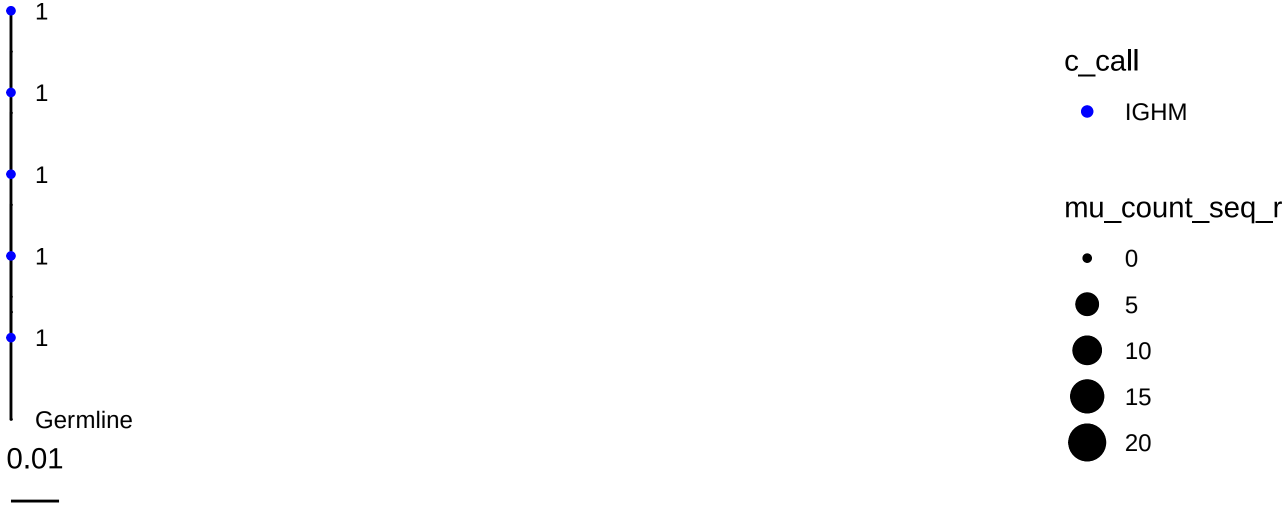
1
1
1
1
1
Germline
0.01
c_call
IGHM
mu_count_seq_r
0
5
10
15
20

4239

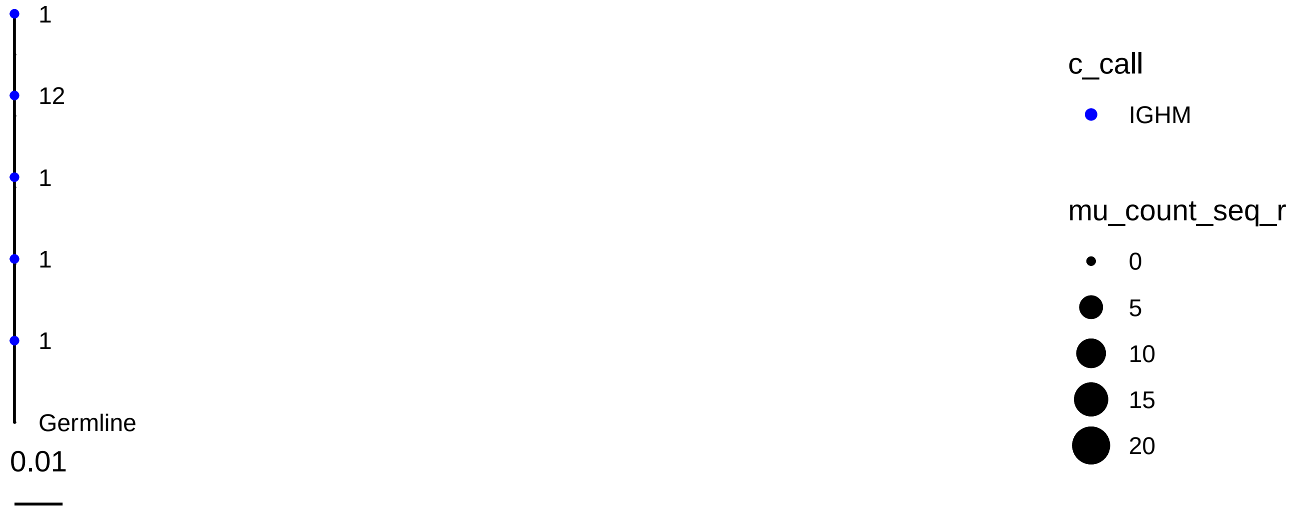
1
12
1
1
1
Germline
0.01
c_call
IGHM
mu_count_seq_r
0
5
10
15
20

4298

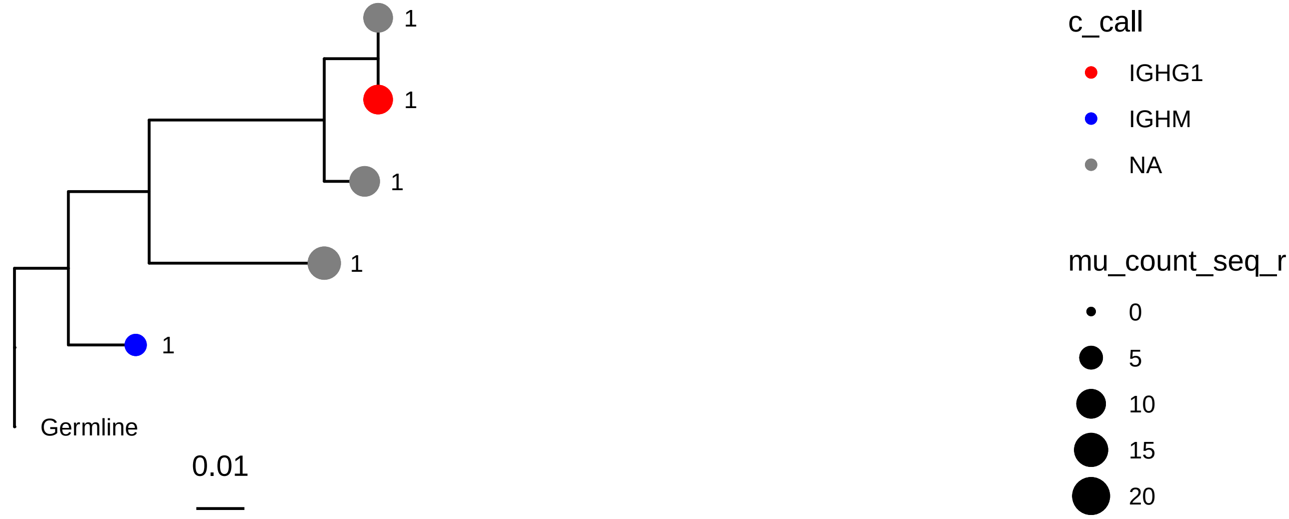
1
1
1
1
1
Germline
0.01
c_call
IGHG1
IGHM
NA
mu_count_seq_r
0
5
10
15
20

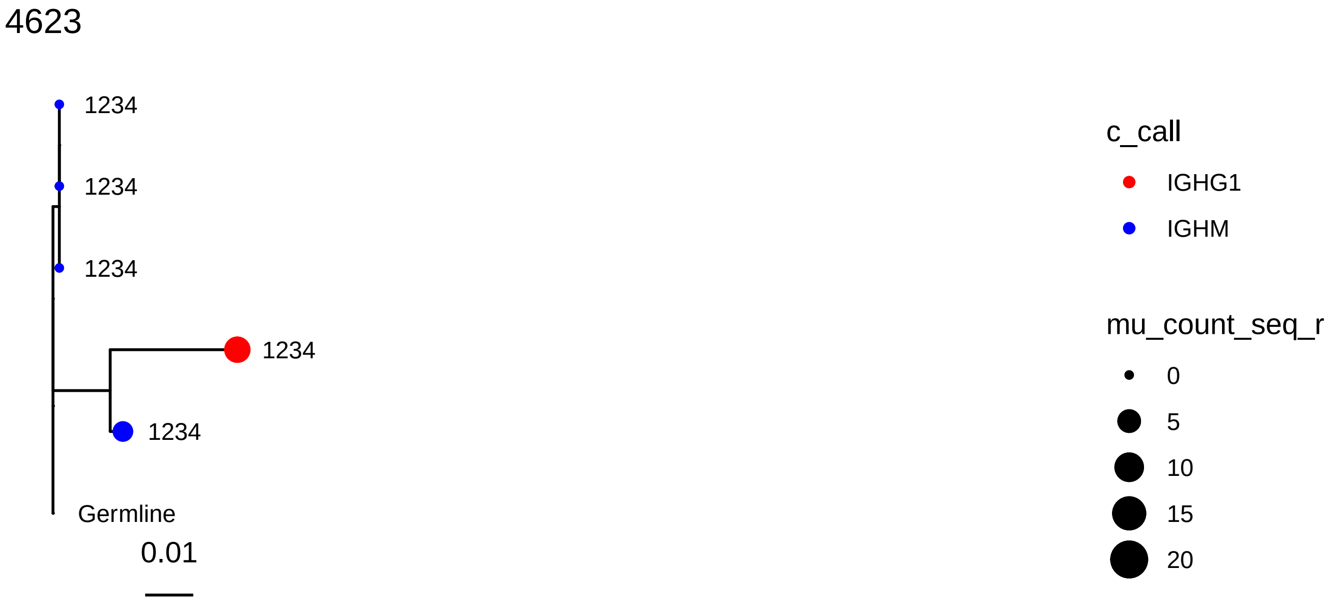
4623
1234
1234
1234
1234
1234
Germline
0.01
c_call
IGHG1
IGHM
mu_count_seq_r
0
5
10
15
20

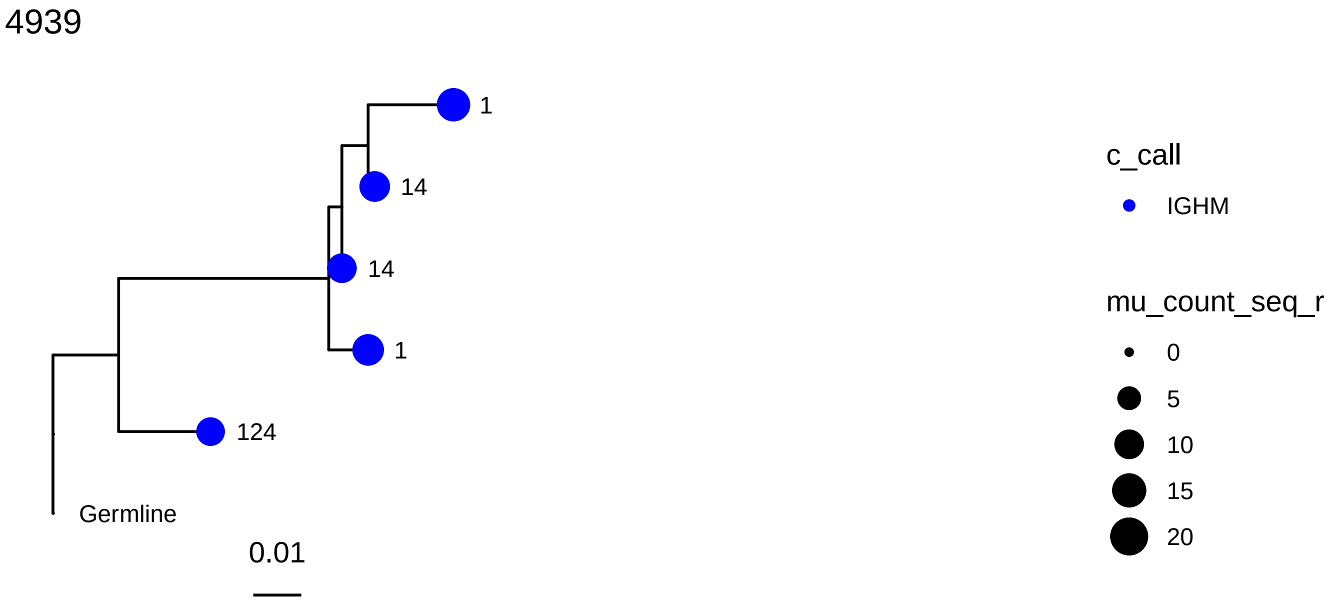
4939
1
14
14
1
124
Germline
0.01
c_call
IGHM
mu_count_seq_r
0
5
10
15
20

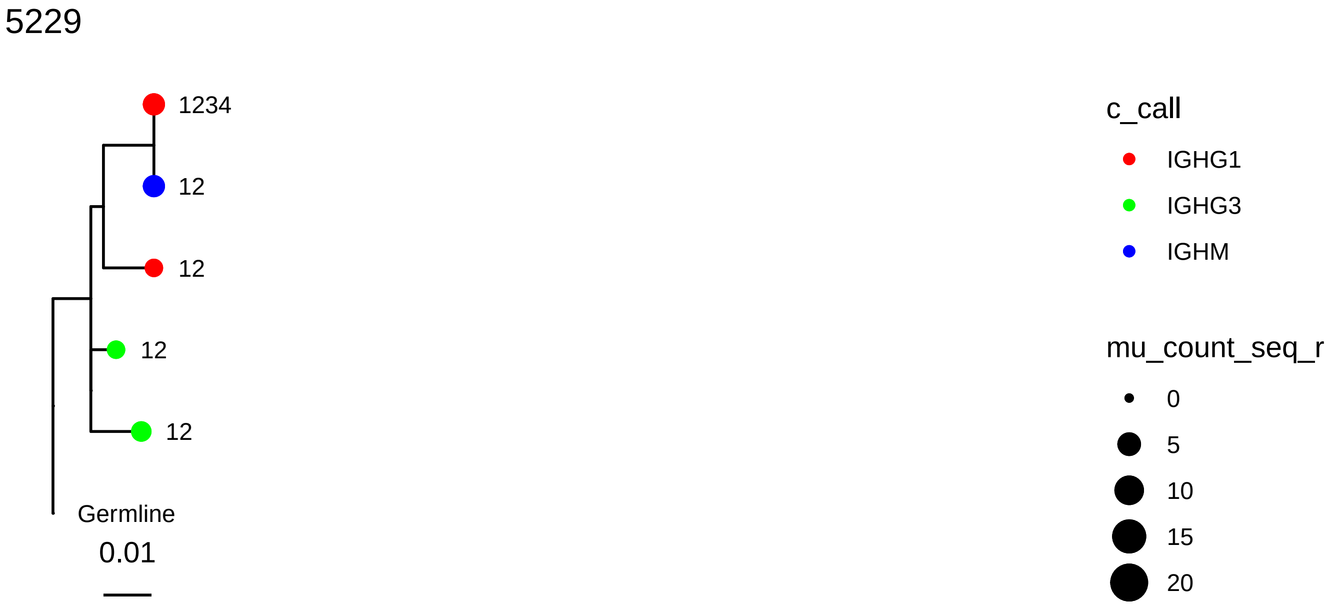
5229
1234
12
12
12
12
Germline
0.01
c_call
IGHG1
IGHG3
IGHM
mu_count_seq_r
0
5
10
15
20

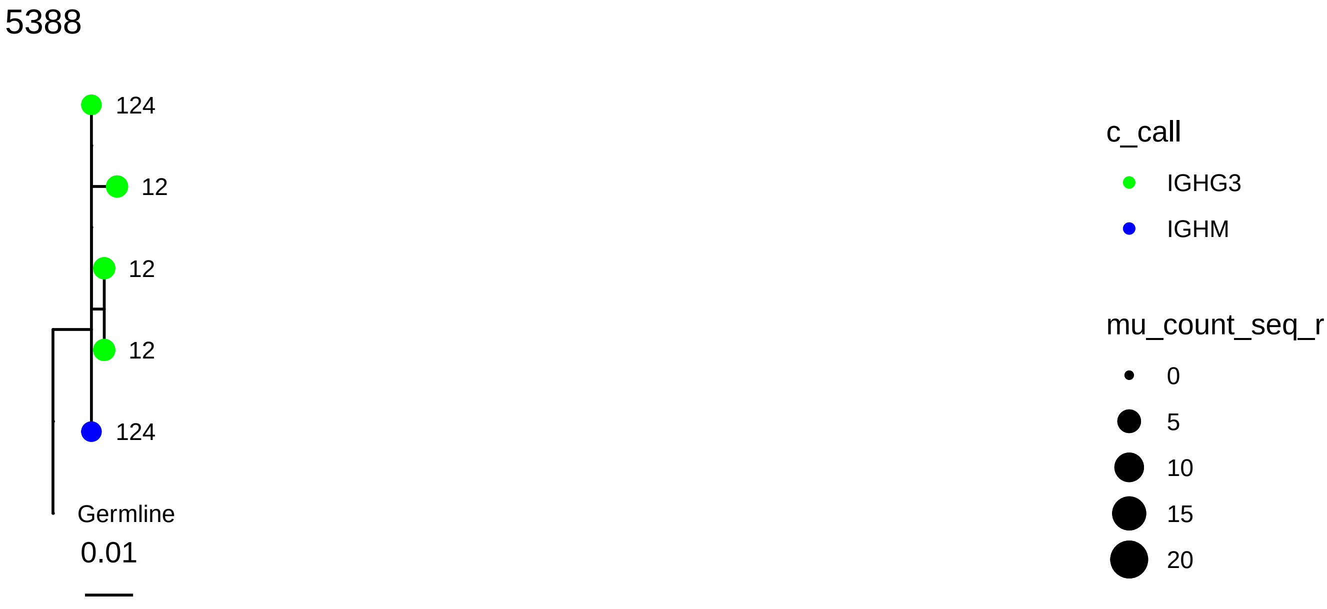
5388
124
12
12
12
124
Germline
0.01
c_call
IGHG3
IGHM
mu_count_seq_r
0
5
10
15
20

5475

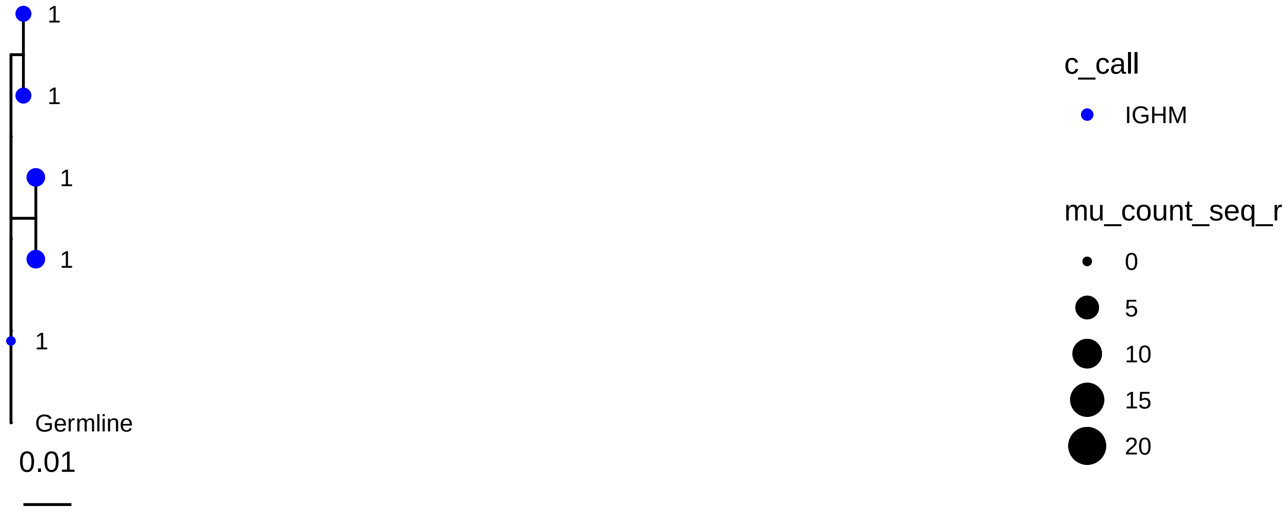
1
1
1
1
1
Germline
0.01
c_call
IGHM
mu_count_seq_r
0
5
10
15
20

5559

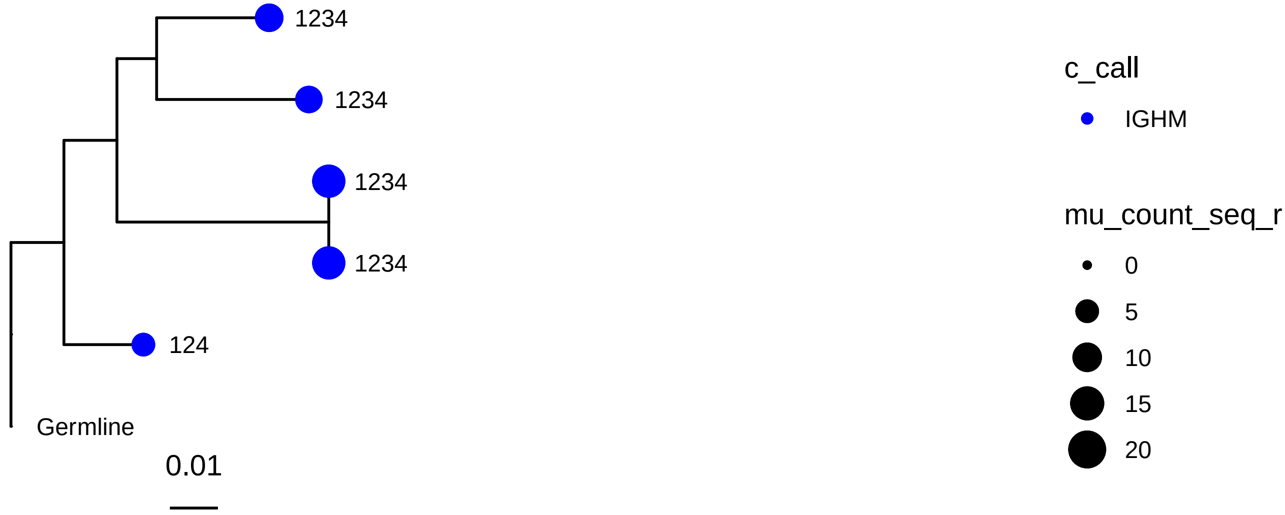
1234
1234
1234
1234
124
Germline
0.01
c_call
IGHM
mu_count_seq_r
0
5
10
15
20
